# Fine-mapping causal tissues and genes at disease-associated loci

**DOI:** 10.1101/2023.11.01.23297909

**Authors:** Benjamin J. Strober, Martin Jinye Zhang, Tiffany Amariuta, Jordan Rossen, Alkes L. Price

**Affiliations:** Department of Epidemiology, Harvard T.H. Chan School of Public Health, Boston, MA, USA; Computational Biology Department, School of Computer Science, Carnegie Mellon University, Pittsburgh, PA, USA; Halicioglu Data Science Institute, University of California San Diego, La Jolla, CA, USA; Department of Medicine, University of California San Diego, La Jolla, CA, USA; Program in Medical and Population Genetics, Broad Institute of MIT and Harvard, Cambridge, MA, USA; Department of Biostatistics, Harvard T.H. Chan School of Public Health, Boston, MA, USA

## Abstract

Heritable diseases often manifest in a highly tissue-specific manner, with different disease loci mediated by genes in distinct tissues or cell types. We propose Tissue-Gene Fine-Mapping (TGFM), a fine-mapping method that infers the posterior probability (PIP) for each gene-tissue pair to mediate a disease locus by analyzing GWAS summary statistics (and in-sample LD) and leveraging eQTL data from diverse tissues to build cis-predicted expression models; TGFM also assigns PIPs to causal variants that are not mediated by gene expression in assayed genes and tissues. TGFM accounts for both co-regulation across genes and tissues and LD between SNPs (generalizing existing fine-mapping methods), and incorporates genome-wide estimates of each tissue’s contribution to disease as tissue-level priors. TGFM was well-calibrated and moderately well-powered in simulations; unlike previous methods, TGFM was able to attain correct calibration by modeling uncertainty in cis-predicted expression models. We applied TGFM to 45 UK Biobank diseases/traits (average *N* = 316K) using eQTL data from 38 GTEx tissues. TGFM identified an average of 147 PIP > 0.5 causal genetic elements per disease/trait, of which 11% were gene-tissue pairs. Implicated gene-tissue pairs were concentrated in known disease-critical tissues, and causal genes were strongly enriched in disease-relevant gene sets. Causal gene-tissue pairs identified by TGFM recapitulated known biology (e.g., *TPO*-thyroid for Hypothyroidism), but also included biologically plausible novel findings (e.g., *SLC20A2*-artery aorta for Diastolic blood pressure). Further application of TGFM to single-cell eQTL data from 9 cell types in peripheral blood mononuclear cells (PBMC), analyzed jointly with GTEx tissues, identified 30 additional causal gene-PBMC cell type pairs at PIP > 0.5—primarily for autoimmune disease and blood cell traits, including the biologically plausible example of *CD52* in classical monocyte cells for Monocyte count. In conclusion, TGFM is a robust and powerful method for fine-mapping causal tissues and genes at disease-associated loci.

## Introduction

Heritable diseases often manifest in a highly tissue-specific manner, motivating intense efforts to elucidate tissue-specific mechanisms of disease^1^. Previous studies have identified disease-critical tissues/cell-types based on genome-wide patterns^2–11^, and have deeply dissected a limited number of GWAS loci^12–16^. However, different GWAS loci may be mediated by different tissues, motivating genome-wide efforts to fine-map causal tissues and genes at individual GWAS loci.

Existing approaches, including colocalization^17–19^ and transcriptome wide association studies (TWAS)^20–22^, have implicated disease genes via the integration of GWAS data with expression quantitative trait loci (eQTLs) while considering the effect of each gene-tissue pair on disease in isolation. However, it is likely that most of these disease-implicated genes are not actually causal in the analyzed tissue; analogous to non-causal tagging variants implicated by linkage disequilibrium (LD) between variants^23^, non-causal gene-tissue pairs can be implicated by correlations with causal gene-tissue pairs (involving a different gene and/or different tissue)^11,22,24–27^. In addition, false-positive gene-tissue pairs can arise from correlations with non-mediated genetic variants, i.e., variants whose causal effects are not mediated by assayed expression levels^22,27,28^. Previous fine-mapping approaches such as FOCUS^24^ and cTWAS^27^ have proven valuable in disentangling causal effects across correlated genes in a single tissue, but have not considered causal gene-tissue pairs.

Here, we introduce a new method, Tissue-Gene Fine-Mapping (TGFM), that infers the posterior inclusion probability (PIP) for each gene-tissue pair to mediate a disease association at a given locus; TGFM also assigns PIPs to causal genetic variants whose effects are not mediated by gene expression in assayed tissues and genes. TGFM models both gene-tissue pairs (using cis-predicted expression^20,21^) and non-mediated genetic variants as potential causal genetic elements, and accounts for both correlations in cis-predicted expression across genes and tissues and LD between genetic variants, generalizing existing fine-mapping methods^23,24,27,29–31^. TGFM incorporates genome-wide estimates of each tissue’s contribution to disease as tissue-level priors and employs a sampling approach to account for uncertainty in cis-predicted gene expression. We validated TGFM using extensive simulations with real genotypes, including comparisons to coloc^17^,FOCUS^24^, and cTWAS^27^. We applied TGFM to 45 UK Biobank traits^32^ using eQTL data from 38 GTEx tissues^25^ and 9 fine-grained single-cell PBMC cell-types^33^.

## Results

### Overview of TGFM

TGFM estimates the posterior inclusion probability (PIP) for each *genetic element* (gene-tissue pair or genetic variant) to have a non-zero causal effect on disease, in a model that includes *mediated* causal effects of each gene-tissue pair (via the cis-genetic component of expression of a given gene in a given tissue) and *non-mediated* causal effects of each genetic variant:

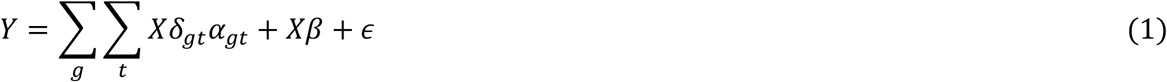

where *Y* denotes phenotypes, *g* indexes genes, *t* indexes tissues, *X* is the matrix of genotypes, *δ*_*gt*_ is the vector of causal cis-eQTL effect sizes of each variant on gene expression in gene *g* and tissue *t* (thus *Xδ*_*gt*_ is the cis-genetic component of gene expression in gene *g* and tissue *t*), *α*_*gt*_ denotes the (scalar) effect of cis-genetic expression in gene *g* and tissue *t* on the disease or trait, *β* is the vector of non-mediated causal effects of each genetic variant on the disease or trait, and *ϵ* denotes environmental noise. We caution that, analogous to previous studies, inference of causal genetic elements relies on the assumption that all causal genetic elements have been assayed, which may not be true in practice (see Discussion); we use the word “causal” for simplicity, with this caveat in mind.

TGFM estimates the PIP of each genetic element by generalizing the Sum of Single Effects (SuSiE)^30,31^ fine-mapping method to include both gene-tissue pairs and genetic variants; gene-tissue pairs are included via cis-*predicted* expression^20,21^ (using an external eQTL data set such as GTEx^25^ to build prediction models), which is an approximation to true cis-*genetic* expression (Methods). This approach allows for fine-mapping multiple causal genetic elements in a given locus, inferring causal effects underlying both marginal GWAS^34^ and marginal TWAS^20–22^ (i.e., the association between cis-predicted expression of a single gene-tissue pair and disease) associations by accounting for correlations between gene-tissue pairs due to co-regulation across genes/tissues^11,22,24–27^, correlations between genetic variants due to LD^23^, and/or correlations between gene-tissue pairs and genetic variants due to the inclusion of a genetic variant in a model of cis-predicted gene expression^22,27,28^. TGFM employs a sampling approach to account for uncertainty in cis-predicted expression, avoiding false positives that arise from noisy estimation of cis-genetic expression.

In detail, TGFM consists of four steps. In step 1, we apply SuSiE to perform eQTL fine-mapping of each gene-tissue pair in the external gene expression data set (estimating a posterior distribution of the causal cis-eQTL effect sizes for each gene-tissue pair). In step 2, we randomly sample 100 cis-predicted expression models for each gene-tissue pair from the posterior distributions of causal cis-eQTL effect sizes estimated in step 1 (Methods). In step 3, we apply SuSiE to perform disease fine-mapping in the target data set (estimating the PIP of each genetic element) 100 times, iterating over the sampled cis-predicted expression models for each gene-tissue pair from step 2. In step 4, we average the results of step 3 across the 100 disease fine-mapping runs. TGFM utilizes a custom implementation of the SuSiE algorithm that provides efficient estimation of PIPs across 100 parallel SuSiE runs that differ only in their cis-predicted expression models (Methods). TGFM inference requires only summary-level GWAS data^31,35^ consisting of GWAS z-scores for each variant and in-sample LD between genetic variants, in addition to external eQTL data sets across tissues of interest.

TGFM increases fine-mapping power by specifying tissue-specific prior probabilities for each genetic element in a locus that are informed by genome-wide data, analogous to functionally informed variant-level fine-mapping methods such as PolyFun^36^; TGFM assigns one prior causal probability *π*_*t*_ for each gene-tissue pair from tissue *t* and one prior causal probability *π*_*nm*_ for each non-mediated genetic variant. We estimate *π*_*t*_ and *π*_*nm*_ in each disease/trait separately by iteratively running a computationally efficient approximation to TGFM (Methods), starting with flat priors and updating *π*_*t*_ and *π*_*nm*_ at each iteration until convergence. When analyzing a given locus with TGFM, we normalize the prior causal probabilities to sum to 1, analogous to PolyFun^36^. We account for uncertainty in estimates of *π*_*t*_ and *π*_*nm*_ by using genomic bootstrapping, randomly sampling 100 sets of values of *π*_*t*_ and *π*_*nm*_ (one for each of the 100 disease fine-mapping runs in step 3) and averaging TGFM results across the random samples.

We restrict cis-predicted expression models to cis-eQTLs within 500kb of each gene’s transcription start site (TSS). We only assign cis-predicted expression models to gene-tissue pairs that are well-predicted by genetic variants, using the SuSiE “purity filter”^30^ (see Methods). We apply TGFM to fine-map any of the 2,682 overlapping 3Mb loci spanning the entire genome^36^ that contain at least 50 genetic variants and at least one genetic variant with marginal GWAS p-value less than 1e-5. Further details, including sampling cis-predicted expression models from SuSiE posterior distributions of causal cis-eQTL effect sizes, the custom implementation of the SuSiE algorithm providing efficient estimation of PIPs across 100 parallel SuSiE runs, and the computationally efficient approximation to TGFM used when estimating tissue-specific prior causal probabilities, are provided in the Methods section. We have released open-source software implementing TGFM (see Code availability), as well as posterior distributions of causal eQTL effect sizes across tissues and genes, and TGFM PIPs from this study (see Data availability).

### Simulations

We performed simulations using real genotypes to assess the calibration and power of TGFM to identify causal tissues and genes underlying GWAS associations. We used real genotypes from unrelated UK Biobank (UKBB) British samples^32^ to simulate both gene expression phenotypes (for each gene-tissue pair) and quantitative trait phenotypes. Default simulation parameters were specified as follows: the gene expression sample size ranged from 300 to 1000, plus a simulation including tissues with unequal sample sizes (denoted as 100-300), approximately matching the upper and lower range of sample sizes in our analyses of GTEx tissues (see Methods); the quantitative trait sample size was set to 100,000 (disjoint from gene expression samples); we analyzed 426,593 SNPs and 1,976 genes on chromosome 1 (following ref. ^11^); the number of tissues was set to 10, of which 2 were causal for the quantitative trait; the quantitative trait architecture was simulated to have average polygenicity^37^, consisting of 2,700 causal non-mediated variants and 300 causal gene-tissue pairs (150 for each causal tissue) with the expected heritability per causal genetic element (non-mediated variant or gene-tissue pair) set to 0.0001 (expected quantitative trait heritability of 0.3, 10% of which was mediated through gene expression, consistent with genome-wide estimates from MESC^28^); causal non-mediated variants were randomly selected with probability proportional to their expected per-variant heritability based on baseline-LD model annotations^3,38–40^ (estimated using S-LDSC^3^ applied to the UKBB trait White blood cell count) in order to make the simulations as realistic as possible; the genetic architecture of gene expression across tissues was specified following ref. ^11^: roughly, each heritable gene-tissue pair was randomly assigned 5 causal cis-eQTLs (expected per-SNP heritability: 0.015), 2 of the 5 causal eQTLs were specific to each tissue, and 3 of the 5 causal eQTLs were shared across tissues with effect size covariance set to mimic that of GTEx tissues^25^; and causal gene-tissue pairs were randomly selected from all genetically heritable genes (true cis-SNP-heritability > 0) in each of the causal tissues. Non-default simulation parameter values were also explored. We performed 100 independent simulations, and averaged results across simulations. Further details of the simulation framework are provided in the Methods section.

We compared TGFM to three previously published methods, coloc^17^, FOCUS^24^, and cTWAS^27^. Briefly, coloc calculates the posterior probability of a shared causal variant between a GWAS disease/trait and a gene expression trait from a single gene-tissue pair without considering correlations between genes or gene-tissue pairs. Both FOCUS and cTWAS assign PIPs for the expression of each gene in a given tissue to have non-zero causal effect on disease, while modeling correlations between genes in that tissue but not modeling correlations between different tissues and not modeling uncertainty in cis-predicted expression. cTWAS additionally models correlations between genes and non-mediated genetic variants. Both FOCUS and cTWAS can naturally be extended to model correlations between all gene-tissue pairs (without modeling correlations between genes and non-mediated genetic variants, in the case of FOCUS); we refer to the resulting methods as FOCUS-TG and cTWAS-TG, respectively. (In contrast, coloc does not model correlations between genes, and cannot be extended in this way.)

We first evaluated the calibration of TGFM, coloc, FOCUS, FOCUS-TG, cTWAS, and cTWAS-TG to fine-map causal gene-tissue pairs. Calibration was assessed using empirical false discovery rate (FDR), estimated as the proportion of false-positive gene-tissue pairs among all gene-tissue pairs above a given PIP threshold, where a false-positive gene-tissue pair is defined as not having a simulated causal effect on the trait. Following ref. ^36^, we assessed whether the empirical FDR is less than or equal to (1 – PIP threshold), a more conservative choice than (1 – average PIP) (which has been shown to be slightly mis-calibrated in previous fine-mapping simulations^36,41^; also see Methods and secondary analyses below). Results are reported in Figure 1a-b and Supplementary Table 1. TGFM produced well-calibrated PIPs at all eQTL sample sizes and PIP thresholds, with slightly increased false positive rates at lower eQTL sample sizes. In contrast, coloc, FOCUS, FOCUS-TG, cTWAS, and cTWAS-TG were poorly calibrated across all PIP thresholds, even at large eQTL sample sizes. We attribute the superior calibration of TGFM over other approaches to its joint modeling of gene-tissue pairs and non-mediated variants, as well as its sampling procedure that accounts for uncertainty in genetically predicted gene expression (see secondary analyses below). Results were similar at other PIP thresholds (Supplementary Figure 1).

**Table 1:**
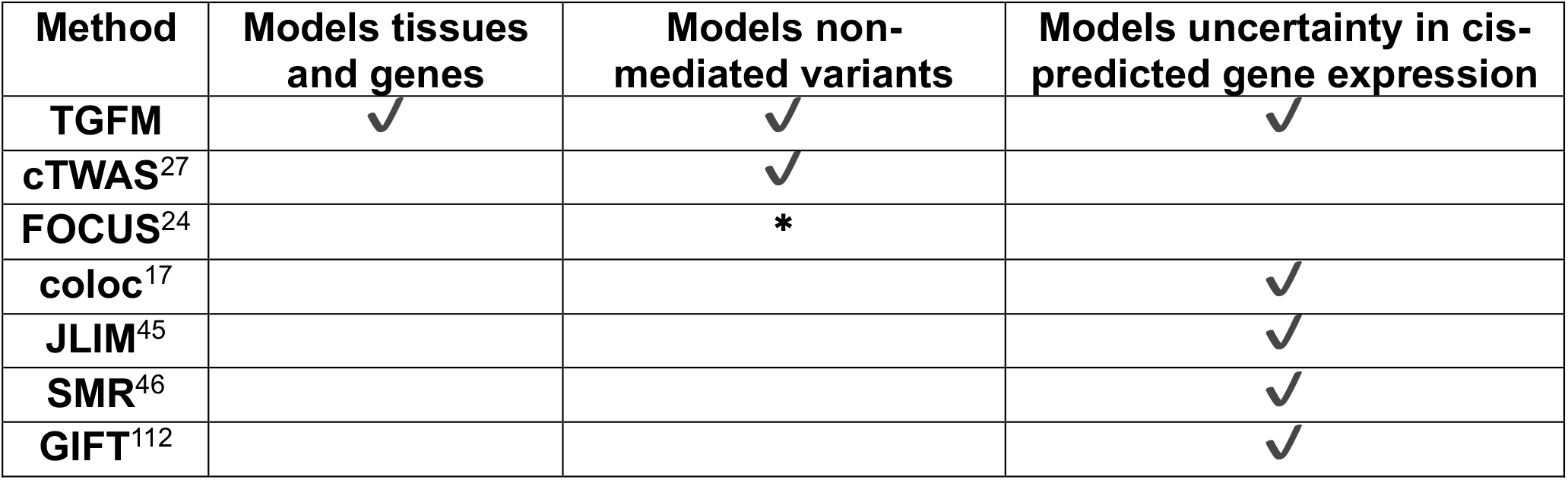
Properties of disease gene fine-mapping methods. For each disease gene fine-mapping method, we report whether or not the method jointly models tissues and genes; models non-mediated variants; and models uncertainty in cis-predicted gene expression. *: FOCUS allows for modeling of non-mediated genetic effects via a single genotype intercept term shared across all variants, but this functionality is not enabled in the default version of FOCUS (and does not ameliorate mis-calibration; Supplementary Figure 23).

| Method | Models tissues and genes | Models non-mediated variants | Models uncertainty in cis-predicted gene expression |
| --- | --- | --- | --- |
| TGFM | ✓ | ✓ | ✓ |
| cTWAS<sup>27</sup> |  | ✓ |  |
| FOCUS<sup>24</sup> |  | * |  |
| coloc<sup>17</sup> |  |  | ✓ |
| JLIM<sup>45</sup> |  |  | ✓ |
| SMR<sup>46</sup> |  |  | ✓ |
| GIFT<sup>112</sup> |  |  | ✓ |

**Figure 1:**
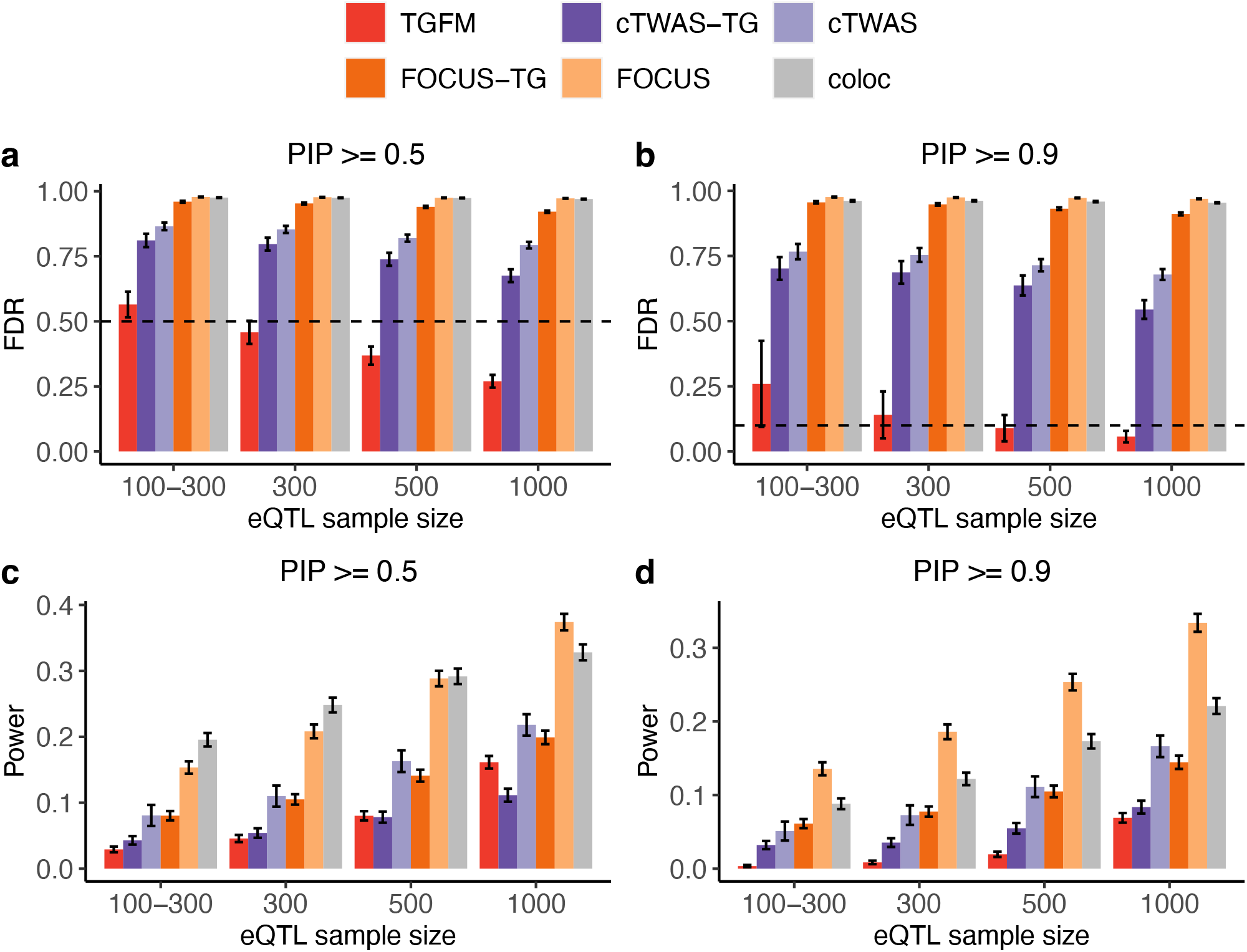
Calibration and power of tissue-gene fine-mapping methods in simulations. **(a,b)** Average gene-tissue pair fine-mapping FDR across 100 simulations for various fine-mapping methods (see legend) across eQTL sample sizes (x-axis) at PIP=0.5 **(a)** and PIP=0.9 **(b)**. Dashed horizontal line denotes 1 – PIP threshold (see main text). Numerical results are reported in Supplementary Table 1. **(c,d)** Average gene-tissue pair fine-mapping power across 100 simulations for various fine-mapping methods (see legend) across eQTL sample sizes (x-axis) at PIP=0.5 **(c)** and PIP=0.9 **(d)**. Error bars denote 95% confidence intervals. Numerical results are reported in Supplementary Table 2.

We next evaluated the power of TGFM, coloc, FOCUS, FOCUS-TG, cTWAS, and cTWAS-TG to fine-map causal gene-tissue pairs. Results are reported in Figure 1c-d and Supplementary Table 2. TGFM was moderately well-powered to detect causal gene-tissue pairs at larger eQTL sample sizes, with power ranging from 0.03-0.16 across eQTL sample sizes at a PIP threshold of 0.5. Other methods (coloc, FOCUS, FOCUS-TG, cTWAS, and cTWAS-TG) achieved higher power than TGFM, but this is largely moot due to the poor calibration of those methods (Figure 1a-b). At the same level of FDR, TGFM attained higher power than coloc, FOCUS, FOCUS-TG, cTWAS, and cTWAS-TG (Supplementary Figure 2). Results were similar at other PIP thresholds (Supplementary Figure 3).

We compared the calibration and power of TGFM for fine-mapping causal *gene-tissue pairs, genes*, or *non-mediated genetic variants*. Gene PIPs were computed by aggregating gene-tissue PIPs across all gene-tissue pairs corresponding to the gene (defining a gene as causal if at least one corresponding gene-tissue pair is causal; Methods). A false-positive gene-tissue pair is defined as not having a simulated causal effect on the trait (see above), a false-positive gene is defined as not having a simulated causal effect on the trait for any tissue, and a false-positive variant is defined as not having a simulated causal non-mediated effect on the trait and not being a causal eQTL variant for a causal gene-tissue pair (also see secondary analyses below); however, causal eQTL variants for a causal gene-tissue pair were not included as true positives in variant-level power computations. Calibration results for TGFM (Gene-Tissue), TGFM (Gene) and TGFM (Variant) are reported in Figure 2a-b and Supplementary Table 3.

**Figure 2:**
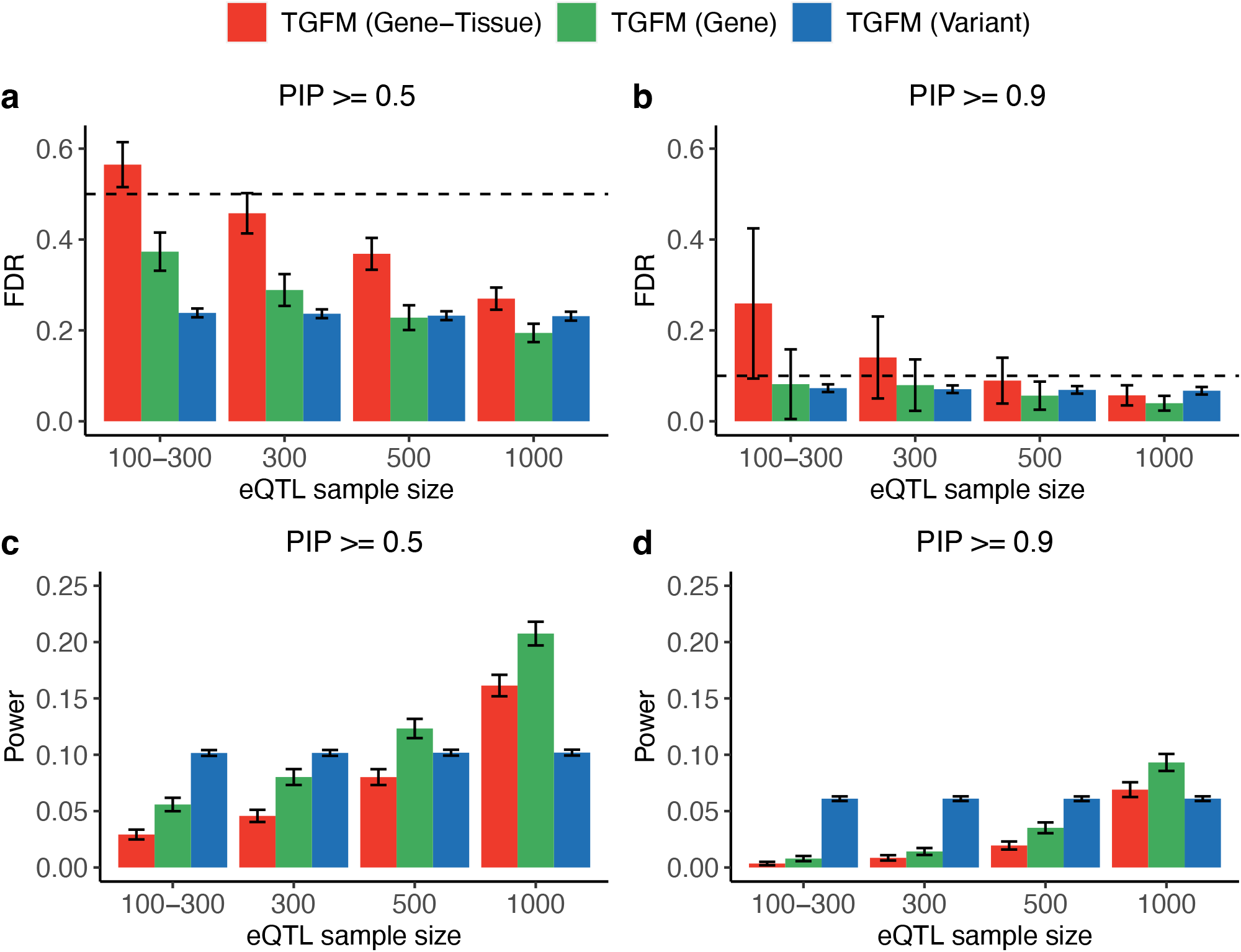
Calibration and power of fine-mapping different classes of genetic elements with TGFM in simulations. **(a,b)** Average fine-mapping FDR across 100 simulations using TGFM for different classes of genetic elements (see legend) across eQTL sample sizes (x-axis) at PIP=0.5 **(a)** and PIP=0.9 **(b)**. Dashed horizontal line denotes 1 – PIP threshold (see main text). Numerical results are reported in Supplementary Table 3. **(c,d)** Average fine-mapping power across 100 simulations using TGFM for different classes of genetic elements (see legend) across eQTL sample sizes (x-axis) at PIP=0.5 **(c)** and PIP=0.9 **(d)**. Error bars denote 95% confidence intervals. Numerical results are reported in Supplementary Table 4.

TGFM produced well-calibrated gene-level and variant-level PIPs. In contrast, gene-level coloc, FOCUS, FOCUS-TG, cTWAS, and cTWAS-TG PIPs were poorly calibrated across all PIP thresholds even at large eQTL sample sizes (Supplementary Figure 4, analogous to Figure 1a-b). Results were generally similar at other PIP thresholds (Supplementary Figure 5), although TGFM (Variant) PIPs were slightly mis-calibrated at very high PIP thresholds (PIP=0.99), consistent with variant-level fine-mapping methods^41^.

Power results for TGFM (Gene-Tissue), TGFM (Gene) and TGFM (Variant) are reported in Figure 2c-d and Supplementary Table 4. TGFM attained higher power to fine-map causal genes than causal gene-tissue pairs, which is expected as fine-mapping causal genes is an easier problem. Power for variant-level fine-mapping was invariant to eQTL sample size, such that variant-level fine-mapping was more powerful than gene-tissue or gene-level fine-mapping at smaller eQTL sample sizes—particularly at the stringent PIP>0.9 threshold, at which the latter were severely underpowered. Results were similar at other PIP thresholds (Supplementary Figure 6).

We performed 10 secondary analyses involving additional evaluations of TGFM, coloc, FOCUS, FOCUS-TG, cTWAS, and cTWAS-TG. First, we modified our calibration analyses to assess whether the empirical FDR is less than or equal to (1 – average PIP), a less conservative choice than (1 – PIP threshold) (ref. ^36,41^; see Methods). We determined that TGFM (Gene-Tissue) was slightly mis-calibrated only at small eQTL sample sizes, TGFM (Gene) was well-calibrated across all eQTL sample sizes and PIP thresholds analyzed, TGFM (Variant) was slightly mis-calibrated at high PIP thresholds regardless of eQTL sample size, and coloc, FOCUS, FOCUS-TG, cTWAS, and cTWAS-TG were severely miscalibrated across all eQTL sample sizes and PIP thresholds analyzed (Supplementary Figures 1, 5). The slight miscalibration of TGFM (Gene-Tissue) and TGFM (Variant) when using (1 – average PIP) is consistent with previous simulations of variant-level fine-mapping methods using polygenic trait architectures^36,41^. Second, we considered a simulation in which the number of causal eQTLs and the cis-heritability varied across gene-tissue pairs, so that for every gene the number of causal eQTLs and cis-heritability varies across tissues. Specifically, the number of causal eQTLs for a given gene-tissue pair was randomly selected to be between 3 and 7 (instead of fixed at 5 as in our primary simulations). TGFM remained well-calibrated, and attained comparable power relative to our primary simulation (Supplementary Figure 7). Third, we considered a simulation in which causal eQTLs are shared across nearby genes^42^ (and shared across tissues, as in our primary simulations). Specifically, in each simulation we randomly selected 250 pairs of genes with overlapping cis-windows (of 1,976 genes total) to have 2 shared causal eQTLs, with effect size covariance equal to the effect size covariance for shared causal eQTLs across tissues for a single gene (as described above). TGFM remained well-calibrated, and attained comparable power relative to our primary simulation (Supplementary Figure 8). Fourth, we considered a simulation in which eQTLs for disease-causal genes (actually gene-tissue pairs) had lower eQTL effect sizes, thus explaining lower cis-heritability, as may be expected due to selection^26,28,43,44^. Specifically, the expected per-SNP heritability for a causal cis-eQTL of a causal gene-tissue pair was set to 0.01 (vs. 0.015 for a causal cis-eQTL of a non-causal gene-tissue pair). TGFM became slightly mis-calibrated, however, TGFM’s calibration remained tolerable and notably superior to all other approaches (Supplementary Figure 9). As expected, TGFM attained decreased power, analogous to simulations with decreased eQTL sample size or decreased cis-heritability. Fifth, we considered a simulation with a single causal tissue per trait (instead of 2 causal tissues). TGFM remained well-calibrated and attained slightly increased power, while other methods remained severely mis-calibrated (Supplementary Figures 10, 11). Sixth, we restricted our evaluation of our primary simulations to loci containing at most 1 causal gene-tissue pair. TGFM remained well-calibrated, whereas coloc, FOCUS, and cTWAS were strongly mis-calibrated (Supplementary Figure 12); this demonstrates that the advantages of TGFM over other methods are not limited to identifying multiple causal gene-tissue pairs at a locus. Seventh, we considered a simulation with low eQTL sample size, equal to 100 in all tissues. TGFM became slightly mis-calibrated, however, TGFM’s calibration remained tolerable and notably superior to all other approaches (Supplementary Figures 13, 14; see Discussion for additional discussion of low eQTL sample size). Eighth, we ran TGFM at different simulated GWAS sample sizes ranging from 50,000 to 200,000 (instead of the default sample size of 100,000). TGFM remained well-calibrated regardless of GWAS sample size but attained increased power at larger GWAS sample sizes (Supplementary Figure 15). TGFM fine-mapping power at PIP > 0.5 increased 2.0-fold when doubling the eQTL sample size vs. 1.2-fold when doubling the GWAS sample size (relative to an eQTL sample size of 500 and GWAS sample size of 100,000), suggesting that TGFM attains a greater benefit from increasing the eQTL sample size under our default parameter settings. Ninth, we ran TGFM at different values of heritability of gene expression, ranging from 0.05 to 0.1 (instead of the default value of 0.075). TGFM remained approximately well-calibrated regardless of gene expression heritability but attained increased power at larger values of gene expression heritability (Supplementary Figure 16), analogous to the impact of varying eQTL sample size (Figure 2). Tenth, we performed an alternative calibration analysis of TGFM (Variant) where causal eQTL variants for causal gene-tissue pairs were considered false positives for variant-level calibration. TGFM (Variant) was mis-calibrated at small eQTL sample sizes and high PIP thresholds in this alternative calibration analysis (Supplementary Figure 17). The calibration worsened at small eQTL sample sizes, likely due to decreased power to detect causal gene-tissue pairs at small eQTL sample sizes, forcing the unmodeled gene-tissue pair effects to be captured by non-mediated variants.

We performed 13 secondary analyses involving additional methods. First, we compared TGFM to two additional methods, JLIM^45^ and SMR (with or without the HEIDI filter)^46^; JLIM tests the existence of a shared causal variant between a GWAS disease/trait and a gene expression trait from a single gene-tissue pair, and SMR tests whether a disease/trait is associated with a single gene-tissue pair because of shared causal variants. JLIM and SMR (with or without the HEIDI filter) were severely mis-calibrated, with high FDR at even the most stringent p-value thresholds (Supplementary Figure 2). Furthermore, TGFM attained higher power than JLIM and SMR (with or without the HEIDI filter) at the same level of FDR (Supplementary Figure 2). Second, we ran TGFM with a uniform prior (same *π*_*nm*_ and *π*_*t*_ for all tissues) instead of the default tissue-specific priors inferred from genome-wide data. TGFM with a uniform prior remained well-calibrated (Supplementary Figure 18a-b) but suffered substantially reduced power (Supplementary Figure 18c-d), highlighting the benefit of tissue-specific priors informed by genome-wide data. Third, we ran TGFM with a uniform prior and a single cis-predicted expression model (based on posterior mean causal cis-eQTL effect sizes) instead of averaging results across 100 sampled cis-predicted expression models.

TGFM without sampling cis-predicted expression models suffered poor calibration, particularly at smaller eQTL sample sizes (Supplementary Figure 18a-b), highlighting the advantages of the sampling approach to account for uncertainty in cis-predicted expression. However, the calibration of this method was still better than the calibration of FOCUS-TG (Figure 1a-b), perhaps because the default version of FOCUS-TG does not account for non-mediated genetic variants (see below for analyses of non-default versions of FOCUS and FOCUS-TG that model non-mediated genetic effects via a single genotype intercept term shared across all variants); the calibration of this method was also slightly better than the calibration of cTWAS-TG (Figure1a-b), which does account for non-mediated genetic variants. Fourth, we ran TGFM using either 50 or 200 posterior samples, instead of the default 100 posterior samples. TGFM performed similarly at all of these values of the number of posterior samples drawn (Supplementary Figure 19). Fifth, we compared TGFM to a two-step fine-mapping approach that first infers the causal tissue and then performs traditional gene-level fine-mapping in the inferred causal tissue. We inferred the causal tissue using the TGFM tissue-specific prior, and performed gene-level fine-mapping in the inferred causal tissue using either TGFM (applied to consider only a single tissue), coloc, FOCUS, or cTWAS; we refer to these two-step fine-mapping methods as two-step-TGFM, two-step-coloc, two-step-FOCUS, and two-step-cTWAS, respectively. TGFM attained superior calibration relative to two-step coloc, two-step-FOCUS, and two-step-cTWAS in both the primary simulation (two causal tissues) and the single causal tissue simulation (see above), and attained slightly improved calibration relative to two-step-TGFM at high PIP thresholds in the primary simulation (Supplementary Figures 20, 21). Sixth, as a special case of two-step fine-mapping methods, we considered a simulation with a single causal tissue that is known (in contrast to our single causal tissue simulation above) and compared TGFM (applied to consider only a single tissue) to coloc, FOCUS, and cTWAS. TGFM remained well-calibrated, whereas coloc, FOCUS, and cTWAS remained mis-calibrated (Supplementary Figure 22); coloc and FOCUS had substantially worse calibration than cTWAS, consistent with ref. ^27^; this implies that TGFM is an advance over previous methods even within the scope of traditional gene-level fine-mapping. Eighth, we evaluated non-default versions of FOCUS and FOCUS-TG that model non-mediated genetic effects via a single genotype intercept term shared across all variants. FOCUS and FOCUS-TG remained strongly mis-calibrated with the genotype intercept enabled (Supplementary Figure 23). Ninth, we investigated TGFM’s ability to provide unbiased posterior estimates of the proportion of causal genetic elements (gene-tissue pairs or non-mediated variants) that are gene-tissue pairs, a parameter closely related to the proportion of disease heritability mediated by gene expression (estimated in ref. ^28^). We first calculated the proportion of fine-mapped genetic elements at various PIP thresholds that are gene-tissue pairs. This approach yielded either upward or downward biased estimates of the true proportion depending on the eQTL sample size and PIP threshold (Supplementary Figure 24a), reflecting differential discovery power of non-mediated variants and gene-tissue pairs as a function of both eQTL sample size and PIP threshold (Figure 2c-d). We next calculated the *expected* proportion of fine-mapped genetic elements that are gene-tissue pairs by summing PIPs across genetic elements. This approach yielded conservative estimates of the true proportion, becoming less conservative at larger eQTL sample sizes (Supplementary Figure 24b), suggesting that this statistic can provide a conservative lower bound on the true proportion of causal genetic elements that are gene-tissue pairs. Tenth, we assessed the unbiasedness of the prior causal probabilities inferred by the TGFM tissue-specific prior. The prior causal probabilities for (gene-tissue pairs involving) causal tissues were approximately unbiased, but the prior causal probabilities for non-causal tissues and non-mediated variants were upwardly biased (Supplementary Figure 25). Eleventh, we examined the impact of mis-specified prior causal probabilities on TGFM fine-mapping calibration and power. Specifically, we scaled the prior causal probabilities for (gene-tissue pairs involving) non-causal tissues by 0.5 or 2.0, and independently scaled the prior causal probabilities for non-mediated variants by 0.5 or 2.0. TGFM calibration and power to fine-map gene-tissue pairs, genes, and non-mediated variants was largely robust to these mis-specified prior causal probabilities (Supplementary Figures 26, 27). Twelfth, we investigated the calibration and power of inference of disease-critical tissues via the TGFM tissue-specific prior, using genomic bootstrap to assess significance (see Methods); although inference of disease-critical tissues is not a primary goal of TGFM (and there exist previous methods for inferring disease-critical tissues using eQTL data^5,11^), assessing the TGFM tissue-specific prior is of interest. We determined that inference of disease-critical tissues via the TGFM tissue-specific prior was well-calibrated and well-powered (Supplementary Figure 28). Thirteenth, we compared TGFM (Variant) PIPs with variant-level PIPs inferred by SuSiE^30,31^. The variant-level PIPs were strongly correlated and consistent in magnitude, particularly after excluding from the analysis any variant that was correlated with a TGFM fine-mapped gene-tissue pair (Supplementary Figure 29).

### Tissue-gene fine-mapping of 45 diseases and complex traits

We applied TGFM to fine-map tissues and genes for 45 diseases and complex traits from the UK Biobank (average *N* = 316K; previously analyzed with functionally informed variant-level fine-mapping^36^; Methods and Supplementary Table 5) using gene expression data from 47 GTEx tissues^25^, which were aggregated into 38 *meta-tissues*^11^ (average N=259; Supplementary Table 6) to minimize eQTL sample size differences across tissues; below, we refer to these as “tissues” for simplicity. For each disease/trait we applied TGFM to 2,682 overlapping 3-Mb loci^36^ spanning 119,270 (protein-coding) gene-tissue pairs with cis-predicted expression models (3,139 genes per tissue on average, Supplementary Table 6; 13,700 unique genes) and 10,545,304 genetic variants with MAF ≥ 0.005. We assigned a PIP to each gene-tissue pair, gene, and non-mediated genetic variant using the locus in which the genetic element was most central^36^; the position of a gene was determined by its TSS. TGFM running times are reported in Supplementary Table 7. We have publicly released PIPs for all gene-tissue pairs, genes, and non-mediated variants for each disease/trait (see Data Availability).

Results are summarized in Figure 3 (16 independent traits^36^), Supplementary Figure 30 (all 45 traits), and Supplementary Table 8. Across all 45 traits, TGFM identified 711 gene-tissue-trait triplets, 2,800 gene-trait pairs (aggregating gene-tissue PIPs across tissues; see above), and 5,893 non-mediated genetic variant-trait pairs at PIP > 0.5 (43 gene-tissue-trait triplets, 382 gene-trait pairs, and 2,675 non-mediated genetic variant-trait pairs at PIP > 0.9). The number of gene-tissue pairs with PIP > 0.5 ranged from 0 (Number of children) to 56 (White blood cell count) across traits, and ranged from 0 (coronary artery) to 197 (whole blood) across tissues. Of the 711 gene-tissue-trait triplets with PIP > 0.5, 180 (25%) had TWAS p-value > 0.05/119,270 = 4.2 × 10^−7^ (the Bonferroni significance threshold based on 119,270 gene-tissue pairs with cis-predicted expression models^47^) and 136 (19%) had no nearby variants in the same fine-mapping region with GWAS p-value ≤ 5 × 10^−8^. Of the 110,828 gene-tissue-trait triplets with TWAS p-value ≤ 4.2 × 10^−7^, only 531 (0.5%) had TGFM PIP > 0.5. The proportion of causal genetic elements (variants or gene-tissue pairs) that were gene-tissue pairs was equal to 8.1% when counting PIP > 0.5 genetic elements across 16 independent traits (271 gene-tissue pairs and 3,074 non-mediated genetic variants), or 10.1% when summing PIPs across 16 independent traits (Methods), consistent with previous estimates of the proportion of trait heritability mediated by gene expression^28^.

**Figure 3:**
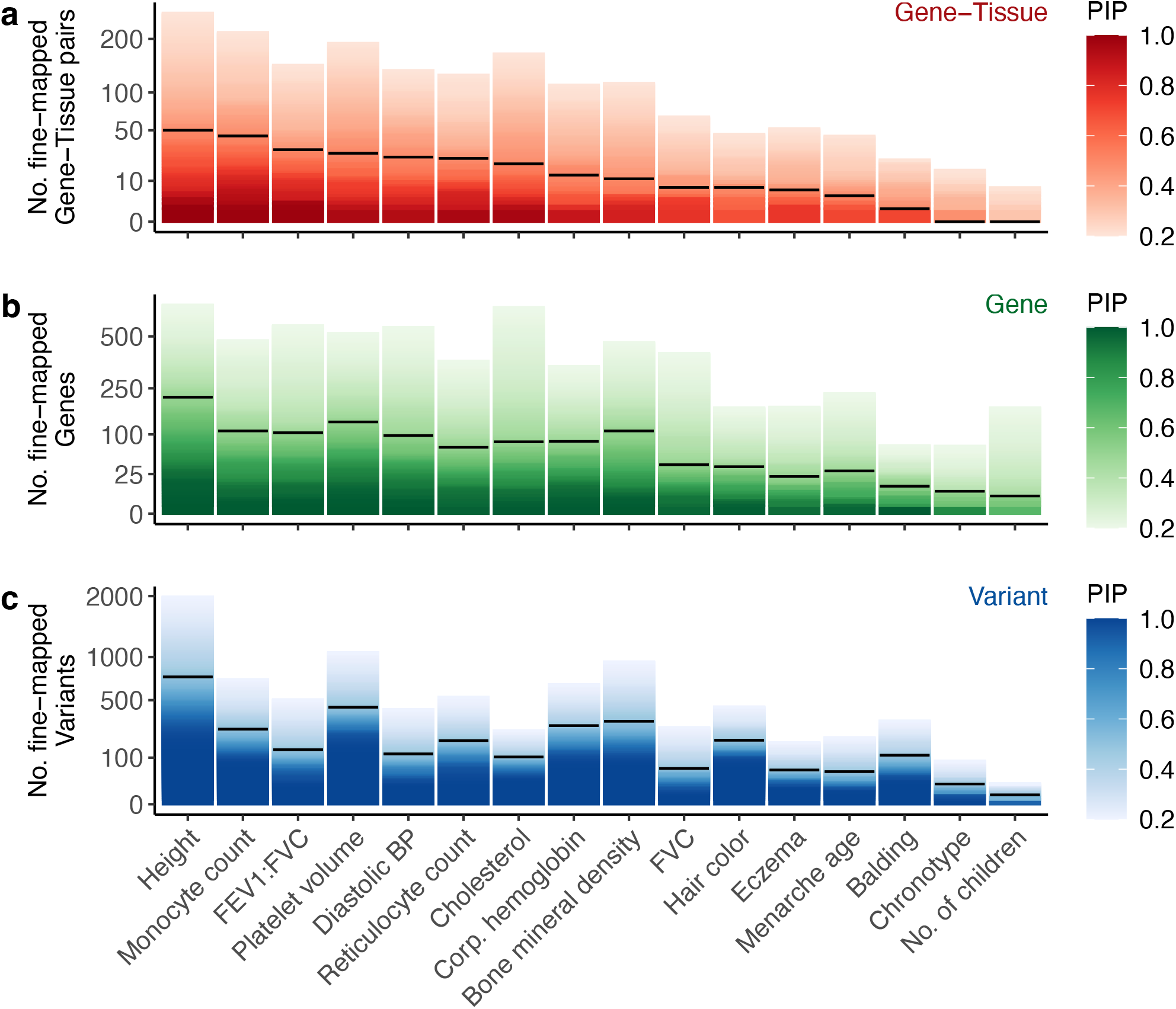
Summary results of fine-mapping genetic elements with TGFM for 16 independent UK Biobank diseases and traits. We report the number of **(a)** Gene-tissue pairs, **(b)** Genes, and **(c)** (non-mediated) Variants fine-mapped using TGFM (y-axis; square root scale) across 16 independent UK Biobank traits (x-axis) at various PIP thresholds ranging from 0.2 to 1.0 (color-bars). Horizontal black lines denote the number of genetic elements fine-mapped at PIP=0.5. FEV1:FVC, ratio of forced expiratory volume in 1 second to forced vital capacity; Platelet volume, Mean platelet volume; Diastolic BP, Diastolic blood pressure; Reticulocyte count, High-light scatter reticulocyte count; Corp. hemoglobin, Mean corpuscular hemoglobin; FVC, Forced vital capacity. Results for all 45 UK Biobank diseases and traits are reported in Supplementary Figure 30, and numerical results are reported in Supplementary Table 8.

For each trait, we identified the most frequently implicated tissues, computing the proportion of fine-mapped gene-tissue pairs in each tissue by counting gene-tissue pairs with PIP > 0.5 (Methods). Results are reported in Figure 4a (14 representative traits), Supplementary Figure 31 (all 45 traits), and Supplementary Table 9. Tissue-trait pairs that were frequently implicated by fine-mapped gene-tissue pairs were concentrated in expected trait-critical tissues (see below). Results were similar at other PIP thresholds (Supplementary Figure 32). We caution that these results may be impacted by limited power at low eQTL sample sizes (see Discussion).

**Figure 4:**
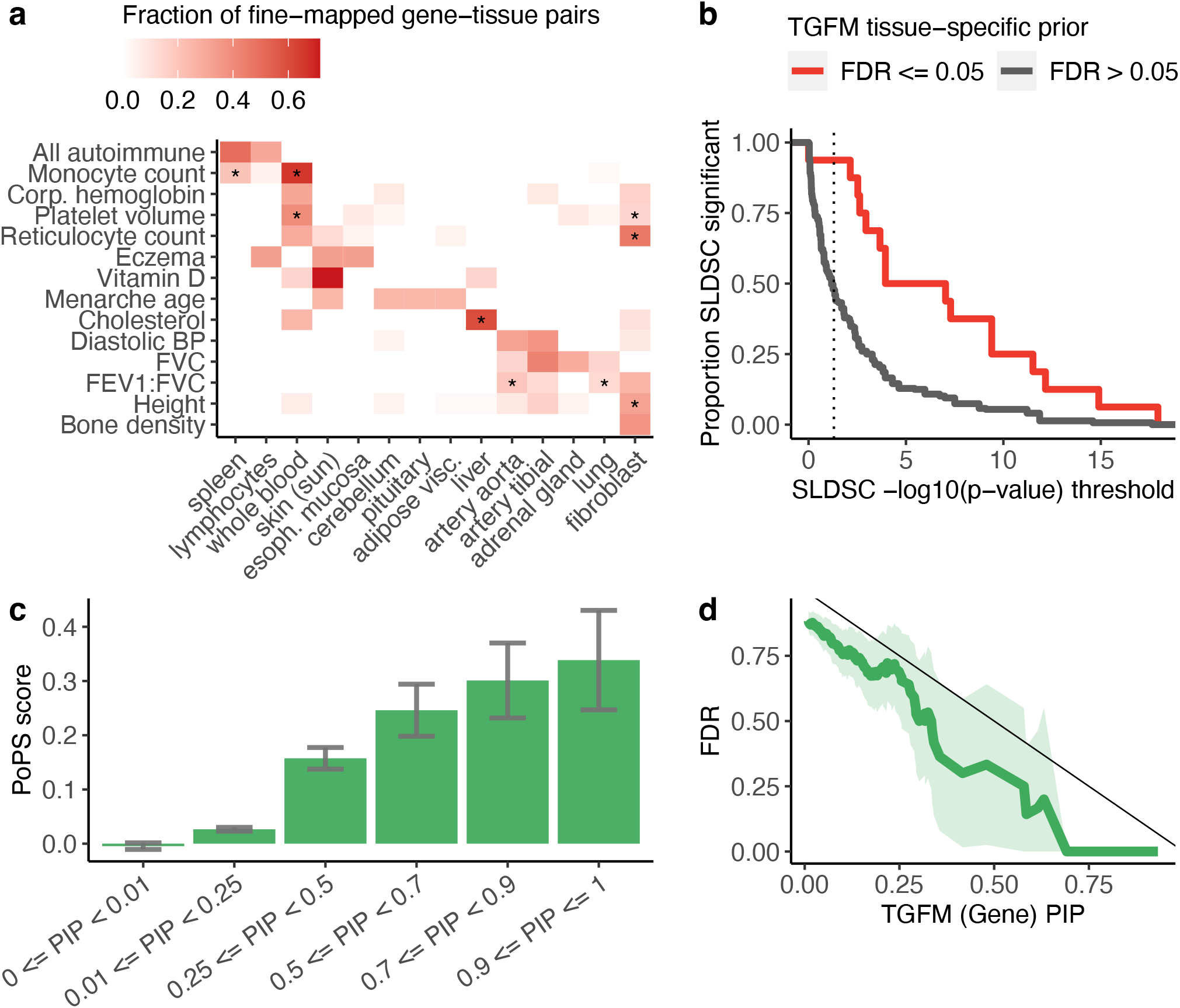
Properties of fine-mapped tissues and genes. **(a)** Proportion of fine-mapped gene-tissue pairs in each tissue (x-axis) for 14 representative traits (y-axis). Proportions for each trait were calculated by counting the number of gene-tissue pairs with TGFM PIP > 0.5 in each tissue and normalizing the counts across tissues. Tissues are only displayed if their proportion is > 0.2 for at least one of the 14 representative traits. Asterisks denote statistical significance (FDR ≤ 0.05 via the TGFM tissue-specific prior) of each tissue-trait pair. Results for all remaining traits and tissues are reported in Supplementary Figure 31, and numerical results are reported in Supplementary Table 9. The 14 representative traits were selected by including 12 of the 16 independent traits (Figure 3) with many high PIP gene-tissue pairs and two additional, interesting traits (All autoimmune and Vitamin D level). **(b)** Proportion of stratified tissue-trait pairs reported as statistically significant in S-LDSC analyses using chromatin data (y-axis) as a function of S-LDSC significance thresholds (x-axis), across all 45 traits analyzed; tissue-trait pairs are stratified according to significance (FDR ≤ 0.05 or FDR > 0.05) via the TGFM tissue-specific prior. Results at alternative TGFM tissue-specific prior significance thresholds are reported in Supplementary Figure 33, and numerical results are reported in Supplementary Table 11. **(c)** Average PoPS score (y-axis) of genes stratified by TGFM (Gene) PIP (x-axis). Averages were computed across genes for the 16 independent traits listed in Figure 3, as both PoPS score and TGFM gene PIPs are trait-specific. Error bars denote 95% confidence intervals. Numerical results are reported in Supplementary Table 13. **(d)** Empirical FDR when distinguishing a silver-standard gene set of 69 known LDL cholesterol genes analyzed in Figure 4 of ref. ^27^ from nearby genes (y-axis), for TGFM (Gene) PIP for LDL cholesterol greater than or equal to a range of PIP thresholds (x-axis). Light green shading denotes 95% confidence intervals. Numerical results are reported in Supplementary Table 15.

**Figure 5:**
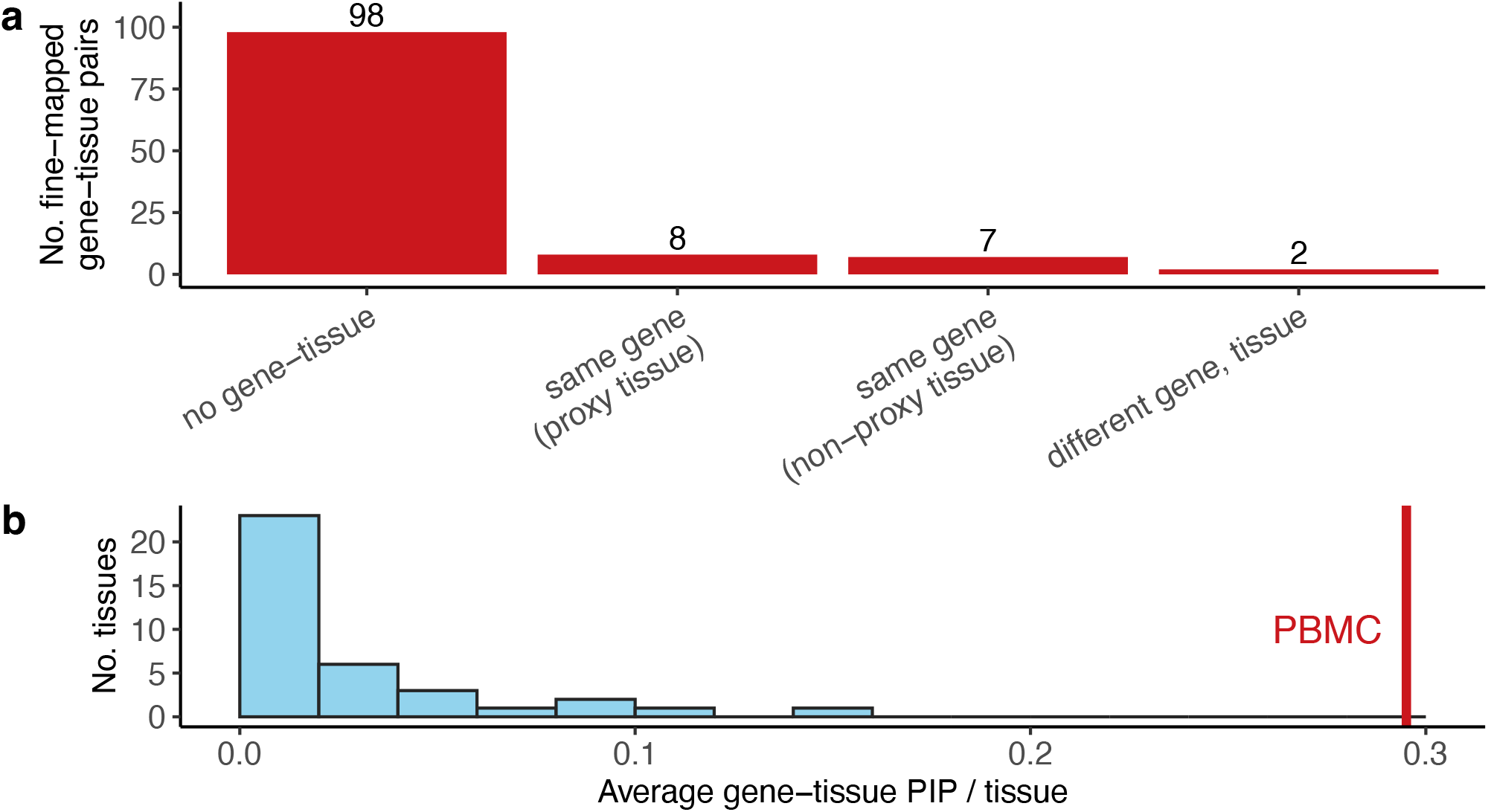
Robustness of TGFM results in analyses of alternative eQTL data sets. **(a)** Results of tissue-ablation analysis of 115 gene-tissue pairs for 18 representative traits that were prioritized by TGFM (PIP > 0.5) in the primary analysis. We report the number of loci in the tissue-ablation analysis with no gene-tissue pair prioritized by TGFM (PIP > 0.5); a gene-tissue pair prioritized by TGFM (PIP > 0.5) corresponding to the same gene and the best proxy tissue (see Methods); a gene-tissue pair prioritized by TGFM (PIP > 0.5) corresponding to the same gene and a non-proxy tissue; or a gene-tissue pair prioritized by TGFM (PIP > 0.5) corresponding to a different gene. Results at alternative PIP thresholds are reported in Supplementary Figure 38, and numerical results are reported in Supplementary Table 16. **(b)** Results of replacing GTEx whole blood (*N*=320) with pseudobulk PBMC (*N*=113) for 62 gene-trait pairs for 18 representative traits that TGFM fine-mapped for GTEx whole blood (PIP > 0.5) in the primary analysis. The vertical red line denotes the average PIP in PBMC, and the histogram summarizes the average PIP of each GTEx tissue (excluding whole blood). Numerical results are reported in Supplementary Table 17. The 18 representative traits consist of the 16 independent traits (Figure 3) and two additional, interesting traits (All autoimmune and Vitamin D levels).

Separately, we assessed the statistical significance of implicated tissue-trait pairs by applying genomic bootstrapping to the TGFM tissue-specific prior. Results are reported in Figure 4a (14 representative traits), Supplementary Figure 31 (all 45 traits), and Supplementary Table 9. This approach identified 23 tissue-trait pairs with FDR ≤ 0.05 (64 tissue-trait pairs with FDR ≤ 0.2). Despite limited power, the TGFM tissue-specific prior identified 6 traits with more than one significantly associated tissue (FDR ≤ 0.05; 17 traits at FDR ≤ 0.2); this result motivates the use of TGFM over a two-step approach of separately identifying the causal gene using a gene-level fine-mapping method^24^ and identifying the causal tissue using a method for identifying trait-critical tissues^5,7,11^.

Although inference of trait-critical tissues is not a primary goal of TGFM (and there exist previous methods for inferring trait-critical tissues using eQTL data^5,11^), these results validate the TGFM tissue-specific prior.

We highlight 4 diseases/traits whose TGFM-implicated tissues recapitulate known biology or nominate biologically plausible hypotheses. First, 60% of Total cholesterol fine-mapped gene-tissue pairs involved liver tissue (FDR ≤ 0.05 for tissue-specific prior); the involvement of liver tissue in Total cholesterol is well-established^48^. Second, 36% and 32% of the Diastolic blood pressure fine-mapped gene-tissue pairs involved artery tibial and artery aorta tissue, respectively (FDR ≤ 0.2 for tissue-specific prior in both tissues); a critical role for artery in Diastolic blood pressure is a biologically plausible hypothesis recapitulated by previous studies^7,11^ and consistent with Diastolic blood pressure measuring the pressure on the aorta^49^. Third, 50% and 30% of the All autoimmune disease fine-mapped gene-tissues pairs involved spleen tissue and lymphocytes respectively (FDR > 0.2 for tissue-specific prior in both tissues); spleen is a biologically plausible tissue^50,51^ (despite being non-significant via the tissue-specific prior), as spleen produces lymphocytes and the role of lymphocytes in autoimmune disease is well-established^52,53^. Fourth, 33%, 33% and 33% of the Eczema fine-mapped gene-tissue pairs involved skin (sun exposed) tissue, lymphocytes, and esophagus mucosa, respectively (FDR > 0.2 for tissue-specific prior in all tissues); both skin (sun exposed) and lymphocytes are biologically plausible tissues (despite being non-significant via the tissue-specific prior), as Eczema is an autoimmune disease mediated by T cells (a lymphocyte cell type)^54,55^ that manifests in skin tissue^55,56^; we also note that the esophagus-eczema pair is corroborated by ref. ^7^, which determined that digestive tissue and cell types were disease critical tissues for eczema in genome-wide heritability analyses of specifically expressed genes. We caution that these results may be impacted by limited power at low eQTL sample sizes (see Discussion).

We evaluated whether the tissue-trait pairs identified as statistically significant (via the TGFM tissue-specific prior) were also identified as statistically significant in S-LDSC analyses using chromatin data from a matched cell type group^3^ (see Methods; Supplementary Table 10). Results are reported in Figure 4b and Supplementary Table 11. Notably 94% of the tissue-trait pairs prioritized by the TGFM tissue-specific prior (16 tissue-trait pairs at FDR < 0.05) were nominally statistically significant (p < 0.05) in S-LDSC analyses using chromatin data. Results were similar at other FDR thresholds (Supplementary Figure 33).

We observed instances where TGFM was unable to distinguish the causal tissue within a small set of highly correlated tissues. For example, for waist-hip-ratio adjusted for BMI (WHRadjBMI), TGFM fine-mapped only 1 gene-tissue pair in adipose visceral and 0 gene-tissue pairs in adipose subcutaneous tissue at PIP > 0.5, despite strong prior evidence of the role of adipose tissue in WHRadjBMI^7,57^; however, TGFM fine-mapped 10 gene-tissue pairs in adipose (defined as adipose subcutaneous U adipose visceral) at PIP > 0.5 when summing PIPs of gene-tissue pairs across the two tissues (Methods). Unsurprisingly, the average correlation in cis-predicted gene expression of adipose subcutaneous vs. adipose visceral across all genes included in TGFM was very large (0.92). Average correlations for all pairs of tissues are reported in Supplementary Figure 34 and Supplementary Table 12; the average correlation ranged from 0.48 (whole blood and brain cerebellum) to 0.96 (brain substantia nigra and brain spinal cord), and the correlation patterns reflected known relationships between tissues.

### Validation of fine-mapped gene-tissue pairs

We sought to validate the gene-tissue pairs prioritized by TGFM. First, we assessed the overlap of genes prioritized by TGFM with genes prioritized by independent gene sets/scores. We assessed overlap with PoPS^58^, a similarity-based gene score that prioritizes trait-relevant genes from gene-level features such as cell-type-specific expression. Results are reported in Figure 4c and Supplementary Table 13. The average PoPS score increased as a function of TGFM (Gene) PIPs in different bins, from −0.0046 (s.e. 0.0031) for trait-gene pairs with PIP < 0.01 to 0.34 (s.e. 0.047) for trait-gene pairs with PIP ≥ 0.9; this provides an external validation of genes prioritized by TGFM. In addition, we assessed overlap with 10 non-disease-specific gene sets (e.g. High-pLI genes^59^) that are known to be enriched/depleted for disease heritability (from Figure 4 of ref. ^26^). Results are reported in Supplementary Figure 35a and Supplementary Table 14. Genes with TGFM (Gene) PIP > 0.5 were significantly (FDR < 0.05) overrepresented in Epigenetic modifier genes^60^ (odds ratio: 1.78), High-pLI genes^59^ (odds ratio: 1.31) and Mouse Genome Informatics (MGI) essential genes^61^ (odds ratio: 1.31) and underrepresented in genes with the most SNPs within 100kb (odds ratio: 0.48), consistent with previous findings^26^. Results were similar at other PIP thresholds (Supplementary Figure 35b).

Second, we empirically evaluated the calibration of TGFM using a silver-standard gene set of 69 known LDL cholesterol genes (analyzed in Figure 4 of ref. ^27^); following ref. ^27^, we used the 69 known LDL cholesterol genes as the positive set, and nearby genes as the negative set. Results are reported in Figure 4d and Supplementary Table 15. The empirical FDR of TGFM (Gene) with respect to the silver-standard gene set was lower than (1 – TGFM (Gene) PIP threshold), implying that TGFM is well-calibrated. Indeed, the empirical FDR tracked with (1 – average TGFM (Gene) PIP), a less conservative choice than (1 – TGFM (Gene) PIP threshold), indicating precise calibration (Supplementary Figure 36). We obtained a similar result when empirically evaluating the calibration of TGFM (Gene-Tissue) PIPs using the same silver-standard gene set in conjunction with liver tissue (Supplementary Figure 37). We also compared the calibration and power of TGFM and cTWAS (applied to GTEx liver tissue)^27^ using the 69 LDL cholesterol genes. Results are reported in Supplementary Figure 36. Unlike TGFM (Gene), the empirical FDR for cTWAS was significantly larger than (1 – average PIP), contrary to perfect calibration. In comparisons at the same level of FDR, TGFM (Gene) attained higher power than cTWAS, but differences were not statistically significant.

Third, we evaluated how TGFM performs in the setting of a missing causal tissue. We reran TGFM on 18 representative diseases/traits while removing (ablating) the most significant tissue for each trait (based on TGFM tissue-specific priors in the primary analysis; Supplementary Figure 31). We considered 115 gene-tissue pairs for which the ablated tissue was prioritized by TGFM (PIP > 0.5) in the primary analysis. Results are reported in Figure 5a and Supplementary Table 16. 98/115 loci had no gene-tissue pair prioritized by TGFM (PIP > 0.5) and 8/115 had only the gene-tissue pair consisting of the same gene and the best proxy tissue (see Methods) prioritized by TGFM. This suggests that when the causal tissue is missing, TGFM will tend to prioritize nothing, or prioritize the best proxy tissue, instead of prioritizing an unrelated tissue. Results were similar at other PIP thresholds (Supplementary Figures 38, 39).

Fourth, we assessed the robustness of TGFM results using independent eQTL data. Specifically, we reran TGFM on 18 representative diseases/traits using pseudobulk peripheral blood mononuclear cell (PBMC) eQTL data^33^ (*N*=113; see Methods) in place of GTEx whole blood (*N*=320) (while retaining other GTEx tissues). We considered 62 gene-trait pairs that TGFM fine-mapped for GTEx whole blood (PIP > 0.5) in the primary analysis. Results are reported in Figure 5b and Supplementary Table 17. PBMC had an average PIP of 0.29 across all genes containing a gene model in PBMC, much larger than any other tissue (average PIP of 0.00-0.15). This indicates that when replacing an implicated gene-tissue pair with a different eQTL data set, TGFM tends to prioritize the same gene-tissue pair. In addition, we performed a similar analysis using GTEx whole blood down-sampled to the PBMC sample size of N=113 (down-sampled from N=320) instead of PBMC. Remarkably, replacing GTEx whole blood with PBMC in a different eQTL data set (*N*=113) performed as well as down-sampling GTEx whole blood to *N*=113 in the same eQTL data set (average PIP of 0.28 vs. 0.00-0.15 for other tissues; Supplementary Figure 40). This indicates that TGFM results are robust across different eQTL data sets.

### TGFM pinpoints disease genes and tissues of action

We highlight 6 examples of fine-mapped (PIP > 0.5) gene-tissue-trait triplets that recapitulate known biology or nominate biologically plausible mechanisms. First, TGFM fine-mapped *TPO* (*Thyroid Peroxidase*) in thyroid for Hypothyroidism (Figure 6a; gene-tissue PIP: 0.88; gene PIP: 0.88). *TPO* is an enzyme involved in thyroid hormone biosynthesis and its involvement in Hypothyroidism has been well-studied^62,63^, and TPO has been linked to Hypothyroidism in genetic association studies^64^. Thyroid was also identified as a Hypothyroidism-critical tissue genome-wide (proportion of fine-mapped gene-tissue pairs = 0.21, bootstrap p = 0.04 for tissue-specific prior; Supplementary Figure 31).

**Figure 6:**
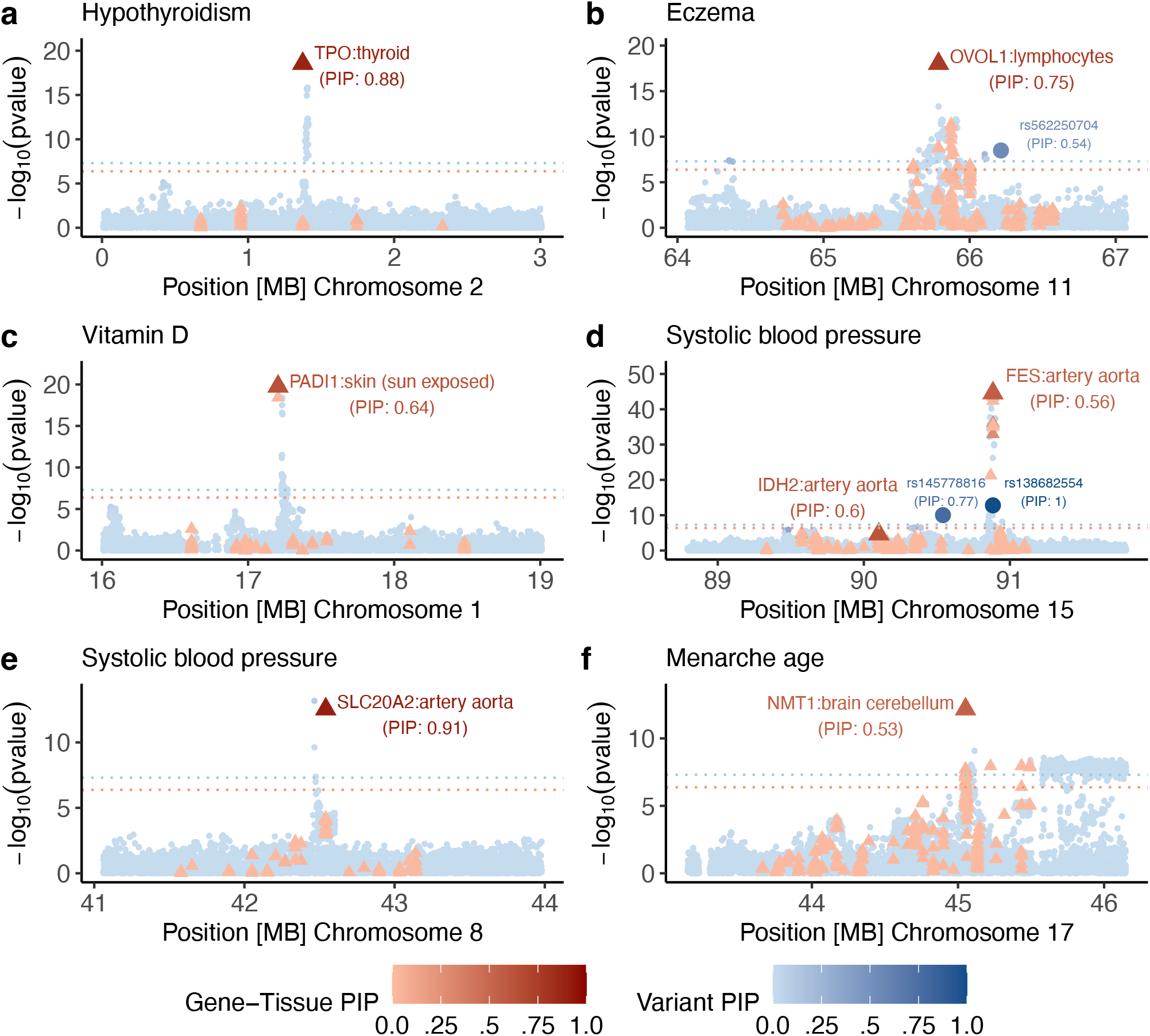
Examples of fine-mapped gene-tissue-disease triplets identified by TGFM. We report 6 example loci for which TGFM fine-mapped a gene-tissue pair (PIP > 0.5). In each example we report the marginal GWAS and TWAS association -log_10_ p-values (y-axis) of non-mediated variants (blue circles) and gene-tissue pairs (red triangles). Marginal TWAS association -log_10_ p-values were calculated by taking the median -log_10_ TWAS p-value across the 100 sets of sampled cis-predicted expression models for each gene-tissue pair. The genomic position of each gene-tissue pair (x-axis) was based on the gene’s TSS. The color shading of each variant and gene-tissue pair was determined by its TGFM PIP. Any genetic element with TGFM PIP > 0.5 was made larger in size. Dashed horizontal blue and red lines represent GWAS significance (5 × 10^−8^) and TWAS significance (4.2 × 10^−7^) thresholds, respectively. Numerical results are reported in Supplementary Table 18.

**Figure 7:**
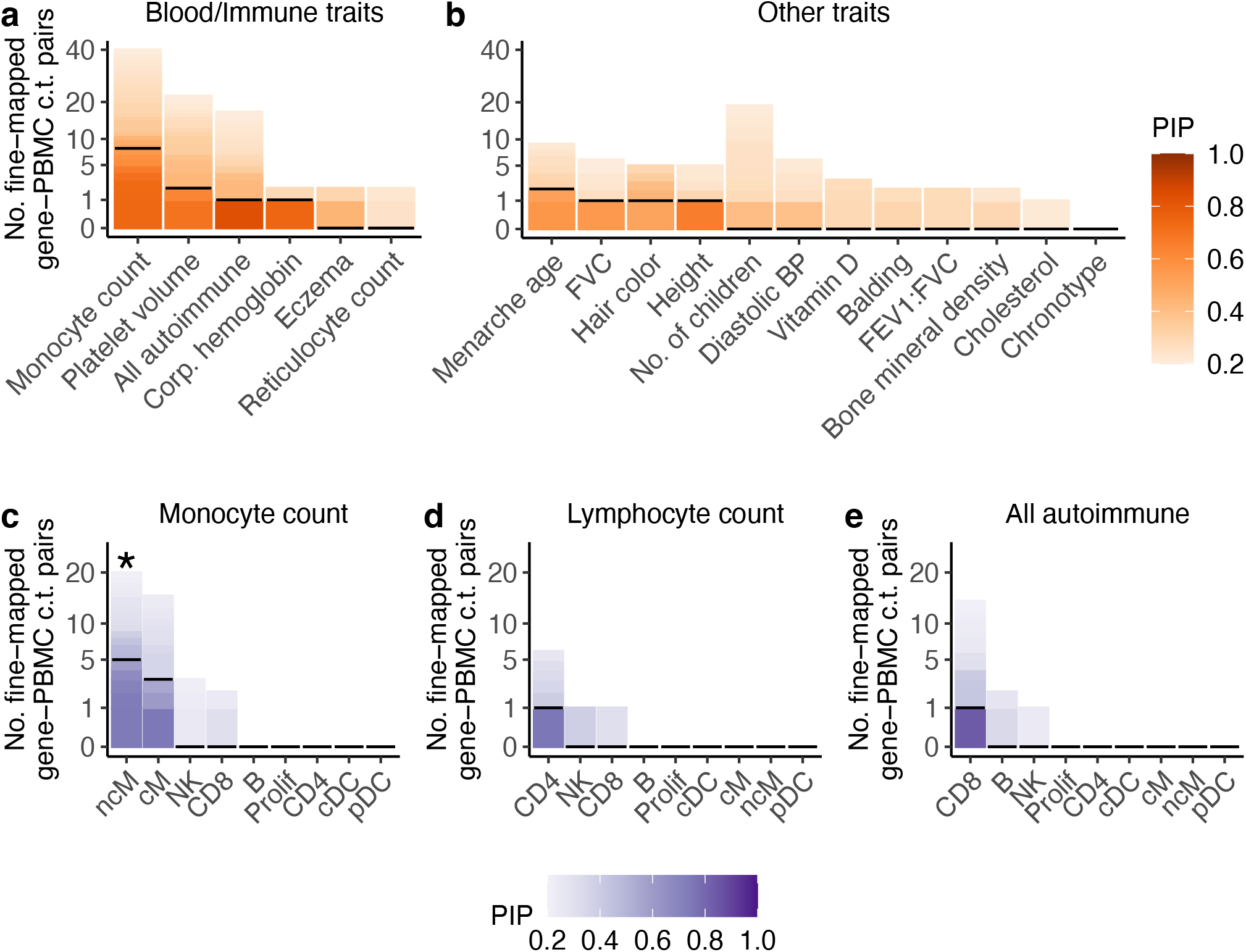
Summary results of fine-mapping gene-PBMC cell type pairs with TGFM for 18 representative UK Biobank diseases and traits. **(a-b)** Number of gene-PBMC cell type pairs fine-mapped using TGFM (y-axis; square root scale) across 18 representative UK Biobank traits (x-axis) at various PIP thresholds ranging from 0.2 to 1.0 (color-bar), distinguishing between **(a)** autoimmune diseases and blood cell traits and **(b)** non-blood-related traits. Horizontal black lines denote the number of gene-PBMC cell type pairs fine-mapped at PIP=0.5. The 18 representative traits (same as Figure 5) consist of the 16 independent traits (Figure 3) and two additional, interesting traits (All autoimmune and Vitamin D levels). Results for all 45 UK Biobank diseases and traits are reported in Supplementary Figure 42. **(c-e)** Number of gene-PBMC cell type pairs fine-mapped using TGFM (y-axis; square root scale) in each of the 9 PBMC cell types (x-axis) at various PIP thresholds ranging from 0.2 to 1.0 (color-bar) for **(c)** Monocyte count, **(d)** Lymphocyte count, and **(e)** All autoimmune disease. Horizontal black lines denote the number of gene-PBMC cell type pairs fine-mapped at PIP=0.5. Asterisks denote statistical significance (FDR ≤ 0.05 via the TGFM tissue-specific prior) of each PBMC cell type-trait pair. Results for all 45 UK Biobank diseases and traits are reported in Supplementary Figure 43. Numerical results are reported in Supplementary Table 20.

Second, TGFM fine-mapped *OVOL1* in lymphocytes for Eczema (Figure 6b, gene-tissue PIP: 0.75, gene PIP: 0.76). Recent work demonstrated that loss of *OVOL1* results in skin inflammation and Eczema via an immune-mediated mechanism in T cells (a lymphocyte cell type)^65–67^, and *OVOL1* has previously been linked to Eczema in genetic association studies^68^. Lymphocyte was suggestively implicated as an Eczema-critical tissue genome-wide (proportion of fine-mapped gene-tissue pairs = 0.36, bootstrap p = 0.08 for tissue-specific prior; Supplementary Figure 31). There exist 28 other gene-tissue pairs within 1 Mb of the TSS of *OVOL1* (4 of which correspond to *OVOL1* in a tissue other than lymphocytes) that had significant TWAS p-values (p ≤ 0.05 / 119,270 = 4.2 × 10^−7^) but were not fine-mapped by TGFM (all with PIP ≤ 0.01), underscoring the benefit of joint fine-mapping of gene-tissue pairs. TGFM also fine-mapped one non-mediated variant (rs56225074; PIP: 0.55) within 1 Mb of the TSS of *OVOL1*, perhaps due to finite eQTL sample size and/or absence of the causal cell-type or context in GTEx expression data^28,69^ (see Discussion).

Third, TGFM fine-mapped *PADI1* in skin (sun exposed) for Vitamin D level (Figure 6c; gene-tissue PIP: 0.64, gene PIP: 0.65). TGFM assigned *PADI1* a PIP of 0.64 in skin (sun exposed) tissue (TWAS p = 1.7 × 10^−20^) and a PIP of 0.01 in skin (not sun exposed) tissue (TWAS p = 3.8 × 10^−19^), demonstrating TGFM’s ability to distinguish trait-critical tissues from closely related tissues. *PADI1* is known to interact with keratins during epidermal differentation^70,71^, and has previously been linked to Vitamin D level in genetic association studies^72,73^. Skin (sun exposed) was also identified as a Vitamin D level-critical tissue genome-wide (proportion of fine-mapped gene-tissue pairs = 0.72, bootstrap p = 0.008 for tissue-specific prior; Supplementary Figure 31).

Fourth, TGFM fine-mapped both *IDH2* in artery aorta and *FES* in artery aorta at the same locus for Systolic blood pressure (Figure 6d, *IDH2* gene-tissue PIP: 0.60, *IDH2* gene PIP: 0.94, *FES* gene-tissue PIP: 0.56, *FES* gene PIP: 0.96); we note that *IDH2* in artery aorta and *FES* in artery aorta are independent genetic elements, with *r*^2^ < 0.001 in their cis-predicted expression. Previous work has experimentally demonstrated that IDH2 deficiency aggravates hypertension and cardiac hypertrophy via IDH2 knockout experiments in mice^74,75^, and that FES inhibits atherosclerosis via CRISPR experiments in cell lines^76^; both genes have been linked to hypertension in genetic association studies^77,78^. Artery aorta was also identified as a Systolic blood pressure-critical tissue genome-wide (proportion of fine-mapped gene-tissue pairs = 0.36, bootstrap p = 0.0008 for tissue-specific prior; Supplementary Figure 31). The result of TGFM fine-mapping two experimentally validated genes at this locus underscores the advantage of TGFM over simply selecting the gene-tissue pair with the most significant TWAS association at a locus. TGFM also fine-mapped two non-mediated variants (rs145778816 and rs138682554; PIP: 0.70 and PIP: 1.0, respectively) within 1 Mb of the TSS of *IDH2* and *FES*, perhaps due to finite eQTL sample size and/or absence of the causal cell-type or context in GTEx expression data^28,69^ (see Discussion).

Fifth, TGFM fine-mapped *SLC20A2* in artery aorta for Systolic blood pressure (Figure 6e, gene-tissue PIP: 0.91, gene PIP: 0.91). Previous work has shown that loss of *SLC20A2* results in Human idiopathic basal ganglia calcification^79,80^ and well as arteriolar calcification^81^, but *SLC20A2* has not previously been linked to Systolic blood pressure to our knowledge. Artery aorta was also identified as a Systolic blood pressure-critical tissue genome-wide (proportion of fine-mapped gene-tissue pairs = 0.36, bootstrap p = 0.0008 for tissue-specific prior; Supplementary Figure 31).

Sixth, TGFM fine-mapped *NMT1* in brain cerebellum for Menarche age (Figure 6f, gene-tissue PIP: 0.53, gene PIP: 0.86); TGFM also assigned *NMT1* in brain limbic a PIP of 0.10. Recent work demonstrated that *NMT1* cis-predicted expression in brain tissues was marginally associated with image-derived brain phenotypes^82,83^, but *NMT1* has not previously been linked to Menarche age to our knowledge. Brain cerebellum was also identified as a Menarche age-critical tissue genome-wide (proportion of fine-mapped gene-tissue pairs = 0.25, bootstrap p = 0.05 for tissue-specific prior; Supplementary Figure 31), consistent with previous studies^3,84^. There exist 20 other gene-tissue pairs within 1 Mb of the TSS of *NMT1* (11 of which correspond to *NMT1* in a tissue other than brain cerebellum) that had significant TWAS p-values (p ≤ 0.05 / 119,270 = 4.2 × 10^−7^) but were not fine-mapped by TGFM (all with PIP ≤ 0.01), underscoring the benefit of joint fine-mapping of gene-tissue pairs.

Additional examples are discussed in the Supplementary Note and Supplementary Figure 41. These examples include additional implicated tissues for Vitamin D level (liver, in addition to skin (sun exposed))^72,85^ and Systolic blood pressure (brain cerebellum, in addition to artery aorta)^86,87^ that are consistent with known biology, highlighting the advantages of TGFM over a two-step fine-mapping approach that first infers the causal tissue^5,7,11^ and then performs traditional gene-level fine-mapping^24,27^ in the inferred causal tissue.

### TGFM pinpoints disease genes and fine-grained cell-types in single-cell eQTL data

It is widely hypothesized that eQTL in fine-grained cell types/contexts may help resolve the limited proportion of disease heritability explained by eQTL in bulk tissues^28,69^.

Accordingly, we applied TGFM to fine-map disease genes and fine-grained cell types in 45 disease and complex traits from the UK Biobank (same diseases/traits as above) using a recently generated single-cell eQTL data set spanning 9 fine-grained peripheral blood mononuclear cell (PBMC) cell types^33^ (average *N*=112; Methods and Supplementary Table 19). We converted single-cell expression measurements in each cell type to pseudobulk expression (see Methods), and also included the 38 GTEx tissues analyzed above. For each disease/trait, we applied TGFM to 2,682 overlapping 3-Mb loci spanning 1,851 (protein-coding) gene-PBMC cell type pairs with cis-predicted expression models (206 genes per cell type on average, Supplementary Table 19; 1,066 unique genes), 119,270 (protein-coding) gene-tissue pairs with cis-predicted expression models (GTEx tissues; 3,139 genes per tissue on average, Supplementary Table 6; 13,700 unique genes) and 10,545,304 genetic variants with MAF ≥ 0.005. We assigned a PIP to each gene-PBMC cell type pair, gene-tissue pair, gene, and non-mediated genetic variant, analogous to above. We have publicly released PIPs for all gene-PBMC cell type pairs, gene-tissue pairs, genes, and non-mediated variants for each disease/trait (see Data Availability).

Results are reported in Figure 7a-b (18 representative traits), Supplementary Figure 42 (all 45 traits), and Supplementary Table 20. Across all 45 traits, TGFM identified 30 gene-PBMC cell type-trait triplets at PIP > 0.5; TGFM was not sufficiently powered to detect any gene-PBMC cell type-trait triplets at PIP > 0.9, likely due to the limited single-cell eQTL sample size. Of the 30 gene-PBMC cell type-trait triplets with PIP > 0.5, 25 involved a trait locus that had no confidently fine-mapped gene-tissue pair (no TGFM PIP > 0.5 in GTEx-only analysis corresponding to Figures 3-6). For the 18 representative traits, TGFM identified 12 gene-PBMC cell type pairs at PIP > 0.5 for autoimmune diseases and blood cell traits (Figure 7a) vs. 5 gene-PBMC cell type pairs at PIP > 0.5 for non-blood-related traits (Figure 7b; includes 2 gene-PBMC cell type pairs for Menarche age, which we conservatively labeled as non-blood-related even though it has been reported to be partially immune-mediated^88,89^), increasing to 23 vs. 7 for all 45 traits; this validates the importance of gene expression in blood cell types for blood-related traits.

**Figure 8:**
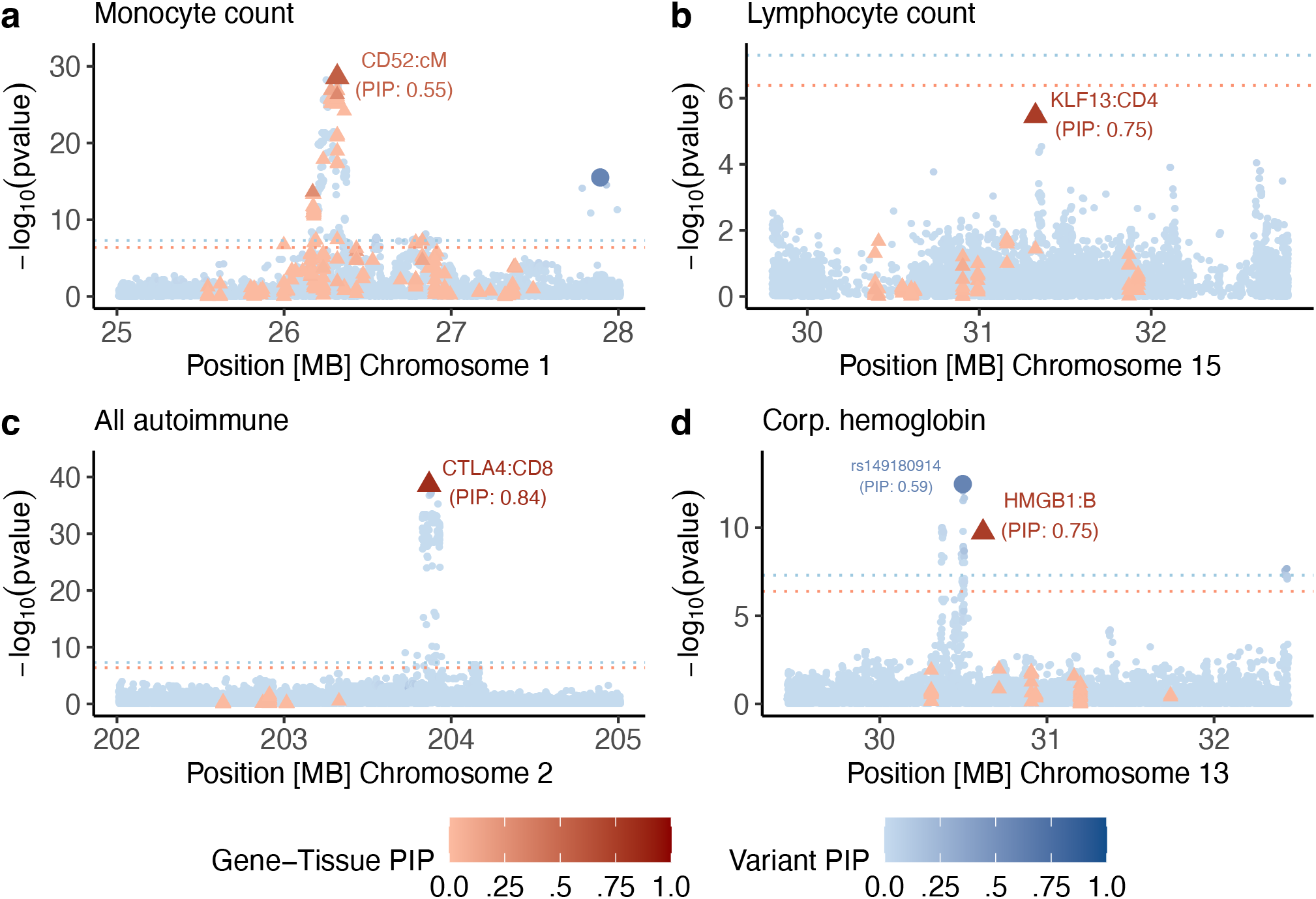
Examples of fine-mapped gene-PBMC cell type-disease triplets identified by TGFM. We report 4 example loci for which TGFM fine-maps a gene-PBMC cell type pair (PIP > 0.5). In each example we report the marginal GWAS and TWAS association -log_10_ p-values (y-axis) of non-mediated variants (blue circles) and gene-tissue (or gene-PBMC cell type) pairs (red triangles). Marginal TWAS association - log_10_ p-values were calculated by taking the median -log_10_ TWAS p-value across the 100 sets of sampled cis-predicted expression models for each gene-tissue (or gene-PBMC cell type) pair. The genomic position of each gene-tissue (or gene-PBMC cell type) pair (x-axis) was based on the gene’s TSS. The color shading of each variant and gene-tissue (or gene-PBMC cell type) pair was determined by its TGFM PIP. Any genetic element with TGFM PIP > 0.5 was made larger in size. Dashed horizontal blue and red lines represent GWAS significance (5 × 10^−8^) and TWAS significance (4.1 × 10^−7^) thresholds, respectively. Numerical results are reported in Supplementary Table 22.

For each trait, we identified the most frequently implicated PBMC cell types, computing the proportion of fine-mapped gene-PBMC cell type pairs in each PBMC cell type by counting gene-PBMC cell type pairs with PIP > 0.5. Results are reported in Figure 7c-e (3 representative blood-related traits), Supplementary Figure 43 (all 45 traits), and Supplementary Table 20. Despite low power, the fine-mapped gene-PBMC cell type pairs at PIP > 0.5 were concentrated in expected trait-critical PBMC cell types, e.g. 62.5% in non-classical monocyte (ncM) cells and 37.5% in classical monocyte (cM) cells for Monocyte count, 100% in CD4^+^ T cells for Lymphocyte count, and 100% in CD8^+^ T cells for All autoimmune disease^7,90,91^. Results were similar at other PIP thresholds (Supplementary Figure 43). Despite published evidence of the biological role of both CD4^+^ T cells^92,93^ and CD8^+^ T^94–96^ cells in autoimmune disease, TGFM exclusively fine-mapped gene-PBMC cell type pairs involving CD8+ T cells for All autoimmune disease; we attribute this incomplete finding to the limited power of TGFM at low eQTL sample sizes (see Discussion).

Separately, we assessed the statistical significance of implicated PBMC cell type-trait pairs. Results are reported in Figure 7c-e (3 representative blood-related traits), Supplementary Figure 43 (all 45 traits), and Supplementary Table 21. Non-classical monocyte (ncM) cells-Monocyte count was the only PBMC cell type-trait pair significant at FDR ≤ 0.05, a finding that is both expected and corroborated by previous work^10^.

We highlight 4 examples of fine-mapped (PIP > 0.5) gene-PBMC cell type-trait triplets that recapitulate known biology or nominate biologically plausible mechanisms. First, TGFM fine-mapped *CD52* in classical monocyte (cM) cells for Monocyte count (Figure 8a; gene-PBMC cell type PIP: 0.55; gene PIP: 0.88; gene-tissue PIP ≤ 0.05 in all GTEx tissues); TGFM also assigned non-negligible PIPs to *CD52* in multiple correlated PBMC cell types and GTEx tissues including 0.29 in non-classical monocyte (ncM) cells and 0.05 in GTEx whole blood. Previous work demonstrated that *CD52* regulates immune homeostasis in monocytes and T cells by inhibiting NF-*κ*B signalling^97–99^, but *CD52* has not previously been linked to Monocyte count to our knowledge. cM cells were also identified as a Monocyte count-critical cell type genome-wide (bootstrap p = 0.04 for tissue-specific prior; Supplementary Table 21). There exist 56 other gene-tissue pairs and 3 other gene-PBMC cell type pairs within 1 Mb of the TSS of *CD52* (14 and 2 of which correspond to *CD52* in a cell type or tissue other than cM, respectively) with significant TWAS p-values (p ≤ 0.05 / 121,121 = 4.1 × 10^−7^, where 121,121 = 119,270 + 1,851) but not fine-mapped by TGFM (all with PIP ≤ 0.01), underscoring the benefit of joint fine-mapping of gene-tissue and gene-PBMC cell type pairs. *CD52* was not prioritized in any tissue in the GTEx-only analysis of Figure 3 (highest gene-tissue PIP = 0.07 (whole blood)), underscoring the advantages of modeling gene expression in fine-grained PBMC cell types.

Second, TGFM fine-mapped *KLF13* in CD4^+^ T (CD4) cells for Lymphocyte count (Figure 8b; gene PBMC cell type PIP: 0.75; gene PIP: 0.75; gene-tissue PIP ≤ 0.01 in all GTEx tissues) despite a non-significant TWAS p-value (p = 3.5 × 10^−6^ > 0.05 / 121,121 = 4.1 × 10^−7^). *KLF13* has been shown to regulate lymphocyte development and survival^100,101^, but *KLF13* has not previously been linked to Lymphocyte count in genetic association studies to our knowledge. CD4 cells were suggestively implicated as a Lymphocyte count-critical cell type genome-wide (bootstrap p = 0.13 for tissue-specific prior; Supplementary Table 21). *KLF13* was not prioritized in any tissue in the GTEx-only analysis of Figure 3 (highest gene-tissue PIP = 0.002 (brain cerebellum)), again underscoring the advantages of modeling gene expression in fine-grained PBMC cell types.

Third, TGFM fine-mapped *CTLA4* (*Cytotoxic T-lymphocyte associated protein 4*) in CD8^+^ T cells for All autoimmune disease (Figure 8c; gene-PBMC cell type PIP: 0.84; gene PIP: 0.85), also assigning a non-negligible PIP to *CTLA4* in CD4^+^ T cells (gene-PBMC cell type PIP: 0.02). *CTLA4* is a well-studied regulator of immune responses in CD4^+^ T cells^102–105^ (with some evidence for a role for *CTLA4* in CD8^+^ T cells in autoimmune disease^94,105–107^), and has previously been linked to autoimmune diseases (rheumatoid arthritis, systemic lupus erythematosus, and type 1 diabetes) in genetic association studies^108,109^. CD8^+^ T cells were also identified as an All autoimmune disease-critical cell type genome-wide (bootstrap p = 0.05 for tissue-specific prior; Supplementary Table 21). *CTLA4* did not meet the criteria for having a cis-predicted expression model in any GTEx tissue, underscoring the advantages of modeling gene expression in fine-grained PBMC cell types.

Fourth, TGFM fine-mapped *HMGB1* in B cells for Mean corpuscular hemoglobin (Figure 8d; gene-tissue PIP: 0.75; gene PIP: 0.75). *HMGB1* has previously been shown to mediate anemia of inflammation (i.e. anemia resulting from a prolonged immune response) in mice^110,111^, but *HMGB1* has not previously been linked to Mean corpuscular hemoglobin to our knowledge. B cells were also identified as a Mean corpuscular hemoglobin-critical cell type genome-wide (bootstrap p = 0.05 for tissue-specific prior; Supplementary Table 21). *HMGB1* did not meet the criteria for having a cis-predicted expression model in any GTEx tissue, again underscoring the advantages of modeling gene expression in fine-grained PBMC cell types. TGFM also fine-mapped one non-mediated variant (rs149180914; PIP: 0.59) within 1 Mb of the TSS of *HMGB1* (see Discussion).

## Discussion

We developed a new method, TGFM, that jointly fine-maps causal gene-tissue pairs and non-mediated genetic variants at disease-associated loci. We applied TGFM to 45 UK Biobank diseases and traits using 38 GTEx tissues and identified many causal gene-tissue pairs (PIP > 0.5), which were concentrated in known disease-critical tissues^2–11^ and strongly enriched in known disease-relevant genes^58,59^. Causal gene-tissue pairs identified by TGFM recapitulated known biology, but also included biologically plausible novel findings. We further applied TGFM to single-cell eQTL data from 9 cell types in PBMC (analyzed jointly with GTEx tissues) and identified additional causal gene-PBMC cell type pairs (PIP > 0.5), primarily for autoimmune disease and blood cell traits.

TGFM has three advantages over previous methods for fine-mapping causal genes. First, TGFM identifies causal gene-tissue pairs, not just causal genes. Second, TGFM jointly models the disease contribution of each gene-tissue pair and non-mediated variant, disentangling causal gene-tissue pairs from both tagging gene-tissue pairs and tagging non-mediated genetic variants. Third, TGFM employs a sampling procedure that accounts for uncertainty in cis-predicted expression models. Our simulations show that TGFM attains accurate calibration of fine-mapped gene-tissue pairs, in contrast to previous methods such as coloc^17^, FOCUS^24^, cTWAS^27^, JLIM^45^, and SMR^46^ (Figure 1 and Supplementary Figure 2), all of which lack some of these advantages (Table 1). The calibration of TGFM strongly outperforms previous methods even in the case of a locus with a single causal gene-tissue pair (Supplementary Figure 12), or in genome-wide data with a single causal tissue (Supplementary Figure 10) or a single causal tissue that is known (Supplementary Figure 22), or when applied to traditional gene-level fine-mapping (Figure 2 and Supplementary Figure 4). We note that a recent study developed a method, GIFT^112^, to jointly associate genes (from a single tissue) with disease while accounting for co-regulation between genes and uncertainty in genetically predicted gene expression; however GIFT only considers genes, not gene-tissue pairs, does not account for the confounding effects of non-mediated genetic variants, and is not computationally scalable to modeling hundreds of gene-tissue pairs in a region, as in our analysis. Another method, CAFEH^19^, was recently developed to identify causal variants underlying GWAS and eQTL in multiple tissues; however, CAFEH does not model co-regulation between gene-tissue pairs and is thus unable to distinguish causal from tagging gene-tissue pairs. Other recent studies have made valuable contributions in nominating causal gene-tissue pairs for disease by linking fine-mapped causal variants to causal genes using tissue-specific SNP-to-gene linking strategies such as Activity-By-Contact or EpiMap enhancer-gene linking^12–14^. However, this approach, unlike TGFM, is based on heuristic prioritization and does not provide direct evidence that the regulatory variant’s effect is mediated by the nominated gene-tissue pair.

We note several limitations of our work. First, TGFM leverages in-sample LD from the GWAS sample^35,113^ (analogous to other fine-mapping methods^36^), but in-sample LD may be unavailable in some applications (e.g. disease consortium meta-analyses). Following ref. ^36^, our recommendation when in-sample LD is unavailable is as follows: if there exists a LD reference panel from a population closely matching the GWAS sample population spanning at least 10% of the GWAS sample size, run the default version of TGFM with the LD reference panel; otherwise, run TGFM assuming a single causal genetic element per locus (in the latter case, no LD reference panel is needed). Second, TGFM may be susceptible to false positives in the case of unassayed causal genetic elements (analogous to other fine-mapping methods^23,30,36,41^), including the specific case of unassayed tissues (analogous to previous studies that have analyzed GTEx data to identify disease-causal tissues based on a genome-wide approach^5,7,8,11^). If the causal gene-tissue pair is not assayed, TGFM may prioritize a correlated assayed gene-tissue pair or a correlated non-mediated genetic variant. In practice, we determined that when the causal tissue is missing, TGFM will tend to prioritize nothing, or prioritize genes from the best proxy tissue, instead of prioritizing genes from an unrelated tissue (Figure 5a). We anticipate that this limitation will be mitigated over time as emerging eQTL data sets increasingly capture diverse tissues, cell types, and cellular contexts^114^. Third, TGFM is only moderately well-powered to detect causal gene-tissue pairs, particularly at lower eQTL sample sizes (Figure 1). In particular, a disease-causal tissue/cell type can have zero or a low number of confidently fine-mapped gene-tissue pairs simply due to limited power of TGFM at low eQTL sample sizes. We anticipate this limitation will be mitigated over time as eQTL data sets increase in size^115^. Fourth, TGFM becomes slightly mis-calibrated at low eQTL sample sizes (e.g. eQTL sample size of 100). In addition, at low eQTL sample sizes, undiscovered causal gene-tissue pairs may be falsely prioritized as non-mediated genetic variants (Supplementary Figure 17). We therefore suggest that some caution is warranted in the interpretation of TGFM PIPs when analyzing eQTL data with sample size of 100 or lower, while also noting that TGFM calibration is far superior to other approaches (Supplementary Figure 13). Again, we anticipate this limitation will be mitigated over time as eQTL data sets increase in size^115^. Fifth, a recent study showed that GWAS and eQTLs studies are well-powered to detect different types of genetic variants, limiting the number of GWAS associations that can be explained by eQTL data at current sample sizes^44^. Nevertheless, TGFM identified causal gene-tissue pairs at hundreds of GWAS loci using current eQTL data, and we anticipate that TGFM will identify a larger number of causal gene-tissue pairs over time as eQTL data sets increase in size^115^ and capture increasingly diverse tissues, cell types, and cellular contexts^114^. Sixth, a related concern is that there is strong evidence that disease-causal genes have lower cis-heritability due to selection^26,28,43^, such that more powerful eQTL discovery in genes with higher cis-heritability may cause TGFM output to be biased towards genes that are not disease-causal, and hence mis-calibrated. Indeed, we observed that TGFM was slightly mis-calibrated in simulations in which causal gene-tissue pairs has lower cis-heritability; however, TGFM’s calibration remained tolerable and notably superior to all other approaches (Supplementary Figure 9). Encouragingly, TGFM was well-calibrated in distinguishing 69 known LDL cholesterol genes from nearby genes (analyzed in Figure 4 of ref. ^27^) (Figure 4d and Supplementary Figures 36, 37), suggesting that our simulation results may overstate this limitation. Seventh, TGFM does not provide unbiased estimates of prior causal probabilities for non-causal tissues and non-mediated variants (Supplementary Figure 25). However, TGFM fine-mapping calibration and power are largely robust to mis-specification of prior causal probabilities (Supplementary Figures 26, 27). Eighth, TGFM (Variant) was slightly mis-calibrated at very high PIP thresholds (PIP=0.99; Supplementary Figure 5f), consistent with increased mis-calibration at high PIP thresholds in variant-level GWAS fine-mapping^41^. Ninth, it is theoretically possible that TGFM could prioritize gene-tissue pairs due to reverse causality, whereby the disease/trait causally impacts the gene-tissue pair.

However, we believe that this is very unlikely in practice, as cis-eQTL variants with a detectable effect on gene expression at current eQTL sample sizes explain a substantial proportion of gene expression variation^25,116,117^ whereas diseases/traits are highly polygenic with each causal variant (outside the HLA locus, which we exclude from our analyses) explaining a small proportion of disease/trait variation^37,118–120^. Tenth, we have focused here on single-ancestry fine-mapping, but an important future direction is to extend this work to multi-ancestry fine-mapping (incorporating multi-ancestry eQTL analysis^121,122^), which is likely to further increase fine-mapping power^123–125^. Eleventh, an important future direction is to extend TGFM to incorporate variant-level functional annotations that are enriched for disease heritability^3,38–40^, which is likely to further increase fine-mapping power^36,126^. Twelfth, another important future direction is to extend TGFM to incorporate gene sets that are enriched for disease heritability explained by cis-predicted expression^26^, which may also further increase fine-mapping power. Finally, we have focused here on cis-genetic components of gene expression, but TGFM could be extended to genetic components of other molecular traits^127–132^.

Despite these limitations, TGFM is a robust and powerful method for fine-mapping causal tissues and genes at disease-associated loci.

## Methods

### TGFM model overview

TGFM estimates the posterior inclusion probability (PIP) for each *genetic element* (gene-tissue pair or genetic variant) to have a non-zero causal effect on disease using a model that includes *mediated* causal effects of each gene-tissue pair (via the cis-genetic component of expression of a given gene in a given tissue) and *non-mediated* causal effects of each genetic variant:

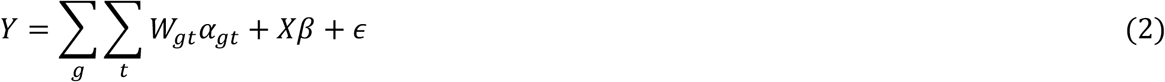

where *Y* denotes the phenotype vector across GWAS individuals, *g* indexes genes, *t* indexes tissues, *X* is the matrix of genotypes, *W*_*gt*_ is the vector of the cis-genetic component of gene expression across GWAS individuals in gene *g* and tissue *t, α*_*gt*_ denotes the (scalar) effect of cis-genetic expression in gene *g* and tissue *t* on the disease or trait, *β* denotes the vector of non-mediated causal effects of each genetic variant on the disease or trait, and *ϵ* denotes environmental noise. We assume that the trait *Y*, the cis-genetic component of gene expression *W*_*gt*_ in each gene *g* and tissue *t*, and the genotype vector of each variant (each column of *X*) are standardized to have mean 0 and variance 1. We model the cis-genetic component of gene expression as a linear combination of variant-level effects:

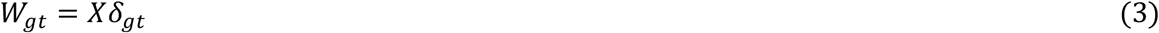

where *δ*_*gt*_ denotes the vector of causal cis-eQTL effect sizes of each variant on gene expression in gene *g* and tissue *t*. We emphasize that we model the phenotype Y as a linear combination of the unobserved true cis-genetic component of gene expression *W*_*gt*_ (a deterministic function of the unobserved true causal eQTL effect sizes *δ*_*gt*_) in each gene and tissue. The *predicted* cis-genetic component of gene expression 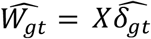 can be estimated (according to predicted causal eQTL effect sizes 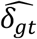, with uncertainty, from finite sample-size eQTL data sets in the specified tissue *t*, and provide noisy estimates of the true unobserved cis-genetic component of gene expression *W*_*gt*_. Later (see below), we explain how TGFM models uncertainty in predicted cis-genetic expression.

TGFM places the Sum of Single Effects (SuSiE)^30,31^ fine-mapping prior distribution on the vector of causal disease (mediated and non-mediated) effect sizes:

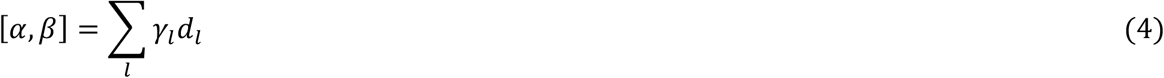

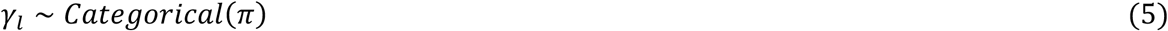

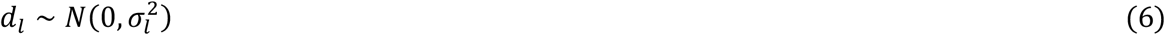

where *α* denotes the vector of expression-mediated causal effects of genetic gene expression of each gene-tissue pair on the disease or trait, *β* denotes the vector of non-mediated causal effects of each genetic variant on the disease or trait, [*α, β*] denotes the concatenated vector of mediated and non-mediated genetic effects, *l* indexes fine-mapping components where a single component represents the disease signal from a single genetic element, *γ*_*l*_ denotes a Categorical random variable indicating which one of the genetic elements disease component *l* comes from, *π* denotes the simplex vector of prior probabilities on each genetic element being causal, *d*_*l*_ denotes a Gaussian random variable specifying the causal effect size of component *l*, and 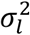 denotes the prior variance on *d*_*l*_. This approach assumes the true causal disease effect sizes originate in a small number (*l*) of genetic elements with non-zero effects.

TGFM will automatically infer posterior distributions on the random variables defining equations 5 and 6 (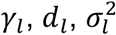 inference details provided below). Posterior inclusion probabilities (PIPs), or the probability that a genetic element has a non-zero effect on disease, can be calculated for each genetic element from these inferred posterior distributions as follows:

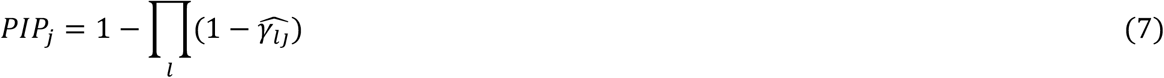

where *j* indexes genetic elements, *l* indexes components, *PIP*_*j*_ denotes the PIP for genetic element *j*, and 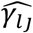 denotes the expected value of the posterior distribution on *γ*_*l*_ for genetic element *j*.

### Sum of Single Effects (SuSiE) fine-mapping prior distribution and inference

The SuSiE prior distribution was developed in refs. ^30,31^ for the purpose of fine-mapping trait-causal variants. We briefly summarize the SuSiE fine-mapping prior distribution here:

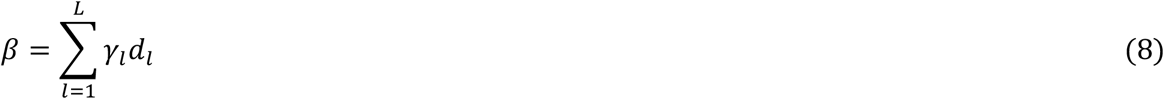

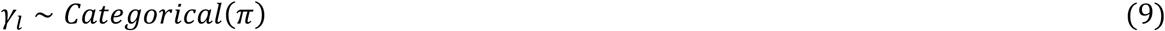

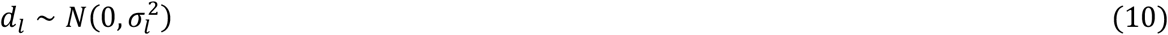

where *β* denotes the vector of causal effects of each genetic variant on the disease or trait, *l* indexes fine-mapping components where a single component represents the disease signal from a single genetic variant, *γ*_*l*_ denotes a Categorical random variable indicating which one of the genetic variants disease component *l* comes from, *π* denotes the simplex vector of prior probabilities on each genetic variant being causal, *d*_*l*_ denotes a Gaussian random variable specifying the causal effect size of component *l*, and 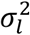 denotes the prior variance on *d*_*l*_.

Briefly, the SuSiE prior distribution assumes that only a subset of variants (*L* total) have non-zero effect (i.e., are causal) on the trait, the effect sizes of each causal variant are independent, and the trait causal effect sizes can be calculated by summing the causal effect size of each of the *L* causal variants. A “single effect” refers to the effect of one of the *L* causal variants; hence why the model is called Sum of Single Effects.

Ref. ^30^ proposed a simple model-fitting approach (to infer posterior distributions on random variables in equations 9 and 10), which the authors referred to as Iterative Bayesian Stepwise Selection (IBSS). Briefly, IBSS iteratively updates each of the *L* single effects while keeping all other single effects fixed. It is computationally simple to update a single effect given fixed values of all other single effects (details of updating a single effect provided in the supplement of Ref. ^30^).

### Overview of TGFM inference

TGFM inference consists of four steps. In step 1, we apply SuSiE to perform eQTL fine-mapping of each gene-tissue pair in the external gene expression data set (estimating a posterior distribution of the causal cis-eQTL effect sizes for each gene-tissue pair). In step 2, we randomly sample 100 cis-predicted expression models for each gene-tissue pair from the posterior distributions of causal cis-eQTL effect sizes estimated in step 1 (Methods). In step 3, we apply SuSiE to perform disease fine-mapping in the target data set (estimating the PIP of each genetic element) 100 times, iterating over the sampled cis-predicted expression models for each gene-tissue pair from step 2. In step 4, we average the results of step 3 across the 100 disease fine-mapping runs. TGFM utilizes a custom implementation of the SuSiE algorithm that provides efficient estimation of PIPs across 100 parallel SuSiE runs that differ only in their cis-predicted expression models.

### TGFM inference Step 1: Estimating causal eQTL effect size distributions from external eQTL data

TGFM inference relies on probability distributions defining the causal eQTL effects for each gene-tissue pair. These causal eQTL effect size distributions are estimated by applying SuSiE^30^ to eQTL data; SuSiE infers the following posterior distribution on the causal eQTL effect sizes for a given gene-tissue pair from eQTL data:

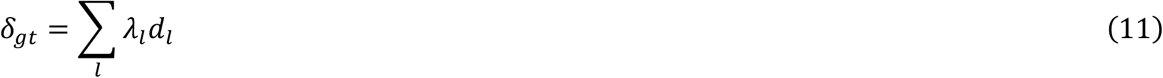

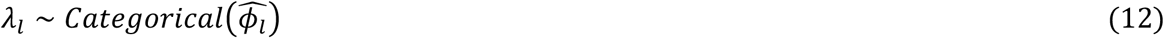

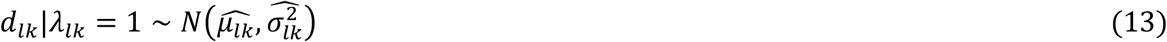

where *δ*_*gt*_ denotes the vector of causal eQTL effect sizes corresponding to the effects of standardized cis-variants on gene *g* in tissue *t, l* indexes fine-mapping components where a single component represents the eQTL signal from a single cis-genetic variant, *λ*_*l*_ denotes a Categorical random variable indicating which one of the genetic variants component *l* comes from, 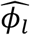 denotes the simplex vector of inferred posterior probabilities on each genetic variant being causal for component *l, d*_*lk*_|*λ*_*lk*_ = 1 denotes a Gaussian random variable specifying the causal effect size of component *l* for variant *k* conditioned on variant *k* being causal for component *l*, and 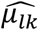 and 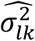 define the inferred posterior mean and variance on *d*_*lk*_|*λ*_*lk*_. The posterior mean causal eQTL effect sizes of this distribution are 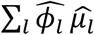.

We restrict TGFM to gene-tissue pairs that are well-predicted by genetic variants, using the SuSiE “purity filter”^30^. Specifically, this filter removes any gene-tissue pair such that all components defining the gene-tissue pair have a minimum absolute correlation between all variants in the component’s 95% credible set less than 0.5. The 95% credible set for a given component is calculated by selecting the minimum set of variants that contain the causal variant with 95% confidence (according to the inferred posterior distribution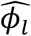).

Although in the present study we utilized SuSiE for generating distributions of causal eQTL effect sizes due to its computational efficiency, the TGFM inference procedure is generalizable to a variety of methods that generate probabilistic cis-predicted expression models potentially including probabilistic/Bayesian multivariable regression methods (for example, BSLMM^133^, SBayesR^134^, or LDpred2^135^) or bootstrapping^136^.

### TGFM inference Step 2: Randomly sample 100 cis-predicted expression models for each gene-tissue pair

Instantiations of the causal eQTL effect sizes (which determine instantiations of cis-predicted expression models) for each gene-tissue pair can be randomly sampled from that gene-tissue pair’s SuSiE-inferred posterior distribution (equations 11-13). To draw a single random sample of the causal eQTL effects for a given gene-tissue pair: (1) randomly sample the causal variant for each of the *l* components from *Categorical* 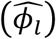 (2) randomly sample the causal eQTL effect size of the sampled causal variant *k* (identified in Step 1) for each of the *l* components from 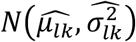 (3) sum the sampled causal eQTL effects across the *l* components.

### TGFM inference step 3: Fine-mapping inference conditioned on a sampled cis-predicted expression models

We describe here tissue-gene fine-mapping inference conditioned on setting the cis genetic component of gene expression for each gene-tissue pair equal to a sampled cis-predicted expression model (see TGFM inference step 2) for that gene-tissue pair. In this setting:

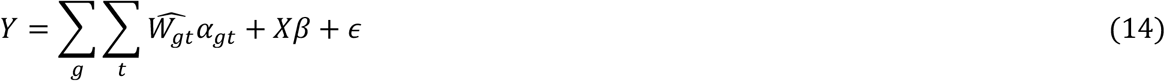

where *g* indexes genes, *t* indexes tissues, 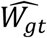 denotes the *predicted* cis-genetic component of gene expression in gene *g* and tissue *t* (as determined by *sampled* cis-predicted expression model’s causal eQTL effect sizes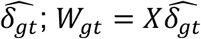, *Y* denotes the phenotype vector across GWAS individuals, *X* is the matrix of genotypes, *α*_*gt*_ denotes the (scalar) effect of cis-genetic expression in gene *g* and tissue *t* on the disease or trait, *β* denotes the vector of non-mediated causal effects of each genetic variant on the disease or trait, and *ϵ* denotes environmental noise. The trait *Y*, the predicted cis-genetic component of gene expression 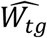 in each gene *g* and tissue *t*, and the genotype vector of each variant (each column of *X*) are standardized to have mean 0 and variance 1. We place SuSiE fine-mapping prior distributions on the disease/trait causal effect sizes (equations 4-6).

In this setting, we use the existing SuSiE software for inference of posterior distributions on the fine-mapping random variables (*γ*_*l*_, *d*_*l*_, and 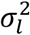 in equations 5-6) and generation of corresponding PIPs for each genetic element (equation 7). We refer to these PIPs as conditional PIPs as they are conditional upon the predicted causal eQTL effect sizes.

SuSiE inference applied to this task requires the following input: GWAS summary statistic z-scores for each non-mediated variant, transcriptome-wide association study (TWAS) summary statistic z-scores for each gene-tissue pair corresponding to the marginal association between predicted genetic gene expression and the trait, in-sample correlations between all pairs of genetic elements (variants and predicted genetic gene-tissue pairs) and specified prior probabilities *π*. We assume the user provides GWAS summary statistic z-scores for each variant, in-sample LD (ie. correlations between all pairs of genetic variants based on the GWAS samples), predicted causal eQTL effect sizes for each gene-tissue pair, and the prior causal probabilities *π* (we discuss below how *π* can be inferred). TWAS summary statistic z-scores and in-sample correlations between all genetic elements can be computed from the user-provided input as follows:

TWAS summary statistic z-scores for a particular gene-tissue pair can be easily calculated from GWAS summary statistic z-scores, in-sample variant LD, and predicted causal cis-eQTL effect sizes defining the genetic component of gene expression for the gene-tissue pair following ref. ^21^:

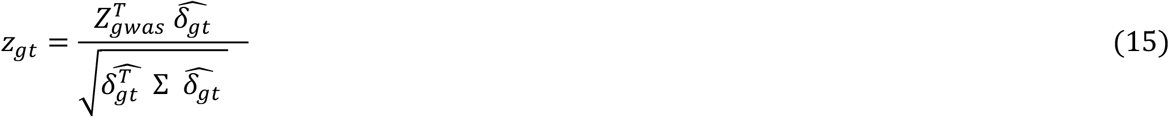

where *z*_*gt*_ denotes the TWAS summary statistic z-score for gene *g* in tissue *t, Z*_*gwas*_ denotes the vector GWAS summary statistic z-scores across variants, 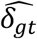 denotes the vector of predicted causal eQTL effect sizes for gene *g* in tissue *t*, and Σ denotes the in-sample variant LD. We note that ref. ^21^ only requires LD from a reference panel (instead of in-sample LD) for TWAS; TGFM instead requires in-sample LD (see Discussion).

The correlation between the predicted genetic gene expression of two genes can be computed from in-sample variant LD and the predicted causal eQTL effect sizes of both genes:

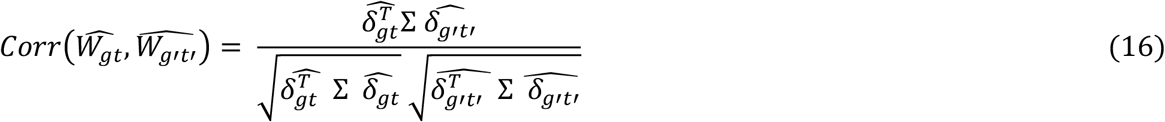

where *gt* indexes one gene-tissue pair and *g*′*t*′ indexes the other gene-tissue pair, 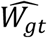 denotes the predicted genetic component of gene expression in gene *g* in tissue *t* across in-sample GWAS individuals, 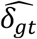 denotes the vector of predicted causal eQTL effect sizes for gene *g* in tissue *t*, and Σ denotes the in-sample variant LD. Similarly, the in-sample correlation between a non-mediated variant and the predicted genetic component of gene-tissue pair is:

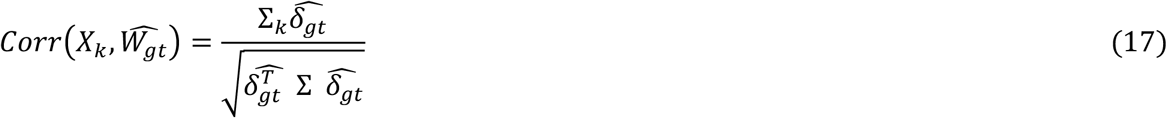

where *k* indexes the non-mediated variant, *gt* indexes the gene tissue pair, *X*_*k*_ is the genotype of variant *k* across in-sample GWAS individuals, 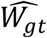 denotes the cis-predicted genetic component of gene expression in gene *g* and tissue *t* across in-sample GWAS individuals, Σ_*k*_ denotes row *k* of the in-sample variant LD matrix, and 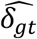 denotes the vector of causal eQTL effect sizes for gene *g* in tissue *t*.

### TGFM inference step 4: Marginalize uncertainty in cis-predicted expression on fine-mapping PIPs by averaging fine-mapping results across 100 runs

TGFM PIPs for a given locus are calculated by marginalizing out the uncertainty in the cis-predicted causal eQTL effect sizes on the conditional PIPs:

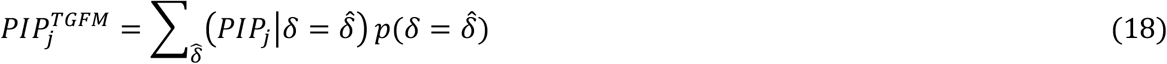

where 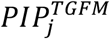 is the TGFM PIP for genetic element *j, δ* is the set of causal eQTL effect sizes for all gene-tissue pairs in the fine-mapping region, 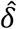 is a specific instantiation of the set of causal eQTL effect sizes for all gene-tissue pairs in the fine-mapping region, 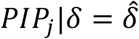 is the conditional PIP of genetic element *j* conditioned on genetic gene expression determined by 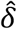, and 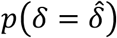 is the probability of estimating causal eQTL effect sizes 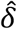 given the eQTL data. The conditional PIPs, 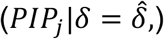, can be inferred using SuSiE fine-mapping of both non-mediated variants and gene-tissue pairs where cis-predicted genetic gene expression is determined by 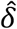 (described in previous Methods subsection, TGFM inference step 3). The causal eQTL effect size distribution, *p*(*δ*), is approximated by the posterior distribution on causal eQTL effect sizes for each gene-tissue pair estimated by applying SuSiE to eQTL data (described in previous Methods subsection, TGFM inference step 1). In practice, we approximate 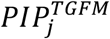 in equation 18 by drawing 100 random samples of causal eQTL effect sizes across gene-tissue pairs from *p*(*δ*) (described in previous Methods subsection, TGFM inference step 2), calculating the conditional PIP 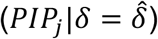 for each of the 100 random samples, and taking the average across 100 conditional PIPs. In the next methods subsection, we discuss how this approach can be extended to integrate the inferred tissue-specific prior.

In practice, we do not run the existing SuSiE package^30,31^ 100 times in series for each fine-mapping region. TGFM utilizes a custom implementation of the SuSiE algorithm that provides efficient estimation of PIPs across 100 parallel runs where each of 100 parallel SuSiE runs differ only in their cis-predicted expression models. The rate-limiting step of each iteration of the SuSiE algorithm involves multiplying the LD matrix with that iteration’s estimate of the trait-causal effect sizes. The custom TGFM implementation exploits the fact that the variant LD matrix (which constitutes the majority of the full correlation matrix of all pairs of genetic elements) is identical across all 100 runs; TGFM uses matrix multiplication to multiply the variant LD matrix with the current causal non-mediated variant effects for each of the 100 runs (corresponding to a single (*K*X*K*)X(*K*X100) matrix multiplication instead of 100 (*K*X*K*)X(*K*X1) multiplications in series where *K* is the number of variants in the region). In addition, the custom TGFM implementation of SuSiE does not compute the ELBO at each iteration for each of the 100 runs; computing the ELBO is computationally intensive and primarily utilized to assess convergence. Instead, we run each of the 100 runs for a pre-specified number of iterations; we use a default of 5 iterations which performed well in simulations (Figure 2). We set the default number of components *l* underlying each of the 100 runs to 10.

The iterative algorithm underlying SuSiE is not guaranteed to reach a global optimum and can get stuck in local optima^30^. We found TGFM was prone to reaching local optima in which a non-causal gene-tissue pair was confidently fine-mapped (i.e., a false positive) when the gene-tissue pair was moderately correlated with multiple independent causal non-mediated variants. This is likely due to the greedy nature of the SuSiE iterative algorithm. We use the following initialization strategy to mitigate convergence on local optima: (1) Run TGFM where the causal effects are initialized to be zero (this is the default initialization used by SuSiE) (2) If the TGFM PIP for any gene-tissue pair in the fine-mapping region is greater than 0.2: (2a) Run SuSiE fine-mapping on only the non-mediated variants (2b) Run TGFM where the non-mediated variant effects are initialized to the converged values from step 2a and the gene-tissue pair effects are initialized to zero (2c) For each of the 100 TGFM runs, select the fitted TGFM model (either (1) or (2b)) with the larger ELBO.

### Inference of tissue-specific TGFM prior causal probabilities

TGFM increases fine-mapping power by specifying tissue-specific prior probabilities for each genetic element in a locus that are informed by genome-wide data, similar to PolyFun^36^. For each trait separately, TGFM assigns one prior causal probability *π*_*t*_ for each gene-tissue pair from tissue *t* and one prior causal probability *π*_*nm*_ for each non-mediated genetic variant where *π*_*t*_ reflects the prior probability that an arbitrary gene from tissue *t* at a disease-associated locus has a non-zero causal effect on the disease/trait and *π*_*nm*_ reflects the prior probability that an arbitrary non-mediated variant at a disease-associated locus has a non-zero causal effect on the disease/trait. We note that *π*_*t*_ is a genome-wide parameter reflecting the probability that an arbitrary gene from tissue *t* has non-zero effect on disease, which is related but distinct from genome-wide expression-mediated disease heritability parameters previously estimated in refs. ^11,26,28^. We infer *π*_*t*_ and *π*_*nm*_ separately for each disease/trait using an iterative algorithm, starting with flat priors, and at each iteration: (1) updating the PIP of each genetic element using a computationally efficient approximation of TGFM (see next paragraph for details) given the current prior causal probabilities, which are normalized to sum to one across genetic elements at each locus, analogous to PolyFun^36^. (2) updating the prior causal probabilities according to:

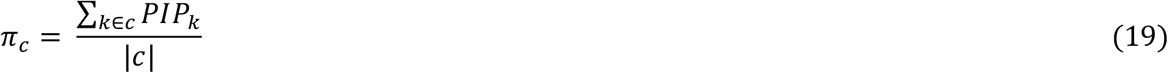

where *c* is the genetic element class (for example a specific tissue or non-mediated variant), *π*_*c*_ is the prior causal probability corresponding to genetic element class *c, k* indexes genetic elements from genetic element class *c, PIP*_*k*_ is the current PIP of genetic element *k*, and |*c*| is the number of genetic elements belonging to genetic element class *c*.

It is computationally prohibitive to run TGFM genome-wide tens to hundreds of times for each trait while updating the prior probabilities at each iteration. We make two approximations to TGFM to allow for efficient computation of genome-wide PIPs at each iteration (and we emphasize that these approximations are only used for inference of the prior). First, we run TGFM with a single cis-predicted expression model for each gene-tissue pair (based on SuSiE posterior mean causal cis-eQTL effect sizes) instead of averaging results across 100 sampled cis-predicted expression models. We refer to this approach as TGFM (no sampling). Thus, PIPs at each iteration can be inferred by applying SuSiE to fine-map both non-mediated variants and gene-tissue pairs where genetic gene expression of each gene-tissue pair is determined by the posterior mean causal eQTL effect sizes. While we show TGFM (no sampling) generates poorly calibrated PIPs (Supplementary Figure 18), using TGFM (no sampling) to infer causal tissues results in well-calibrated p-values in simulations (Supplementary Figure 28a), perhaps because the causal prior probabilities integrate information across the genome (equation 19) and fine-mapping errors resulting from uncertainty in the genetic component of gene expression will be averaged out across all genes in the genome.

Second, we only run TGFM (no sampling) inference once, during the first iteration. After running TGFM (no sampling) in the first iteration, we save the resulting Bayes Factors^30^ for each component-genetic element pair; a Bayes Factor reflects the relative support for including that genetic element in that component of the model, irrespective of the prior probabilities. PIPs can be easily calculated based on the Bayes Factors and the current prior:

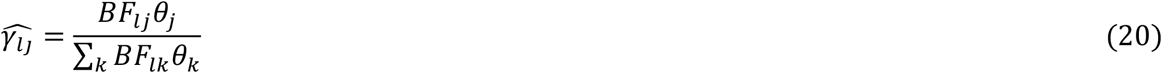

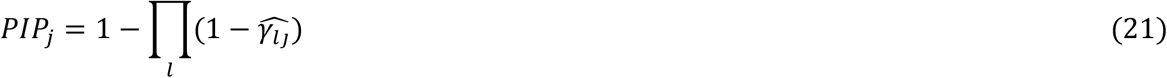

where *l* indexes fine-mapping components, *j* and *k* index genetic elements in the fine-mapping region, 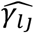 denotes the expected value of the posterior distribution on *γ*_*l*_ for genetic element *j, BF*_*lj*_ denotes the Bayes Factor for genetic element *j* on component *l, θ*_*j*_ denotes the normalized prior causal probability of genetic element *j*, and *PIP*_*j*_ is the TGFM (no sampling) PIP for genetic element *j*. We do not recalculate the Bayes Factors after the first iteration, and simply used the saved Bayes Factors from the first iteration in all subsequent iterations. This approximation is reasonable as while we expect PIPs to change with the evolving prior, we do not expect posterior mean causal effect sizes of each fine-mapping component to drastically change with the evolving prior, ultimately leaving the Bayes Factors stable.

During inference of the causal prior probabilities, TGFM (no sampling) is run genome-wide on overlapping 3Mb windows^36^. The prior probability updates at each iteration (equation 16) are calculated from PIPs corresponding to genetic elements located in the middle Mb of these 3 Mb windows. To limit to windows with at least one disease causal signal, we remove 3Mb windows from the prior probability inference procedure that do not pass the SuSiE “purity filter” (see above) after running TGFM (no sampling) with a uniform prior.

In addition, TGFM inference can also rely on probability distributions defining the uncertainty in our estimated prior causal probabilities (see below). Empirical distributions, as well as significance testing, on the causal prior probabilities can be calculated using 100 iterations of bootstrapping^136^ across the genome (we refer to this as “genomic bootstrapping”). Specifically, for each of the 100 bootstraps, we randomly sample *T* fine-mapping 3Mb windows with replacement (assuming *T* total 3Mb fine-mapping windows for the disease/trait) and run the iterative algorithm on the bootstrapped regions. This procedure results in 100 empirical samples of the causal prior probabilities which reflect their estimation uncertainty. These 100 empirical samples are input directly into the sampling procedure underlying TGFM inference (see below). Significance testing of whether the prior causal probability is greater than zero for a particular genetic element class can be generated using a z-score computed from the mean and standard error of the bootstrapped distribution. Any bootstrapped prior causal probability < 1x10^−15^ was set to 0 when calculating z-scores. If all bootstrapped prior causal probabilities corresponding to a given trait-tissue pair were equal to 0, the z-score was undefined but subsequently set to 0. For a single analyzed trait, we assess significance using Benjamini-Hochberg^137^ FDR correction across all tissues and/or cell types included in the analysis.

Inference of TGFM (no sampling) is performed using the function ‘susie_rss’ from the SuSiE package^30,31^ with default parameters. We used the same initialization strategy that was used by TGFM (described above). The iterative algorithm for inference of prior causal probabilities was run for 400 iterations.

### TGFM inference including uncertainty in tissue-specific prior causal probabilities

The previous subsection described inference of probability distributions defining the tissue-specific prior causal probabilities. Here we described an extension of TGFM inference step 4 that integrates out uncertainty in *both* cis-predicted causal eQL effect sizes and the prior causal probabilities on the conditional PIPs:

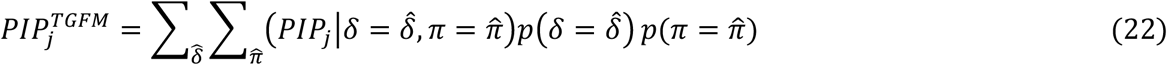

where 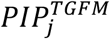 is the TGFM PIP for genetic element *j, δ* is the set of causal eQTL effect sizes for all gene-tissue pairs in the fine-mapping region, 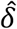 is a specific instantiation of the set of causal eQTL effect sizes for all gene-tissue pairs in the fine-mapping region, *π* are the causal prior probabilities, 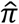 are a specific instantiation of the causal prior probabilities, 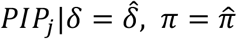 is the conditional PIP of genetic element *j* conditioned on genetic gene expression determined by 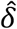 and causal prior probabilities equal to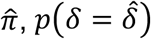,is the probability of estimating causal eQTL effect sizes 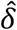 given the eQTL data, and 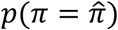 is the probability of estimating prior causal probabilities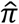. In practice, we approximate 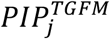 by drawing 100 random samples of causal eQTL effect sizes across gene-tissue pairs from *p*(*δ*) and prior causal probabilities from *p*(*π*), calculating the conditional PIP 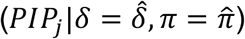 for each of the 100 random samples, and taking the average across 100 conditional PIPs.

### Calculating gene-level PIPs with TGFM

We define a gene as causal for a disease/trait if there exists at least one tissue where the gene-tissue pair is causal for the trait. Gene-level PIPs can be computed by aggregating gene-tissue pair fine-mapping results across all gene-tissue pairs corresponding to the gene of interest:

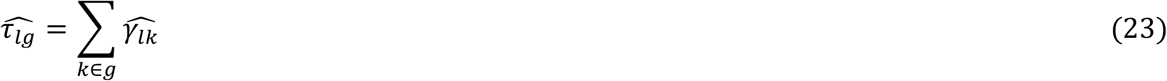

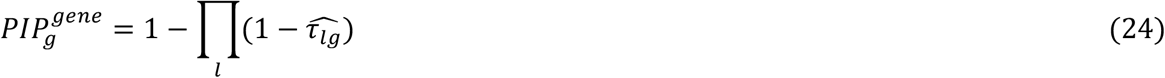

where *l* indexes fine-mapping components, *g* indexes genes, *k* ∈ *g* indexes all gene-tissue pairs corresponding to gene *g*, 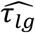 denotes the expected value of the posterior distribution on *γ*_*l*_ for gene *g*, 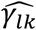 denotes the expected value of the posterior distribution on *γ*_*l*_ for gene-tissue pair *k*, and 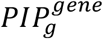 is the gene-level PIP corresponding to gene *g*. Similar to TGFM PIPs for variants and gene-tissue pairs, we are describing here the calculation of conditional gene PIPs (conditional upon a given instantiation of predicted causal cis-eQTL effect sizes and prior causal probabilities). These conditional gene PIPs will be averaged across 100 samples of cis-predicted genetic gene expression and predicted causal prior probabilities (equation 22).

This approach can also be used to calculate PIPs for causal genes in a specified subset of tissues, or gene-tissue subset PIPs. For example, identifying gene-tissue subset pairs for the subset of adipose tissues (defined as adipose subcutaneous U adipose visceral). Gene-tissue subset PIPs can be computed by aggregating (as done in equations 23-24) gene-tissue pair fine-mapping results across all gene-tissue pairs corresponding to the gene of interest from tissues in the tissue subset of interest.

### Simulation framework

We used real genotypes from unrelated UK Biobank British (UKBB) samples^32^ to simulate both gene expression phenotypes (for each gene-tissue pair) and quantitative trait phenotypes. Default simulation parameters were specified as follows: the gene expression sample size ranged from 300 to 1000, plus a simulation including tissues with unequal sample sizes (denoted as 100-300); the tissue sample sizes of the ten simulated tissues in the 100-300 simulation were randomly set (without replacement) to 320, 320, 320, 320, 320, 122, 116, 115, 108, and 101 in order to approximately match the upper and lower range of sample sizes in our analyses of GTEx tissues (Supplementary Table 6); the quantitative trait sample size was set to 100,000 (disjoint from gene expression samples); we analyzed 426,593 SNPs and 1,976 genes on chromosome 1; the number of tissues was set to 10, of which 2 were causal for the quantitative trait; the quantitative trait architecture was simulated to have average polygenicity^37^, consisting of 2,700 causal non-mediated variants and 300 causal gene-tissue pairs (150 for each causal tissue allowing for the option of two causal gene-tissue pairs from the same gene) with the expected heritability per causal genetic element (non-mediated variant or gene-tissue pair) set to 0.0001 (expected quantitative trait heritability of 0.3, 10% of which was mediated through gene expression, consistent with genome-wide estimates from MESC^28^); causal non-mediated variants were randomly selected with probability proportional to their expected per-variant heritability based on baseline-LD model annotations^3,38,39^ (estimated using S-LDSC^3^ applied to the UKBB trait White blood cell count) in order to make the simulations as realistic as possible. We simulated the genetic architecture of gene expression across related tissues using an approach similar to the approach implemented in ref. ^11^: we simulated all 1,976 protein-coding genes on chromosome 1 to be expressed in all tissues, with 50% of these gene-tissue pairs being cis-genetically heritable (only cis-genetically heritable gene-tissue pairs could have a simulated causal effect on the trait, though we considered all expressed gene-tissue pairs for fine-mapping, not just those that were heritable); each heritable gene-tissue pair was randomly assigned 5 causal *cis*-eQTLs within 100Kb of the gene’s TSS, 3 of the 5 causal *cis*-eQTLs were shared across tissues and 2 of the 5 causal *cis*-eQTLs were specific to each tissue; each causal *cis*-eQTL explains 1.5% of the variance of each gene-tissue pair (resulting in an average gene heritability of .075); effect sizes of shared causal cis-eQTLs covaried across tissues as follows (based on ref. ^11^): the tissues were split into 3 categories to mimic biological tissue modules in GTEx^25^ (tissues 1-3, tissues 4-6, and tissues 7-10) and the correlation of shared cis-eQTL effect sizes across tissues was set to 0.8 and 0.74 for tissues in the same tissue category and across tissues in different tissue categories, respectively.

Our simulation indicates that PIPs reported by TGFM are slightly anti-conservative with respect to (1 – average PIP) (Supplementary Figures 1, 5), consistent with previous simulations of variant-level fine-mapping methods using polygenic trait architectures^36,41^. Following ref. ^36^, we circumvent this problem by using an alternative FDR estimator given by (1 – PIP threshold); setting all PIPs greater than the specified PIP threshold equal to the PIP threshold. For example, at a PIP threshold of 0.9, we treat all genetic elements with PIP ≥ 0.9 as if they had PIP = 0.9.

We compared TGFM to several published methods: coloc, FOCUS, cTWAS, JLIM, and SMR. coloc was run, using default settings, independently for each gene-tissue pair; the summary statistics used in the coloc analysis were restricted to variants within 100kb of the gene’s TSS. FOCUS was run using default settings; the genotype data used by FOCUS corresponded to the in-sample genotype of the simulated GWAS individuals, and the gene models used by FOCUS were SuSiE fine-mapping^30^ posterior mean causal eQTL effect sizes restricted to variants within 100Kb of the gene’s TSS, in order to make FOCUS maximally comparable to TGFM. cTWAS was run using default settings; variant LD used by cTWAS was calculated from the in-sample genotype of the simulated GWAS individuals while utilizing European ancestry genomic regions (build b37) built into the cTWAS package, and the gene models used by cTWAS were lasso gene models, restricted to variants within 100Kb of the gene’s TSS, produced by FUSION^21^, as the cTWAS software recommends using a sparse gene model. JLIM was run using default settings, independently for each gene-tissue pair; the summary statistics used in the JLIM analysis were restricted to variants within 100kb of the gene’s TSS, and the genotype data used by JLIM was the Non-Finish European, build 37, reference genotype available for download on the JLIM GitHub page (https://github.com/cotsapaslab/jlim), in concordance with JLIM recommendations. SMR was run, using default settings, independently for each gene-tissue pair; the summary statistics used in the SMR analysis were restricted to variants within 2Mb of the gene’s TSS, in concordance with SMR recommendations, though we exclude trans summary statistics from SMR analysis (despite their recommended inclusion) as our simulation was limited to only chromosome 1; the genotype data used by SMR corresponded to the in-sample genotype of the simulated GWAS individuals.

### UK Biobank GWAS summary statistics and in-sample LD

We applied to TGFM to 45 of the 49 UK Biobank traits analyzed via functionally informed fine-mapping in ref. ^36^ (average *N*=316K; Supplementary Table 5); the four excluded traits were Dermatology, Diabetes (any), Endocrine Diabetes, and Childless; these four diseases and traits were excluded due to redundancy and low heritability. We considered the set of 10,545,304 UK Biobank imputed variants with MAF ≥ 0.5% and INFO score ≥ 0.6, similar to previous work^36,138^. We used GWAS summary statistics that were generated and described in ref. ^36^. Briefly, the summary statistics were computed in ref. ^36^ from n=337,426 unrelated British-ancestry individuals in UK Biobank using BOLT-LMM^139^ adjusting for sex, age and age squared, assessment center, genotyping platform, the top 20 genotyping principal components, and dilution factor for biochemical traits (see ref. ^36^ for complete details). We used Liftover^140^ to convert variant positions from hg19 to hg38. Z-scores used by TGFM were computed from the Bolt-LMM output as follows: as the noninfinitesimal version of BOLT-LMM does not calculate effect sizes, we calculated z-scores by taking the square root of the BOLT-LMM chi-squared statistics and multiplying them by the sign of the effect size estimate from the infinitesimal version of BOLT-LMM.

We computed in-sample variant LD matrices using 337,426 unrelated British-ancestry individuals in UK Biobank (same individuals as summary statistics); missing values were imputed by the mean of a variant across individuals.

### Overlapping 3Mb loci used for fine-mapping

We applied TGFM to fine-map each of the 2,682 overlapping 3Mb loci spanning the entire genome. Analogous to ref. ^36^, the overlapping 3Mb loci had 1 Mb spacing between the start points of consecutive loci, were limited to autosomal chromosomes, and did not include 3 long-range LD regions including the MHC region (chromosome 6 positions 25499772-33532223, chromosome 8 positions 8142478-12142491, and chromosome 11 positions 45978449-57232526 in hg38; lifted over from long-range LD regions ignored in ref. ^36^). Distinct from the windows generated in ref. ^36^, for each disease/trait, we limited TGFM fine-mapping to loci with at least 50 genetic variants and at least one genetic variant with marginal GWAS p-value less than 1e-5.

### GTEx cis-predicted expression models

We analyzed GTEx^25^ data from 47 GTEx tissues, which were aggregated into 38 meta tissues of similar sample size consisting of European-ancestry individuals (average *N*=259, range: *N*=101-320 individuals, 23 meta-tissues with *N*=320; Supplementary Table 6) to reduce heterogeneity in eQTL sample sizes across tissues. Testis tissue was removed from analysis as it has outlier (cis- and trans-) eQTL discovery power after controlling for sample size (see ref. ^25^ Figure 2C). Meta-tissues were constructed using the same individuals and tissue aggregation strategy as described in ref. ^11^. Normalized expression matrices and covariates used in GTEx consortium’s single-tissue cis-eQTL analysis^25^ were downloaded from the GTEx portal (https://gtexportal.org/home/datasets) for each of the 47 analyzed tissues. In each tissue, we subset individuals to those composing the corresponding meta-tissue, and then re-standardize gene expression of each gene to have mean zero and variance one in each subsetted tissue. Cis-eQTLs were called in each tissue independently while controlling for covariates and limiting to variant-gene pairs such that the variant is within 500Kb of the gene’s TSS. For each meta-tissue, we removed variants with MAF < .05 across samples in the meta-tissue, were strand-ambiguous, or did not overlap the 10,545,304 analyzed UK Biobank variants. Cis-predicted expression models were generated using SuSiE^30,31^ applied to eQTL summary statistics and eQTL in-sample LD matrices (using the function ‘susie_rss’ from the SuSiE package^30,31^ with default parameters) for each (gene, meta-tissue) pair, independently. If a meta-tissue is composed of more than 1 constituent tissues, meta-analyzed eQTL summary statistics were generated using a fixed-effect meta-analysis across constituent tissues and meta-analyzed in-sample LD was generated by computing variant-variant correlations across all samples composing the meta-analyzed tissue. After removing gene-tissue pairs that did not pass the SuSiE “purity filter” (see above), we identified 119,270 gene-tissue pairs with a cis-predicted expression model; all 119,270 cis-predicted expression models are publicly available (see Data availability).

### Pseudobulk PBMC cis-predicted expression models

Pseudobulk PBMC cis-predicted expression models were used as input to TGFM in the analysis where GTEx whole blood (*N*=320) was replaced with pseudobulk PBMC (Figure 5b).

PBMC single-cell eQTL (described and generated in ref. ^33^) expression data was downloaded from the Human Cell Atlas Data Coordination Platform and genotype data downloaded from dbGaP (accession number phs002812.v1.p1). We removed individuals that were not European ancestry or had fewer than 2,500 detected cells. We generated pseudobulk expression across all cells (regardless of cell-type assignment) for all individuals with greater than 5 cells. We removed genes that were expressed in less than 80% of the pseudobulk samples. Pseudobulk expression was transformed using EdgeR’s logCPM function^141^ followed by normalizing each gene to have mean 0 and variance 1. The top 10 expression PCs were calculated based on the normalized expression matrix.

Cis-eQTLs were called while controlling for covariates (the top 10 expression PCs) and limiting to variant-gene pairs such that the variant is within 500Kb of the gene’s TSS. We removed variants with MAF < 0.05 across samples, that were strand-ambiguous, or did not overlap the 10,545,304 analyzed UK Biobank variants. Cis-predicted expression models were generated using SuSiE^30,31^ applied to eQTL summary statistics and eQTL in-sample LD matrices (using the function ‘susie_rss’ from the SuSiE package^30,31^ with default parameters) for each gene, independently. After removing genes that did not pass the SuSiE “purity filter” (see above), we identified 923 genes with a cis-predicted expression model; all 923 cis-predicted expression models are publicly available (see Data availability).

### PBMC cis-predicted expression models in 9 fine-grained cell types

PBMC single-cell eQTL (described and generated in ref. ^33^) expression data was downloaded from the Human Cell Atlas Data Coordination Platform and genotype data downloaded from dbGaP (accession number phs002812.v1.p1). We removed individuals that were not European ancestry or had fewer than 2,500 detected cells. We generated pseudobulk expression in the 9 most abundant cell types (cell type assignment determined by ref. ^33^) for all individuals in each cell type with greater than 5 cells. In each of the 9 cell types separately, we removed genes that were expressed in less than 80% of that cell type’s pseudobulk samples. Pseudobulk expression in each cell type was transformed using EdgeR’s logCPM function^141^ followed by normalizing each gene to have mean 0 and variance 1. The top 10 expression PCs for each cell type were calculated based on the normalized expression matrix. The number of pseudobulk samples per cell type is described in Supplementary Table 19 (average *N*=112).

Cis-eQTLs were called in each cell type independently while controlling for covariates (the top 10 expression PCs) and limiting to variant-gene pairs such that the variant is within 500Kb of the gene’s TSS. For each cell type, we removed variants with MAF < 0.05 across samples in the cell type, that were strand-ambiguous, or did not overlap the 10,545,304 analyzed UK Biobank variants. Cis-predicted expression models were generated using SuSiE^30,31^ applied to eQTL summary statistics and eQTL in-sample LD matrices (using the function ‘susie_rss’ from the SuSiE package^30,31^ with default parameters) for each (gene, cell type) pair, independently. After removing gene-PBMC cell type pairs that did not pass the SuSiE “purity filter” (see above), we identified 1,851 gene-PBMC cell type pairs with a cis-predicted expression model; all 1,851 cis-predicted expression models are publicly available (see Data availability).

### Tissue-trait significance using S-LDSC with chromatin data

We sought to evaluate whether the tissue-trait pairs identified as statistically significant via the TGFM tissue-specific prior were also identified as statistically significant in S-LDSC analyses using chromatin data in ref. ^3^. This analysis is summarized in Figure 4b. The chromatin data in ref. ^3^ consisted of data from 10 cell-type groups: adrenal & pancreas, CNS, cardiovascular, connective & bone, gastrointestinal, immune & hematopoietic, kidney, liver, skeletal muscle, and other. Following ref. ^3^, we ran S-LDSC on each of the 45 analyzed traits and 10 cell-type groups using the baselineLD annotations and one of the cell-type group chromatin annotations at a time (i.e., one run of S-LDSC for each trait-cell-type group pair). Significance of a trait-cell-type group pair was assessed using the S-LDSC p-value of the S-LDSC coefficient corresponding the cell-type group annotation. Following the approach taken in ref. ^7^, we assigned one of the 10 cell-type groups to each of the 38 analyzed GTEx tissues (Supplementary Table 10), and used the S-LDSC p-value from the matched cell-type group for each GTEx tissue. Any tissue assigned to the cell-type group “other” was excluded from analysis because the cell-type group “other” represents a heterogenous mix of tissues and cell-types.

### Proxy tissues in tissue ablation analysis

We assigned a tissue a “proxy tissue” if there existed a single tissue that had highly correlated cis-eQTL effect sizes in ref. ^25^ (see Figure 6a of ref. ^25^). We selected 6 pairs of proxy tissues: artery aorta and artery tibial, skin (sun exposed) and skin (not sun exposed), heart left ventricle and heart atrial appendage, adipose subcutaneous and adipose visceral, liver and pancreas, and whole blood and spleen. Many tissues will therefore have no assigned proxy tissue; this was a conservative decision to ensure proxy tissues exclusively captured highly correlated tissues. Proxy tissues were used an analyzing results from tissue ablation analysis (Figure 5a).

## Data availability

We have made TGFM PIPs for gene-tissue pairs, gene-PBMC cell type pairs, genes, and non-mediated variants across 45 diseases/traits (for both analyses of 38 GTEx tissues + analyses of 38 GTEx tissues and 9 PBMC cell types) publicly available at https://doi.org/10.7910/DVN/S26PFI, GTEx cis-predicted expression models for all gene-tissue pairs publicly available at https://doi.org/10.7910/DVN/8IPOPK, pseudobulk PBMC cis-predicted expression for all gene-PBMC pairs publicly available at https://doi.org/10.7910/DVN/8UL8XB, PBMC cis-predicted expression models for all gene-PBMC cell type pairs publicly available at https://doi.org/10.7910/DVN/A6K9QW, GWAS summary statistics for all 45 diseases/traits publicly available at https://doi.org/10.7910/DVN/GTEGPE. To limit the use of computational resources, we refer the reader to UK Biobank in-sample LD (337K unrelated British-ancestry samples) from ref. ^36^, which is publicly available at https://registry.opendata.aws/ukbb-ld/. The UK Biobank resource is publicly available via application (http://www.ukbiobank.ac.uk/).

## Code availability

Software implementing TGFM and code generating all results of the paper are available at https://github.com/BennyStrobes/TGFM_workflow.

## Acknowledgements

We are grateful to Arun Durvasula and Xilin Jiang for helpful discussions. This research was conducted using the UK Biobank resource under application no. 16549 and funded by National Institutes of Health (NIH) grants R01 MH101244, R37 MH107649, R01 HG006399, R01 MH115676, U01 HG012009, R56 HG013083 and F32 HG012889. The funders had no role in study design, data collection and analysis, decision to publish or preparation of the manuscript.

## Notes

### Competing Interest Statement

The authors have declared no competing interest.

### Author Declarations

The study used only openly available human data that were originally located at http://www.ukbiobank.ac.uk/ and https://gtexportal.org/home/datasets.

### Summary of Updates

We have made many revisions to the paper including a comparison to cTWAS, an empirical assessment of the calibration of the method applied to real traits, and a validation of the trait-tissue pairs prioritized by TGFM.

## References

1. Hekselman, I. & Yeger-Lotem, E. Mechanisms of tissue and cell-type specificity in heritable traits and diseases. Nat. Rev. Genet. 21, 137–150 (2020).

2. Trynka, G. et al. Chromatin marks identify critical cell types for fine mapping complex trait variants. Nat. Genet. 45, 124–130 (2013).

3. Finucane, H. K. et al. Partitioning heritability by functional annotation using genome-wide association summary statistics. Nat. Genet. 47, 1228–1235 (2015).

4. Kundaje, A. et al. Integrative analysis of 111 reference human epigenomes. Nature 518, 317–330 (2015).

5. Ongen, H. et al. Estimating the causal tissues for complex traits and diseases. Nat. Genet. 49, 1676–1683 (2017).

6. Calderon, D. et al. Inferring relevant cell types for complex traits by using single-cell gene expression. Am. J. Hum. Genet. 101, 686–699 (2017).

7. Finucane, H. K. et al. Heritability enrichment of specifically expressed genes identifies disease-relevant tissues and cell types. Nat. Genet. 50, 621–629 (2018).

8. Shang, L., Smith, J. A. & Zhou, X. Leveraging gene co-expression patterns to infer trait-relevant tissues in genome-wide association studies. PLoS Genet. 16, e1008734 (2020).

9. Zhang, M. J. et al. Polygenic enrichment distinguishes disease associations of individual cells in single-cell RNA-seq data. Nat. Genet. 54, 1572–1580 (2022).

10. Jagadeesh, K. A. et al. Identifying disease-critical cell types and cellular processes by integrating single-cell RNA-sequencing and human genetics. Nat. Genet. 54, 1479–1492 (2022).

11. Amariuta, T., Siewert-Rocks, K. & Price, A. L. Modeling tissue co-regulation estimates tissue-specific contributions to disease. Nat. Genet. 55, 1503–1511 (2023).

12. Fulco, C. P. et al. Activity-by-contact model of enhancer–promoter regulation from thousands of CRISPR perturbations. Nat. Genet. 51, 1664–1669 (2019).

13. Nasser, J. et al. Genome-wide enhancer maps link risk variants to disease genes. Nature 593, 238–243 (2021).

14. Boix, C. A., James, B. T., Park, Y. P., Meuleman, W. & Kellis, M. Regulatory genomic circuitry of human disease loci by integrative epigenomics. Nature 590, 300–307 (2021).

15. Downes, D. J. et al. Identification of LZTFL1 as a candidate effector gene at a COVID-19 risk locus. Nat. Genet. 53, 1606–1615 (2021).

16. Kosoy, R. et al. Genetics of the human microglia regulome refines Alzheimer’s disease risk loci. Nat. Genet. 54, 1145–1154 (2022).

17. Giambartolomei, C. et al. Bayesian test for colocalisation between pairs of genetic association studies using summary statistics. PLoS Genet. 10, e1004383 (2014).

18. Hormozdiari, F. et al. Colocalization of GWAS and eQTL signals detects target genes. Am. J. Hum. Genet. 99, 1245–1260 (2016).

19. Arvanitis, M., Tayeb, K., Strober, B. J. & Battle, A. Redefining tissue specificity of genetic regulation of gene expression in the presence of allelic heterogeneity. Am. J. Hum. Genet. 109, 223–239 (2022).

20. Gamazon, E. R. et al. A gene-based association method for mapping traits using reference transcriptome data. Nat. Genet. 47, 1091–1098 (2015).

21. Gusev, A. et al. Integrative approaches for large-scale transcriptome-wide association studies. Nat. Genet. 48, 245–252 (2016).

22. Wainberg, M. et al. Opportunities and challenges for transcriptome-wide association studies. Nat. Genet. 51, 592–599 (2019).

23. Schaid, D. J., Chen, W. & Larson, N. B. From genome-wide associations to candidate causal variants by statistical fine-mapping. Nat. Rev. Genet. 19, 491–504 (2018).

24. Mancuso, N. et al. Probabilistic fine-mapping of transcriptome-wide association studies. Nat. Genet. 51, 675–682 (2019).

25. The GTEx Consortium et al. The GTEx Consortium atlas of genetic regulatory effects across human tissues. Science 369, 1318–1330 (2020).

26. Siewert-Rocks, K. M., Kim, S. S., Yao, D. W., Shi, H. & Price, A. L. Leveraging gene co-regulation to identify gene sets enriched for disease heritability. Am. J. Hum. Genet. 109, 393–404 (2022).

27. Zhao, S. et al. Adjusting for genetic confounders in transcriptome-wide association studies improves discovery of risk genes of complex traits. Nat. Genet. 56, 336–347 (2024).

28. Yao, D. W., O’Connor, L. J., Price, A. L. & Gusev, A. Quantifying genetic effects on disease mediated by assayed gene expression levels. Nat. Genet. 52, 626–633 (2020).

29. Benner, C. et al. FINEMAP: efficient variable selection using summary data from genome-wide association studies. Bioinformatics 32, 1493–1501 (2016).

30. Wang, G., Sarkar, A., Carbonetto, P. & Stephens, M. A simple new approach to variable selection in regression, with application to genetic fine mapping. J. R. Stat. Soc. Series B Stat. Methodol. 82, 1273–1300 (2020).

31. Zou, Y., Carbonetto, P., Wang, G. & Stephens, M. Fine-mapping from summary data with the “Sum of Single Effects” model. PLoS Genet. 18, e1010299 (2022).

32. Bycroft, C. et al. The UK Biobank resource with deep phenotyping and genomic data. Nature 562, 203–209 (2018).

33. Perez, R. K. et al. Single-cell RNA-seq reveals cell type-specific molecular and genetic associations to lupus. Science 376, eabf1970 (2022).

34. Abdellaoui, A., Yengo, L., Verweij, K. J. H. & Visscher, P. M. 15 years of GWAS discovery: Realizing the promise. Am. J. Hum. Genet. 110, 179–194 (2023).

35. Pasaniuc, B. & Price, A. L. Dissecting the genetics of complex traits using summary association statistics. Nat. Rev. Genet. 18, 117–127 (2017).

36. Weissbrod, O. et al. Functionally informed fine-mapping and polygenic localization of complex trait heritability. Nat. Genet. 52, 1355–1363 (2020).

37. O’Connor, L. J. et al. Extreme polygenicity of complex traits is explained by negative selection. Am. J. Hum. Genet. 105, 456–476 (2019).

38. Gazal, S. et al. Linkage disequilibrium–dependent architecture of human complex traits shows action of negative selection. Nat. Genet. 49, 1421–1427 (2017).

39. Gazal, S. et al. Functional architecture of low-frequency variants highlights strength of negative selection across coding and non-coding annotations. Nat. Genet. 50, 1600–1607 (2018).

40. Gazal, S., Marquez-Luna, C., Finucane, H. K. & Price, A. L. Reconciling S-LDSC and LDAK functional enrichment estimates. Nat. Genet. 51, 1202–1204 (2019).

41. Cui, R. et al. Improving fine-mapping by modeling infinitesimal effects. Nat. Genet. 56, 162–169 (2024).

42. GTEx Consortium. Genetic effects on gene expression across human tissues. Nature 550, 204–213 (2017).

43. Wang, X. & Goldstein, D. B. Enhancer domains predict gene pathogenicity and inform gene discovery in complex disease. Am. J. Hum. Genet. 106, 215–233 (2020).

44. Mostafavi, H., Spence, J. P., Naqvi, S. & Pritchard, J. K. Systematic differences in discovery of genetic effects on gene expression and complex traits. Nat. Genet. (2023) doi:10.1038/s41588-023-01529-1.

45. Chun, S. et al. Limited statistical evidence for shared genetic effects of eQTLs and autoimmune-disease-associated loci in three major immune-cell types. Nat. Genet. 49, 600–605 (2017).

46. Zhu, Z. et al. Integration of summary data from GWAS and eQTL studies predicts complex trait gene targets. Nat. Genet. 48, 481–487 (2016).

47. Mancuso, N. et al. Large-scale transcriptome-wide association study identifies new prostate cancer risk regions. Nat. Commun. 9, 1–11 (2018).

48. Nguyen, P. et al. Liver lipid metabolism. J. Anim. Physiol. Anim. Nutr. (Berl.) 92, 272–283 (2008).

49. Homan, T. D., Bordes, S. J. & Cichowski, E. Physiology, Pulse Pressure. (StatPearls Publishing, 2023).

50. Di Sabatino, A. et al. Depletion of immunoglobulin M memory B cells is associated with splenic hypofunction in inflammatory bowel disease. Am. J. Gastroenterol. 100, 1788–1795 (2005).

51. Giuffrida, P. et al. Defective spleen function in autoimmune gastrointestinal disorders. Intern. Emerg. Med. 15, 225–229 (2020).

52. Khan, U. & Ghazanfar, H. T Lymphocytes and Autoimmunity. in International Review of Cell and Molecular Biology vol. 341 125–168 (Elsevier, 2018).

53. Sun, L., Su, Y., Jiao, A., Wang, X. & Zhang, B. T cells in health and disease. Signal Transduct. Target. Ther. 8, 1–50 (2023).

54. Biedermann, T., Skabytska, Y., Kaesler, S. & Volz, T. Regulation of T cell immunity in atopic dermatitis by microbes: The yin and yang of cutaneous inflammation. Front. Immunol. 6, (2015).

55. Roediger, B. & Schlapbach, C. T cells in the skin: Lymphoma and inflammatory skin disease. J. Allergy Clin. Immunol. 149, 1172–1184 (2022).

56. Ho, A. W. & Kupper, T. S. T cells and the skin: from protective immunity to inflammatory skin disorders. Nat. Rev. Immunol. 19, 490–502 (2019).

57. Emdin, C. A. et al. Genetic association of waist-to-hip ratio with cardiometabolic traits, type 2 diabetes, and coronary heart disease. JAMA 317, 626 (2017).

58. Weeks, E. M. et al. Leveraging polygenic enrichments of gene features to predict genes underlying complex traits and diseases. Nat. Genet. 55, 1267–1276 (2023).

59. Lek, M. et al. Analysis of protein-coding genetic variation in 60,706 humans. Nature 536, 285–291 (2016).

60. Boukas, L. et al. Coexpression patterns define epigenetic regulators associated with neurological dysfunction. Genome Res. 29, 532–542 (2019).

61. Georgi, B., Voight, B. F. & Bućan, M. From mouse to human: evolutionary genomics analysis of human orthologs of essential genes. PLoS Genet. 9, e1003484 (2013).

62. Bikker, H. et al. A 20-basepair duplication in the human thyroid peroxidase gene results in a total iodide organification defect and congenital hypothyroidism. J. Clin. Endocrinol. Metab. 79, 248–252 (1994).

63. Bakker, B. et al. Two decades of screening for congenital hypothyroidism in The Netherlands: TPO gene mutations in total iodide organification defects (an update). J. Clin. Endocrinol. Metab. 85, 3708–3712 (2000).

64. Hwangbo, Y. & Park, Y. J. Genome-wide association studies of autoimmune thyroid diseases, thyroid function, and thyroid cancer. Endocrinol. Metab. (Seoul) 33, 175 (2018).

65. Furue, K. et al. The IL -13– OVOL 1– FLG axis in atopic dermatitis. Immunology 158, 281–286 (2019).

66. Sun, P. et al. OVOL1 regulates psoriasis-like skin inflammation and epidermal hyperplasia. J. Invest. Dermatol. 141, 1542–1552 (2021).

67. Dragan, M. et al. Ovol1/2 loss-induced epidermal defects elicit skin immune activation and alter global metabolism. EMBO Rep. 24, (2023).

68. the EArly Genetics and Lifecourse Epidemiology (EAGLE) Eczema Consortium. Multi-ancestry genome-wide association study of 21,000 cases and 95,000 controls identifies new risk loci for atopic dermatitis. Nat. Genet. 47, 1449–1456 (2015).

69. Umans, B. D., Battle, A. & Gilad, Y. Where are the disease-associated eQTLs? Trends Genet. 37, 109–124 (2021).

70. Nachat, R. et al. Peptidylarginine deiminase isoforms are differentially expressed in the anagen hair follicles and other human skin appendages. J. Invest. Dermatol. 125, 34–41 (2005).

71. Zhang, X. et al. Peptidylarginine deiminase 1-catalyzed histone citrullination is essential for early embryo development. Sci. Rep. 6, 1–11 (2016).

72. Revez, J. A. et al. Genome-wide association study identifies 143 loci associated with 25 hydroxyvitamin D concentration. Nat. Commun. 11, 1647 (2020).

73. Manousaki, D. et al. Genome-wide association study for vitamin D levels reveals 69 independent loci. Am. J. Hum. Genet. 106, 327–337 (2020).

74. Ku, H. J., Ahn, Y., Lee, J. H., Park, K. M. & Park, J.-W. IDH2 deficiency promotes mitochondrial dysfunction and cardiac hypertrophy in mice. Free Radic. Biol. Med. 80, 84–92 (2015).

75. Noh, M. R., Kong, M. J., Han, S. J., Kim, J. I. & Park, K. M. Isocitrate dehydrogenase 2 deficiency aggravates prolonged high-fat diet intake-induced hypertension. Redox Biol. 34, 101548 (2020).

76. Karamanavi, E. et al. The FES gene at the 15q26 coronary-artery-disease locus inhibits atherosclerosis. Circ. Res. 131, 1004–1017 (2022).

77. Garimella, P. S., du Toit, C., Le, N. N. & Padmanabhan, S. A genomic deep field view of hypertension. Kidney Int. 103, 42–52 (2023).

78. Curtis, D. Analysis of rare variants in 470,000 exome-sequenced UK Biobank participants implicates novel genes affecting risk of hypertension. Pulse (Basel) 11, 9–16 (2023).

79. de Oliveira, D. F., de Lemos, R. R. & de Oliveira, J. R. M. Mutations at the SLC20A2 gene and brain resilience in families with idiopathic basal ganglia calcification (“Fahr’s disease”). Front. Hum. Neurosci. 7, 420 (2013).

80. Inden, M., Kurita, H. & Hozumi, I. Characteristics and therapeutic potential of sodium-dependent phosphate cotransporters in relation to idiopathic basal ganglia calcification. J. Pharmacol. Sci. 148, 152–155 (2022).

81. Wallingford, M. C. et al. SLC20A2 deficiency in mice leads to elevated phosphate levels in cerbrospinal fluid and glymphatic pathway-associated arteriolar calcification, and recapitulates human idiopathic basal ganglia calcification. Brain Pathol. 27, 64–76 (2017).

82. Sargurupremraj, M. et al. Cerebral small vessel disease genomics and its implications across the lifespan. Nat. Commun. 11, 1–18 (2020).

83. Zhao, B. et al. Transcriptome-wide association analysis of brain structures yields insights into pleiotropy with complex neuropsychiatric traits. Nat. Commun. 12, (2021).

84. Herting, M. M. & Sowell, E. R. Puberty and structural brain development in humans. Front. Neuroendocrinol. 44, 122–137 (2017).

85. Yuan, S. & Larsson, S. C. Inverse association between serum 25-hydroxyvitamin D and nonalcoholic fatty liver disease. Clin. Gastroenterol. Hepatol. 21, 398-405.e4 (2023).

86. Guyenet, P. G. The sympathetic control of blood pressure. Nat. Rev. Neurosci. 7, 335–346 (2006).

87. do Carmo, J. M. et al. Role of the brain melanocortins in blood pressure regulation. Biochim. Biophys. Acta Mol. Basis Dis. 1863, 2508–2514 (2017).

88. Kane, L. & Ismail, N. Puberty as a vulnerable period to the effects of immune challenges: Focus on sex differences. Behav. Brain Res. 320, 374–382 (2017).

89. Resztak, J. A. et al. Analysis of transcriptional changes in the immune system associated with pubertal development in a longitudinal cohort of children with asthma. Nat. Commun. 14, 1–14 (2023).

90. Alberts, B. et al. Lymphocytes and the Cellular Basis of Adaptive Immunity. (Garland Science, London, England, 2002).

91. Lloyd, C. M. & Hessel, E. M. Functions of T cells in asthma: more than just TH2 cells. Nat. Rev. Immunol. 10, 838–848 (2010).

92. Skapenko, A., Leipe, J., Lipsky, P. E. & Schulze-Koops, H. Arthritis Res. 7, S4 (2005).

93. Raphael, I., Joern, R. R. & Forsthuber, T. G. Memory CD4+ T cells in immunity and autoimmune diseases. Cells 9, 531 (2020).

94. Deng, Q. et al. The emerging epigenetic role of CD8+T cells in autoimmune diseases: A systematic review. Front. Immunol. 10, (2019).

95. Collier, J. L., Weiss, S. A., Pauken, K. E., Sen, D. R. & Sharpe, A. H. Not-so-opposite ends of the spectrum: CD8+ T cell dysfunction across chronic infection, cancer and autoimmunity. Nat. Immunol. 22, 809–819 (2021).

96. Liblau, R. S., Wong, F. S., Mars, L. T. & Santamaria, P. Autoreactive CD8 T cells in organ-specific autoimmunity. Immunity 17, 1–6 (2002).

97. Bandala-Sanchez, E. et al. T cell regulation mediated by interaction of soluble CD52 with the inhibitory receptor Siglec-10. Nat. Immunol. 14, 741–748 (2013).

98. Rashidi, M. et al. CD52 inhibits Toll-like receptor activation of NF-κB and triggers apoptosis to suppress inflammation. Cell Death Differ. 25, 392–405 (2018).

99. Bhamidipati, K. et al. CD52 is elevated on B cells of SLE patients and regulates B cell function. Front. Immunol. 11, (2021).

100. Zhou, M. et al. Krüppel-like transcription factor 13 regulates T lymphocyte survival in vivo. J. Immunol. 178, 5496–5504 (2007).

101. Outram, S. V. et al. KLF13 influences multiple stages of both B and T cell development. Cell Cycle 7, 2047–2055 (2008).

102. Walker, L. S. K. & Sansom, D. M. The emerging role of CTLA4 as a cell-extrinsic regulator of T cell responses. Nat. Rev. Immunol. 11, 852–863 (2011).

103. Oyewole-Said, D. et al. Beyond T-cells: Functional characterization of CTLA-4 expression in immune and non-immune cell types. Front. Immunol. 11, (2020).

104. Hosseini, A., Gharibi, T., Marofi, F., Babaloo, Z. & Baradaran, B. CTLA-4: From mechanism to autoimmune therapy. Int. Immunopharmacol. 80, 106221 (2020).

105. Hossen, M. M. et al. Current understanding of CTLA-4: from mechanism to autoimmune diseases. Front. Immunol. 14, (2023).

106. Pedicord, V. A., Montalvo, W., Leiner, I. M. & Allison, J. P. Single dose of anti– CTLA-4 enhances CD8 + T-cell memory formation, function, and maintenance. Proc. Natl. Acad. Sci. U. S. A. 108, 266–271 (2011).

107. Chan, D. V. et al. Differential CTLA-4 expression in human CD4+ versus CD8+ T cells is associated with increased NFAT1 and inhibition of CD4+ proliferation. Genes Immun. 15, 25–32 (2014).

108. Ueda, H. et al. Association of the T-cell regulatory gene CTLA4 with susceptibility to autoimmune disease. Nature 423, 506–511 (2003).

109. Wang, K. et al. CTLA-4 +49 G/A polymorphism confers autoimmune disease risk: An updated meta-analysis. Genet. Test. Mol. Biomarkers 21, 222–227 (2017).

110. Valdés-Ferrer, S. I. et al. HMGB1 mediates anemia of inflammation in Murine sepsis survivors. Mol. Med. 21, 951–958 (2015).

111. Dulmovits, B. M. et al. HMGB1-mediated restriction of EPO signaling contributes to anemia of inflammation. Blood 139, 3181–3193 (2022).

112. Liu, L. et al. Conditional transcriptome-wide association study for fine-mapping candidate causal genes. Nat. Genet. 56, 348–356 (2024).

113. Benner, C. et al. Prospects of fine-mapping trait-associated genomic regions by using summary statistics from genome-wide association studies. Am. J. Hum. Genet. 101, 539–551 (2017).

114. Cuomo, A. S. E., Nathan, A., Raychaudhuri, S., MacArthur, D. G. & Powell, J. E. Single-cell genomics meets human genetics. Nat. Rev. Genet. 24, 535–549 (2023).

115. Võsa, U. et al. Large-scale cis- and trans-eQTL analyses identify thousands of genetic loci and polygenic scores that regulate blood gene expression. Nat. Genet. 53, 1300–1310 (2021).

116. Jansen, R. et al. Conditional eQTL analysis reveals allelic heterogeneity of gene expression. Hum. Mol. Genet. 26, 1444–1451 (2017).

117. Liu, C. et al. Whole genome DNA and RNA sequencing of whole blood elucidates the genetic architecture of gene expression underlying a wide range of diseases. Sci. Rep. 12, 1–11 (2022).

118. Zhang, Y., Qi, G.Park, J.-H. & Chatterjee, N. Estimation of complex effect-size distributions using summary-level statistics from genome-wide association studies across 32 complex traits. Nat. Genet. 50, 1318–1326 (2018).

119. O’Connor, L. J. The distribution of common-variant effect sizes. Nat. Genet. 53, 1243–1249 (2021).

120. Yengo, L. et al. A saturated map of common genetic variants associated with human height. Nature 610, 704–712 (2022).

121. Bhattacharya, A. et al. Best practices for multi-ancestry, meta-analytic transcriptome-wide association studies: Lessons from the Global Biobank Meta-analysis Initiative. Cell Genom. 2, 100180 (2022).

122. Kachuri, L. et al. Gene expression in African Americans, Puerto Ricans and Mexican Americans reveals ancestry-specific patterns of genetic architecture. Nat. Genet. 55, 952–963 (2023).

123. Zaitlen, N., Paşaniuc, B., Gur, T., Ziv, E. & Halperin, E. Leveraging genetic variability across populations for the identification of causal variants. Am. J. Hum. Genet. 86, 23–33 (2010).

124. Kichaev, G. & Pasaniuc, B. Leveraging functional-annotation data in trans-ethnic fine-mapping studies. Am. J. Hum. Genet. 97, 260–271 (2015).

125. Lu, Z. et al. Multi-ancestry fine-mapping improves precision to identify causal genes in transcriptome-wide association studies. Am. J. Hum. Genet. 109, 1388–1404 (2022).

126. Kichaev, G. et al. Integrating functional data to prioritize causal variants in statistical fine-mapping studies. PLoS Genet. 10, e1004722 (2014).

127. McVicker, G. et al. Identification of genetic variants that affect histone modifications in human cells. Science 342, 747–749 (2013).

128. Li, Y. I. et al. RNA splicing is a primary link between genetic variation and disease. Science 352, 600–604 (2016).

129. Xu, Y. et al. An atlas of genetic scores to predict multi-omic traits. Nature 616, 123–131 (2023).

130. Sun, B. B. et al. Plasma proteomic associations with genetics and health in the UK Biobank. Nature 622, 329–338 (2023).

131. Hou, L. et al. Multitissue H3K27ac profiling of GTEx samples links epigenomic variation to disease. Nat. Genet. 55, 1665–1676 (2023).

132. Oliva, M. et al. DNA methylation QTL mapping across diverse human tissues provides molecular links between genetic variation and complex traits. Nat. Genet. 55, 112–122 (2023).

133. Zhou, X., Carbonetto, P. & Stephens, M. Polygenic modeling with Bayesian sparse linear mixed models. PLoS Genet. 9, e1003264 (2013).

134. Lloyd-Jones, L. R. et al. Improved polygenic prediction by Bayesian multiple regression on summary statistics. Nat. Commun. 10, 1–11 (2019).

135. Privé, F., Arbel, J. & Vilhjálmsson, B. J. LDpred2: better, faster, stronger. Bioinformatics 36, 5424–5431 (2021).

136. Marriott, P., Efron, B. & Tibshirani, R. J. An introduction to the bootstrap. J. R. Stat. Soc. Ser. A Stat. Soc. 158, 347 (1995).

137. Benjamini, Y. & Hochberg, Y. Controlling the false discovery rate: A practical and powerful approach to multiple testing. J. R. Stat. Soc. 57, 289–300 (1995).

138. Yuan, K. et al. Fine-mapping across diverse ancestries drives the discovery of putative causal variants underlying human complex traits and diseases. bioRxiv 2023.01.07.23284293 (2023) doi:10.1101/2023.01.07.23284293.

139. Loh, P.-R. et al. Efficient Bayesian mixed-model analysis increases association power in large cohorts. Nat. Genet. 47, 284–290 (2015).

140. Kent, W. J. et al. The human genome browser at UCSC. Genome Res. 12, 996–1006 (2002).

141. Robinson, M. D., McCarthy, D. J. & Smyth, G. K. edgeR: a Bioconductor package for differential expression analysis of digital gene expression data. Bioinformatics 26, 139–140 (2010).

