## Supplementary Material for "Fine-mapping causal tissues and genes at disease-associated loci"

*Supplementary Table Captions*

**Supplementary Table 1: Numerical data underlying Figure 1a-b.** For each (tissue-gene fine-mapping method, eQTL sample size, PIP threshold) triplet, we report the average empirical False Discovery Rate (FDR) across 100 default simulations, as well as the 95% confidence interval lower and upper bounds on the average empirical FDR. We consider the tissue-gene fine-mapping methods TGFM (Gene-Tissue), FOCUS-TG, FOCUS, and coloc.

**Supplementary Table 2: Numerical data underlying Figure 1c-d.** For each (tissue-gene fine-mapping method, eQTL sample size, PIP threshold) triplet, we report the average power for detecting causal gene-tissue pairs across 100 default simulations, as well as the 95% confidence interval lower and upper bounds on the average power. We consider the tissue-gene fine-mapping methods TGFM (Gene-Tissue), FOCUS-TG, FOCUS, and coloc.

**Supplementary Table 3: Numerical data underlying Figure 2a-b.** For each (genetic element, eQTL sample size, PIP threshold) triplet, we report the average empirical False Discovery Rate (FDR) across 100 default simulations, as well as the 95% confidence interval lower and upper bounds on the average empirical FDR. Genetic elements include gene-tissue pairs, genes, and non-mediated genetic variants.

**Supplementary Table 4: Numerical data underlying Figure 2c-d.** For each (genetic element, eQTL sample size, PIP threshold) triplet, we report the average power across 100 default simulations, as well as the 95% confidence interval lower and upper bounds on the average power. Genetic elements include gene-tissue pairs, genes, and non-mediated genetic variants.

**Supplementary Table 5: Description of 45 analyzed UK Biobank diseases and traits.** For each of the 45 analyzed UK Biobank diseases and complex traits, we report the study name, the study sample size, and study's estimated heritability.

**Supplementary Table 6: Description of GTEx tissues.** For each of the 38 GTEx metatissues we report the metatissue sample size, the composite tissues defining the metatissue (','-seperated if multiple composite tissues), the sample size of each composite tissue (','-seperated if multiple composite tissues), and the number of genes with a gene model in the metatissue.

**Supplementary Table 7: TGFM runtimes for UKBB traits.** We split TGFM into 3 tasks. For each of the three tasks, we report the total TGFM runtime in hours. The first task is running TGFM step 1 and TGFM step 2 (in TGFM step 1, we apply SuSiE to perform eQTL fine-mapping of each gene-tissue pair in the external gene expression data set (estimating a posterior distribution of the causal cis-eQTL effect sizes for each gene-tissue pair). In TGFM step 2, we randomly sample 100 cis-predicted expression models for each gene-tissue pair from the posterior distributions of causal cis-eQTL effect sizes estimated in TGFM step 1). We report the runtime of the first task for a single, average sample size GTEx tissue: Heart Left Ventricle. We note that this task can be parallelized across genes (i.e., each gene can be run independently). The second task is running the TGFM tissue specific prior. We report the runtime of the second task for a single, average heritability trait: Total cholesterol. We note that this task can be parallelized across fine-mapping windows (i.e., each fine-mapping window can be run independently). The third task is running TGFM step 3 and TGFM step 4 (In TGFM step 3, we apply SuSiE to perform disease fine-mapping in the target data set (estimating the PIP of each genetic element) 100 times, iterating over the sampled cis-predicted expression models for each gene-tissue pair from TGFM step 2. In TGFM step 4, we average the results of TGFM step 3 across the 100 disease fine-mapping runs.). We report the runtime of the third task for a single, average heritability trait: Total cholesterol. We note that this task can be parallelized across fine-mapping windows (i.e., each fine-mapping window can be run independently). Runtimes do not include computing LD matrices, as that is considered part of the input to TGFM (see data availability).

**Supplementary Table 8: Number of fine-mapped genetic elements for UK Biobank diseases and traits.** For each trait-genetic element pair with TGFM PIP ≥ 0.2 corresponding to one of the 16 independent traits (Figure 3), we report the corresponding genetic element class (ie. gene-tissue, gene, or variant), the trait, and the TGFM PIP.

**Supplementary Table 9: Proportion of fine-mapped gene-tissue pairs in each tissue for UK Biobank diseases and traits.** For each trait-tissue pair, we report the proportion of the trait's fine-mapped gene-tissue pairs in the tissue (proportions for each trait were calculated by counting the number of gene-tissue pairs with TGFM PIP > 0.5 in each tissue and normalizing the counts across tissues), the p-value of the trait-tissue pair determined by applying genomic bootstrapping to the TGFM tissue-specific prior, and the FDR significance (calculated by applying Benjamini-Hochberg correction for 38 tissues; '**' represents FDR ≤ 0.05, '**' represents FDR ≤ 0.2, 'null' denotes FDR > 0.2).

**Supplementary Table 10: Matched cell-type groups of GTEx tissues.** For each of the 38 GTEx tissues, we report the matched cell-type group used in S-LDSC analyses with chromatin data.

**Supplementary Table 11: Numerical data underlying Figure 4b.** For each trait-tissue pair, we report the matched cell type group name used in S-LDSC chromatin analysis, the number corresponding to that cell type group, a binary variable corresponding to significance of the TGFM tissue-specific prior (FDR ≤ 0.05), the p-value corresponding to significance of the TGFM tissue-specific prior, a z-score from S-LDSC corresponding to significance of the chromatin annotation from the matched cell type group on the heritability of the trait, and a one-sided p-value corresponding to the significance of the chromatin annotation from the matched cell type group on the heritability of the trait.

**Supplementary Table 12: Average correlation in cis-predicted gene expression between pairs of GTEx tissues.** For each pair of tissues, we report the average correlation in cis-predicted gene expression across genes (we only consider genes with a cis-predicted expression model in both tissues) and the number of genes the average is taken across. We observed that several of the low sample size tissues (e.g. uterus, vagina, artery coronary, minor salivary gland, prostate; Supplementary Table 6) have high correlation with nearly all other tissues. We assume this is a technical artifact of low sample size tissues being limited to discovering very large eQTL effect size variants (due to statistical power), which are known to be less tissue-specific than small eQTL effect size variants.

**Supplementary Table 13: Average PoPS score of genes stratified by TGFM (Gene) PIP.** For each TGFM (Gene) PIP bin (we use the following bins: $0 \leq PIP < 0.01$, $0.01 \leq PIP < 0.25$, $0.25 \leq PIP < 0.5$, $0.5 \leq PIP < 0.7$, $0.7 \leq PIP < 0.9$, and $0.9 \leq PIP$), we report the average PoPS score of all gene-trait pairs in the bin across 16 independent traits, and the standard error of the average PoPS score.

**Supplementary Table 14: Enrichment of fine-mapped TGFM genes within non-disease-specific gene sets.** For each (non-disease-specific gene set, TGFM PIP threshold) pair, we report enrichment statistics corresponding to enrichment of genes with TGFM (Gene) PIP > TGFM PIP threshold within the non-disease-specific gene set meta-analyzed over 16 independent traits. Enrichment statistics consist of enrichment odds ratio, enrichment odds ratio 95% confidence interval lower bound, enrichment odds ratio 95% confidence interval upper bound, and enrichment p-value. Odds ratios and standard errors on the odds ratio were computed using logistic regression.

**Supplementary Table 15. Numerical data underlying Figure 4d.** For each TGFM (Gene) PIP threshold, we report the empirical FDR according to the 69 silver standard LDL genes, along with 95% confidence interval lower and upper bound on the empirical FDR, as well as the (1 – average PIP) at the PIP threshold.

**Supplementary Table 16. Numerical data underlying Figure 5a.** We report the number of loci in each loci category resulting from the tissue ablation analysis.

**Supplementary Table 17. Numerical data underlying Figure 5b.** For each GTEx tissue, we report the average PIP in that tissue/cell type across all gene-tissue pairs that were fine-mapped in GTEx whole blood (PIP > 0.5) in the primary analysis. Other than PBMC, the tissue with the second highest average PIP was spleen, a proxy tissue of whole blood.

**Supplementary Table 18: Numerical data underlying examples of fine-mapped gene-tissue-disease triplets identified by TGFM.** For each genetic element-disease pair visualized in Figure 6, we report the -log10(p-value) of the marginal association between the genetic element and disease (we report the median -log10(TWAS p-value) across the 100 sets of sampled cis-predicted expression for gene-tissue pairs and the -log10(GWAS p-value) for variants), the location of the genetic element (we report the gene's TSS for gene-tissue pairs and the variant location for variants), and the TGFM PIP of the genetic element.

**Supplementary Table 19: Description of PBMC cell types from Perez et al. Science 2022.** For each of the 9 PBMC cell types we report the sample size, the average number of cells per donor, and the number of genes with a gene model.

**Supplementary Table 20: Number of fine-mapped gene-PBMC cell type pairs for all 45 UK Biobank diseases and traits.** For each gene-PBMC cell type-trait triplets with TGFM PIP $\geq 0.2$ corresponding to one of the 45 analyzed UKBB diseases and complex traits, we report the corresponding trait, PBMC cell type, and the TGFM PIP.

**Supplementary Table 21: Significance of tissue-specific prior for each trait-tissue pair and each trait-PBMC cell type pair.** For each trait-tissue pair and each trait-PBMC cell type pair, we report the p-value of the trait-tissue pair or the trait-PBMC cell type pair determined by applying genomic bootstrapping to the TGFM tissue-specific prior, and the FDR significance (calculated by applying Benjamini-Hochberg correction for 38 tissues and 9 cell types; '**' represents FDR $\leq$ 0.05, '**' represents FDR $\leq$ 0.2, 'null' denotes FDR $>$ 0.2).

**Supplementary Table 22: Numerical data underlying examples of fine-mapped gene-PBMC cell type-disease triplets identified by TGFM.** For each genetic element-disease pair visualized in Figure 8, we report the -log10(p-value) of the marginal association between the genetic element and disease (we report the median -log10(TWAS p-value) across the 100 sets of sampled cis-predicted expression for gene-tissue pairs and gene-PBMC cell type pairs and the -log10(GWAS p-value) for variants), the location of the genetic element (we report the gene's TSS for gene-tissue pairs and gene-PBMC cell type pairs, and the variant location for variants), and the TGFM PIP of the genetic element.

### ***Supplementary Note***

#### Additional examples of fine-mapped gene-tissue pairs

We highlight 6 additional examples of fine-mapped (PIP > 0.5) gene-tissue-trait triplets that recapitulate known biology or nominate biologically plausible mechanisms. First, TGFM fine-mapped *LIPC* in liver for Vitamin D level (Supplementary Figure 41a; gene-tissue PIP: 0.83; gene PIP: 0.87). The involvement of *LIPC* in lipid metabolism is well-studied[1,2], and other work has demonstrated the impact of lipid biology on Vitamin D levels; Vitamin D is a fat-soluble hormone[3,4]. *LIPC* has previously been linked to Vitamin

D level in genetic association studies[3,5]. Liver was also identified as a Vitamin D level-critical tissue genome-wide (proportion of fine-mapped gene-tissue pairs = 0.14, bootstrap p = 0.02 for tissue-specific prior; Supplementary Figure 31). We note that we have highlighted two fine-mapped gene-tissue pairs for Vitamin D level (Figure 6c and Supplementary Figure 41a) involving two different tissues (skin (sun exposed) and liver); this demonstrates the advantages of TGFM over a two-step approach of separately identifying the causal gene using a gene-level fine-mapping[6] and identifying the causal tissue using a method for identifying trait-critical tissues[7–9].

Second, TGFM fine-mapped *MC4R* in brain cerebellum for Systolic blood pressure (Supplementary Figure 41b, gene-tissue PIP: 0.66, gene PIP: 0.66), despite a non-significant TWAS p-value ($p = 3.3 \times 10^{-5} > 0.05/119{,}270 = 4.2 \times 10^{-7}$). Previous work has shown that activation of *MC4R* in the central nervous system increases sympathetic nervous system activity and blood pressure[10–12], and *MC4R* has previously been linked to hypertension in genetic association studies[13]. Brain cerebellum was identified as a Systolic blood pressure-critical tissue genome-wide (proportion of fine-mapped gene-tissue pairs = 0.16, bootstrap p = 0.002 for tissue-specific prior; Supplementary Figure 31), consistent with previous studies[11,14,15]. Strong support of brain cerebellum genes from the tissue-specific prior enabled TGFM to prioritize *MC4R*-brain cerebellum instead of more significant nearby GWAS associations of non-mediated genetic variants. We note that we have highlighted two fine-mapped gene-tissue pairs for Systolic blood pressure (Figure 6d and Supplementary Figure 41b) involving two different tissues (artery aorta and brain cerebellum); this demonstrates the advantages of TGFM over a two-step approach of separately identifying the causal gene using a gene-level fine-mapping[6] and identifying the causal tissue using a method for identifying trait-critical tissues[7–9].

Third, TGFM fine-mapped *SMIM1* in whole blood for Red blood cell count (Supplementary Figure 41c; gene-tissue PIP: 0.78; gene PIP: 0.84). *SMIM1* was previously reported to encode the Vel blood group protein involved in red blood cell formation[16,17], and has previously been linked to Red blood cell count in genetic association studies[18,19]. Whole blood was also identified as a Red blood cell count-critical tissue genome-wide (proportion of fine-mapped gene-tissue pairs = 0.23, bootstrap p = 0.01 for tissue-specific prior; Supplementary Figure 31), an intuitive finding given that red blood cells constitute a large proportion of whole blood tissue. There exist 25 other gene-tissue within 1 Mb of the TSS of *SMIM1* (2 of which correspond to *SMIM1* in a tissue other than whole blood) that had significant TWAS p-values ($p \leq 0.05 / 119{,}270 = 4.2 \times 10^{-7}$) but were not fine-mapped by TGFM (all with PIP $\leq$ 0.01), underscoring the benefit of joint fine-mapping of gene-tissue pairs. TGFM also fine-mapped one non-mediated variant (rs1569419; PIP: 1.0) within 1 Mb of the TSS of *SMIM1*, perhaps due to finite eQTL sample size and/or absence of the causal cell-type or context in GTEx expression data (see Discussion).

Fourth, TGFM fine-mapped *NYNRIN* in liver for Total cholesterol (Supplementary Figure 41d; gene-tissue PIP: 0.70; gene PIP: 0.98). *NYNRIN* has previously been linked to LDL cholesterol in genetic association studies[20,21]. Liver was also identified as a Total cholesterol-critical tissue genome-wide (proportion of fine-mapped gene-tissue pairs = 0.60, bootstrap $p = 2.0 \times 10^{-6}$ for tissue-specific prior; Supplementary Figure 31). There exist 2 other gene-tissue within 1 Mb of the TSS of *NYNRIN* that had significant TWAS p-values ($p \leq 0.05 / 119{,}270 = 4.2 \times 10^{-7}$) but were not fine-mapped by TGFM (both with PIP $\leq 0.01$), underscoring the benefit of joint fine-mapping of gene-tissue pairs.

Fifth, TGFM fine-mapped *NRP2* in lung for FEV1:FVC (ratio of forced expiratory volume in 1 second to forced vital capacity; Supplementary Figure 41e; gene-tissue PIP: 0.98; gene PIP: 0.99). Recent work has demonstrated the role of *NRP2* in regulating airway inflammatory responses in the lungs[22,23], but *NRP2* has not previously been linked to FEV1:FVC to our knowledge. Lung was also identified as a FEV1:FVC-critical tissue genome-wide (proportion of fine-mapped gene-tissue pairs = 0.13, bootstrap p = 0.002 for tissue-specific prior; Supplementary Figure 31).

Sixth, TGFM fine-mapped *SIX3* in pancreas for HbA1c levels (Supplementary Figure 41f; gene-tissue PIP: 0.67; gene PIP: 0.67). Previous work has demonstrated that *SIX3* regulates the functional maturity of human pancreatic $\beta$ cells[24,25], but *SIX3* has not previously been linked to *HbA1c* levels in genetic association studies to our knowledge. Pancreas was suggestively implicated as a HbA1c-critical tissue genome-wide (proportion of fine-mapped gene-tissue pairs = 0.13, bootstrap p = 0.06 for tissue-specific prior; Supplementary Figure 31).

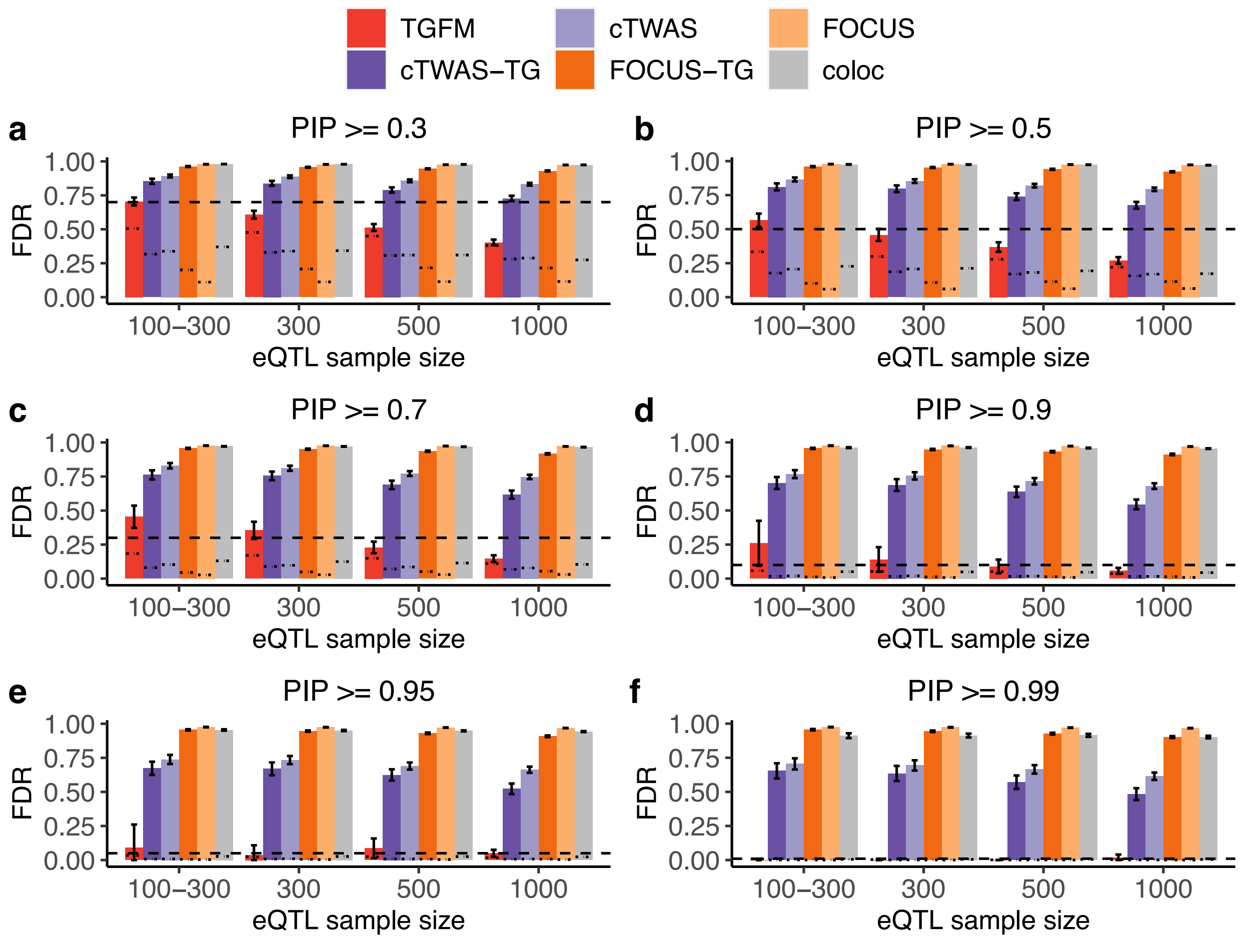


**Supplementary Figure 1: Calibration of tissue-gene fine-mapping methods at various PIP thresholds in simulations.** Average gene-tissue pair fine-mapping FDR across 100 simulations for various fine-mapping methods (see legend) across eQTL sample sizes (x-axis) at PIP=0.3 **(a)**, PIP=0.5 **(b)**, PIP=0.7 **(c)**, PIP=0.9 **(d)**, PIP=0.95 **(e)**, and PIP=0.99 **(f)**. In each plot, the single thick-dashed horizontal line (1 – PIP) threshold (see main text). The thin dashed lines specific to each bar denotes (1 – average PIP) (where average is taken across all genetic elements belonging to that bar; see main text). Error bars denote 95% confidence intervals. This supplementary figure is similar to Fig. 1a-b, except it includes more PIP thresholds, as well as dashed lines denoting (1 – average PIP).

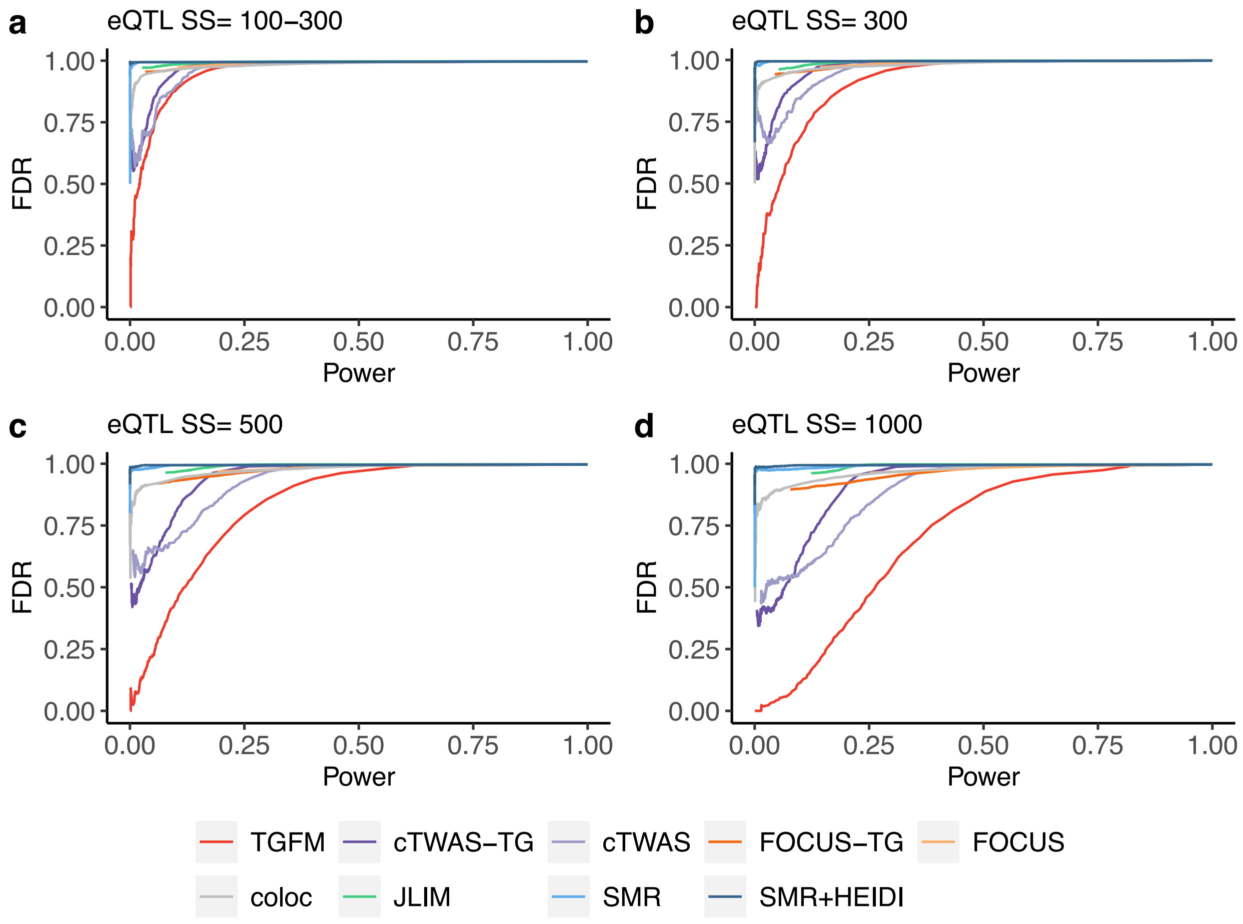


**Supplementary Figure 2: Comparison of tissue-gene fine-mapping power at same level of FDR in simulations.** Average gene-tissue fine-mapping power (x-axis) at a specific level of FDR (y-axis) across 100 simulations for various fine-mapping methods (see legend) at eQTL sample size 100-300 **(a)**, 300 **(b)**, 500 **(c)**, and 1000 **(d).** We note that all methods other than TGFM (cTWAS-TG, cTWAS, FOCUS-TG, FOCUS, coloc, JLIM, SMR, and SMR+HEIDI) are severely mis-calibrated, with high FDR at even the most stringent p-value or posterior probability thresholds, as evident by no method other than TGFM achieving an FDR <= 0.34 at any threshold. JLIM, SMR, and SMR+HEIDI compute p-values for each gene-tissue pair, whereas TGFM, cTWAS-TG, cTWAS, FOCUS-TG, FOCUS, and coloc calculate posterior probabilities for each gene-tissue pair. SMR corresponds to using the SMR p-value to assess the significance of a gene-tissue pair, whereas SMR+HEIDI corresponds to using the SMR p-value to assess the significance of a gene-tissue pair after filtering to gene-tissue pairs with HEIDI p-value <= 0.05. We do not visualize FDR and power of any p-value or posterior probability threshold containing fewer than 2 gene-tissue pairs in order to remove highly uncertain FDR and power estimates from the visualization.

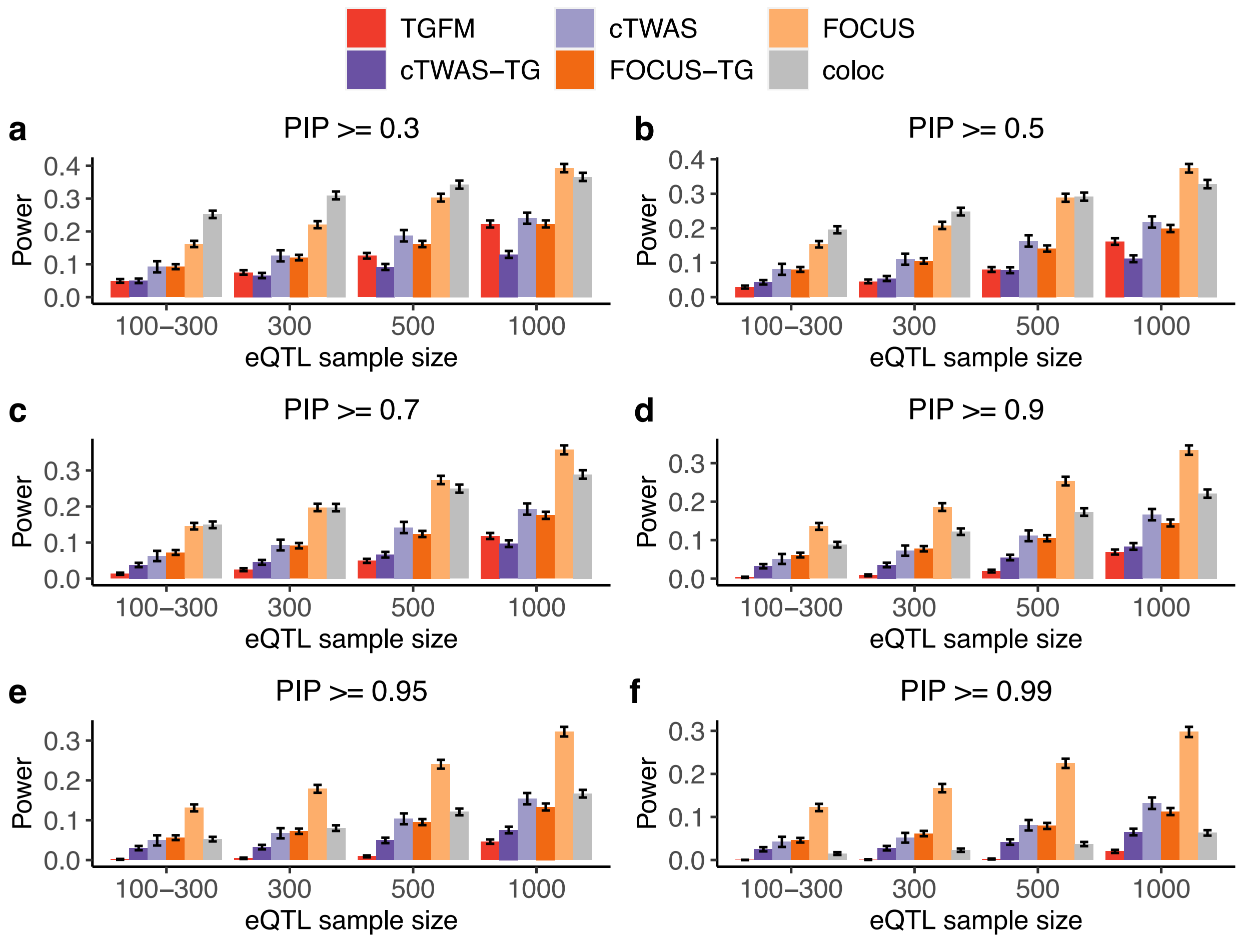


**Supplementary Figure 3: Power of tissue-gene fine-mapping methods at various PIP thresholds in simulations.** Average gene-tissue pair fine-mapping power across 100 simulations for various fine-mapping methods (see legend) across eQTL sample sizes (x-axis) at PIP=0.3 **(a)**, PIP=0.5 **(b)**, PIP=0.7 **(c)**, PIP=0.9 **(d)**, PIP=0.95 **(e)**, and PIP=0.99 **(f)**. Error bars denote 95% confidence intervals. This supplementary figure is similar to Fig. 1c-d, except it includes more PIP thresholds.

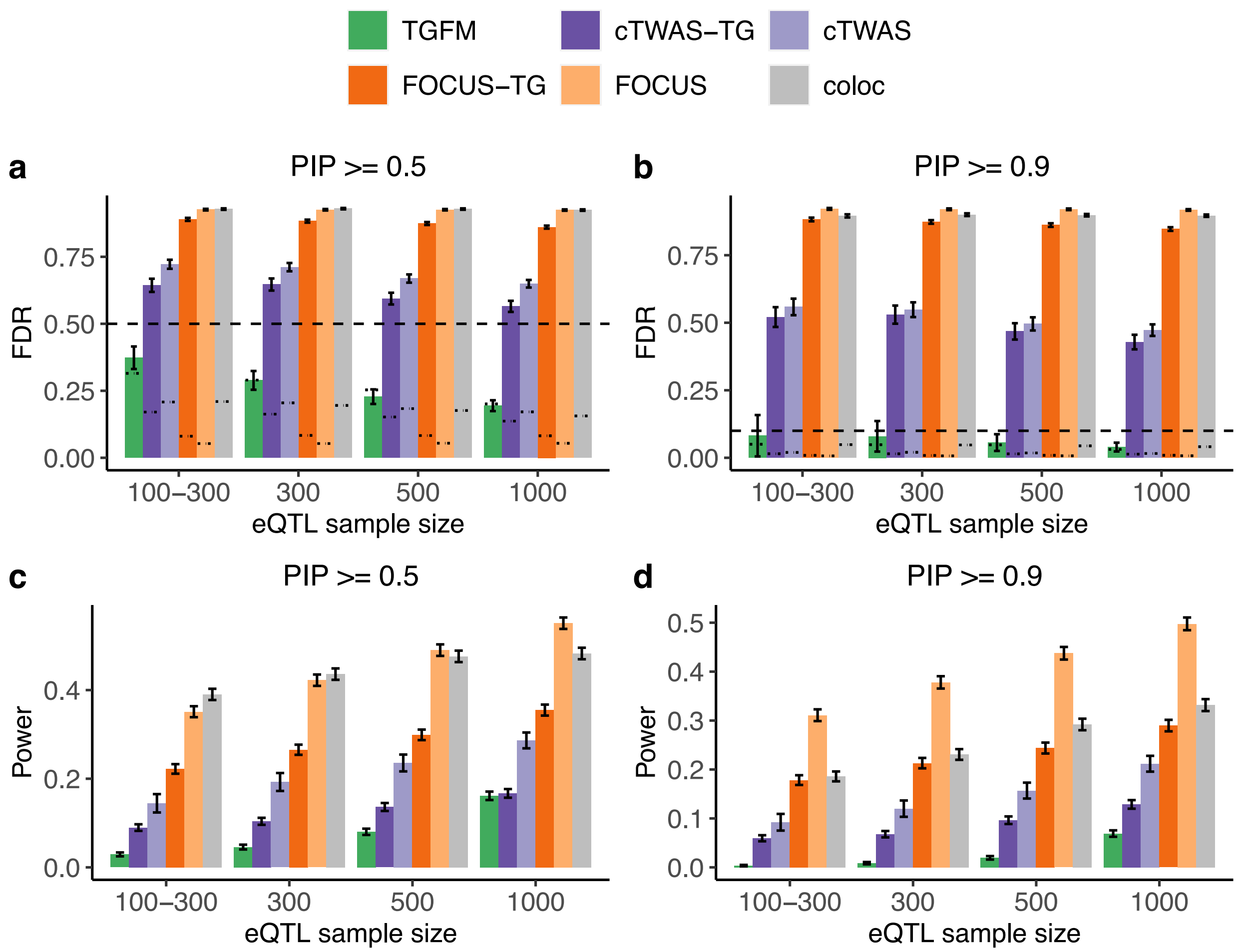


**Supplementary Figure 4: Calibration and power of gene fine-mapping methods at various PIP thresholds in simulations. (a,b)** Average *gene* fine-mapping FDR across 100 simulations for various fine-mapping methods (see legend) across eQTL sample sizes (x-axis) at PIP=0.5 **(a)** and PIP=0.9 **(b)**. Single thick, dashed horizontal line denotes 1 – PIP threshold (see main text). The thin dashed lines specific to each bar denotes (1 – average PIP) (where average is taken across all genetic elements belonging to that bar; see main text). **(c,d)** Average gene fine-mapping power across 100 simulations for various fine-mapping methods (see legend) across eQTL sample sizes (x-axis) at PIP=0.5 **(c)** and PIP=0.9 **(d)**. Error bars denote 95% confidence intervals. This supplementary figure is similar to Figure 1, except it shows calibration and power of gene fine-mapping, instead of tissue-gene fine-mapping. “TGFM” corresponds to TGFM (Gene). “cTWAS-TG” corresponds to running cTWAS applied to the task of tissue-gene fine-mapping, and then summing PIPs across tissues. “cTWAS” corresponds to running cTWAS in each tissue, independently, and taking the maximum cTWAS PIP across tissues for each gene. “FOCUS-TG” corresponds to running FOCUS applied to the task of tissue-gene fine-mapping, and then summing PIPs across tissues. “FOCUS” corresponds to running FOCUS in each tissue, independently, and taking the maximum FOCUS PIP across tissues for each gene. “coloc” corresponds to running coloc in each tissue, independently, and taking the maximum coloc PPH4 across tissues for each gene.

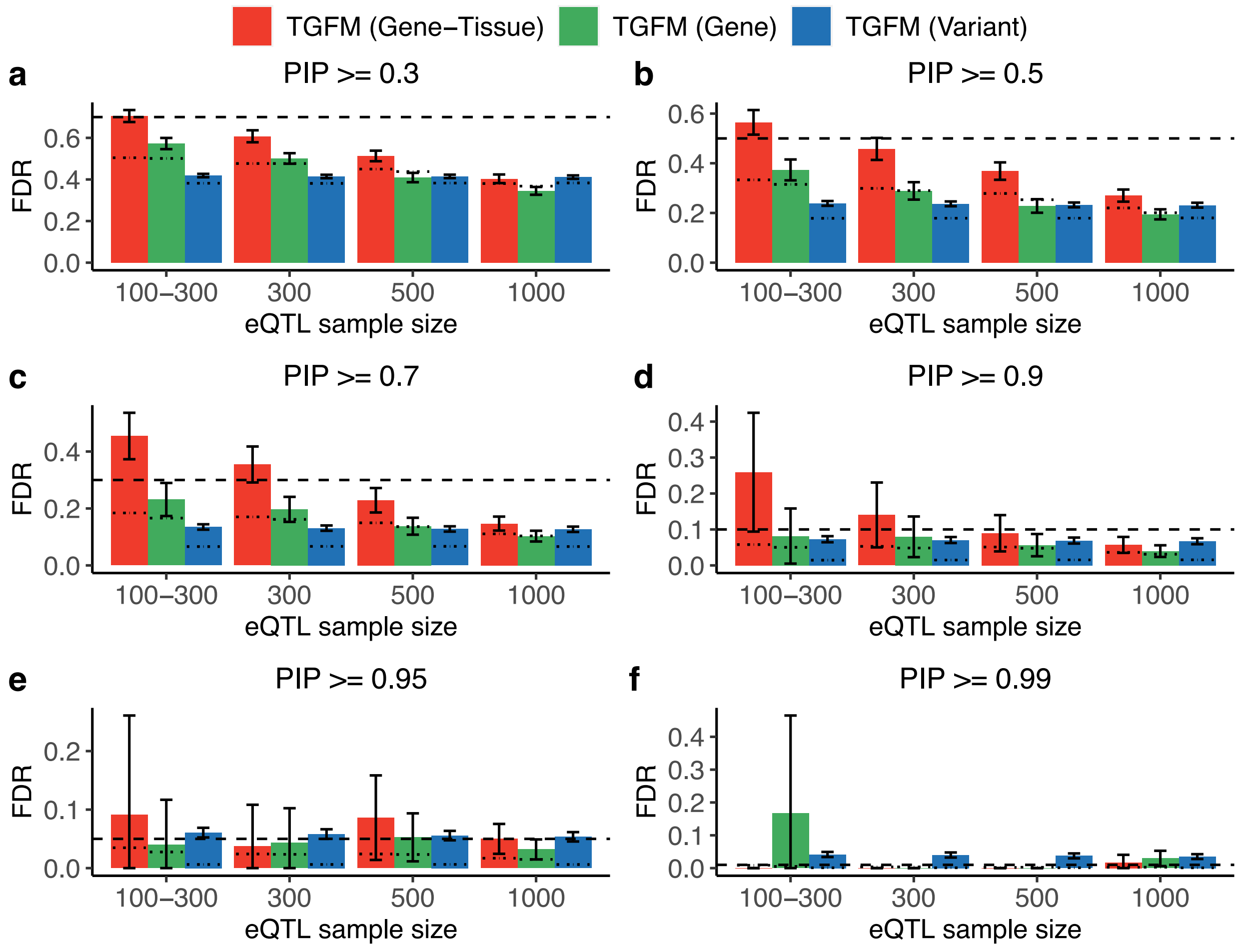


**Supplementary Figure 5: Calibration of fine-mapping genetic elements with TGFM at various PIP thresholds in simulations. (a,b)** Average fine-mapping FDR across 100 simulations using TGFM for different classes of genetic elements (see legend) across eQTL sample sizes (x-axis) at PIP=0.3 **(a)**, PIP=0.5 **(b)**, PIP=0.7 **(c)**, PIP=0.9 **(d)**, PIP=0.95 **(e)**, and PIP=0.99 **(f)**. In each plot, the single thick-dashed horizontal line (1 – PIP) threshold (see main text). The thin dashed lines specific to each bar denotes (1 – average PIP) (where average is taken across all genetic elements belonging to that bar; see main text). Error bars denote 95% confidence intervals. This supplementary figure is similar to Fig. 2a-b, except it includes more PIP thresholds, as well as dashed lines denoting (1 – average PIP).

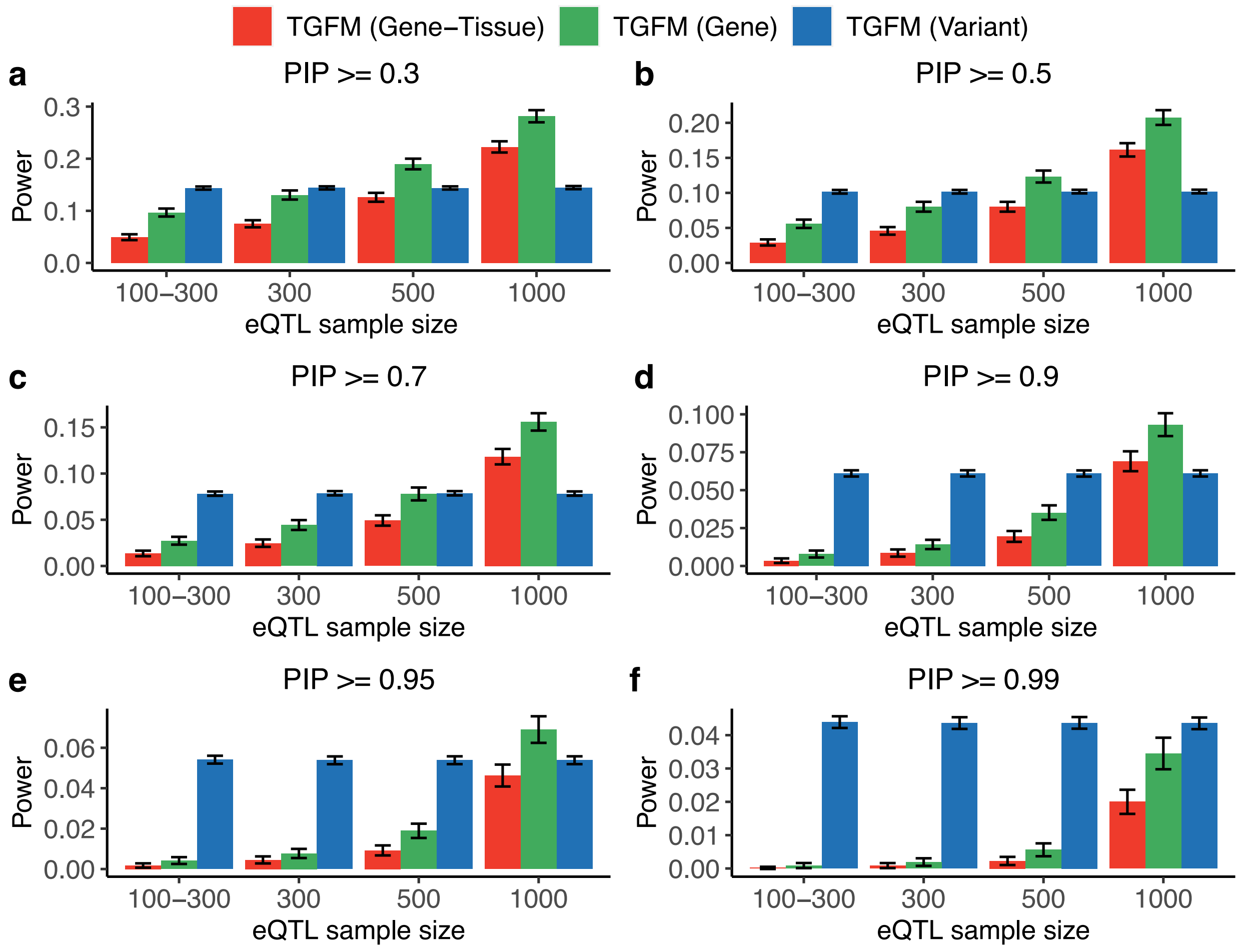


**Supplementary Figure 6: Power of fine-mapping genetic elements with TGFM at various PIP thresholds in simulations.** Average fine-mapping power across 100 simulations using TGFM for different classes of genetic elements (see legend) across eQTL sample sizes (x-axis) at PIP=0.3 **(a)**, PIP=0.5 **(b)**, PIP=0.7 **(c)**, PIP=0.9 **(d)**, PIP=0.95 **(e)**, and PIP=0.99 **(f)**. Error bars denote 95% confidence intervals. This supplementary figure is similar to Fig. 2c-d, except it includes more PIP thresholds.

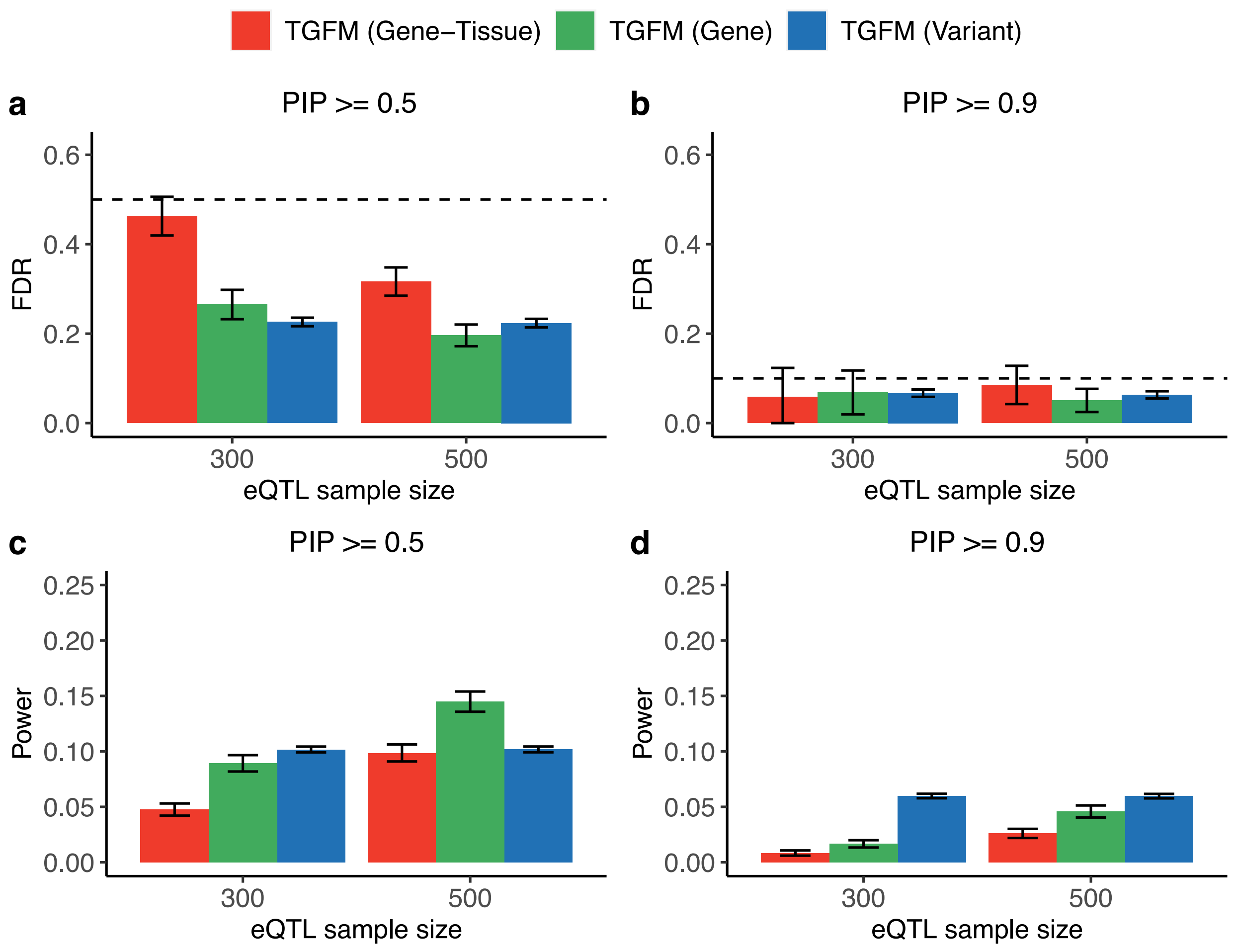


**Supplementary Figure 7: Calibration and power of fine-mapping in simulation setting where the number of causal eQTLs is varied across gene-tissue pairs.** In this simulation setting, the number of causal eQTLs for a given gene-tissue pairs was randomly selected to be between 3 and 7 (inclusive), instead of fixed at 5 as in our primary simulations. **(a,b)** Average fine-mapping FDR across 100 simulations using TGFM for different classes of genetic elements (see legend) across eQTL sample sizes (x-axis) at PIP=0.5 **(a)** and PIP=0.9 **(b)**. Dashed horizontal line denotes 1 – PIP threshold (see main text). **(c,d)** Average fine-mapping power across 100 simulations using TGFM for different classes of genetic elements (see legend) across *eQTL* sample sizes (x-axis) at PIP=0.5 **(c)** and PIP=0.9 **(d)**. Error bars denote 95% confidence intervals. This supplementary figure is similar to Figure 2, except it only shows simulation results at eQTL sample size of 300 and 500, *and* it based on data from an alternative simulation setting described at the beginning of the caption.

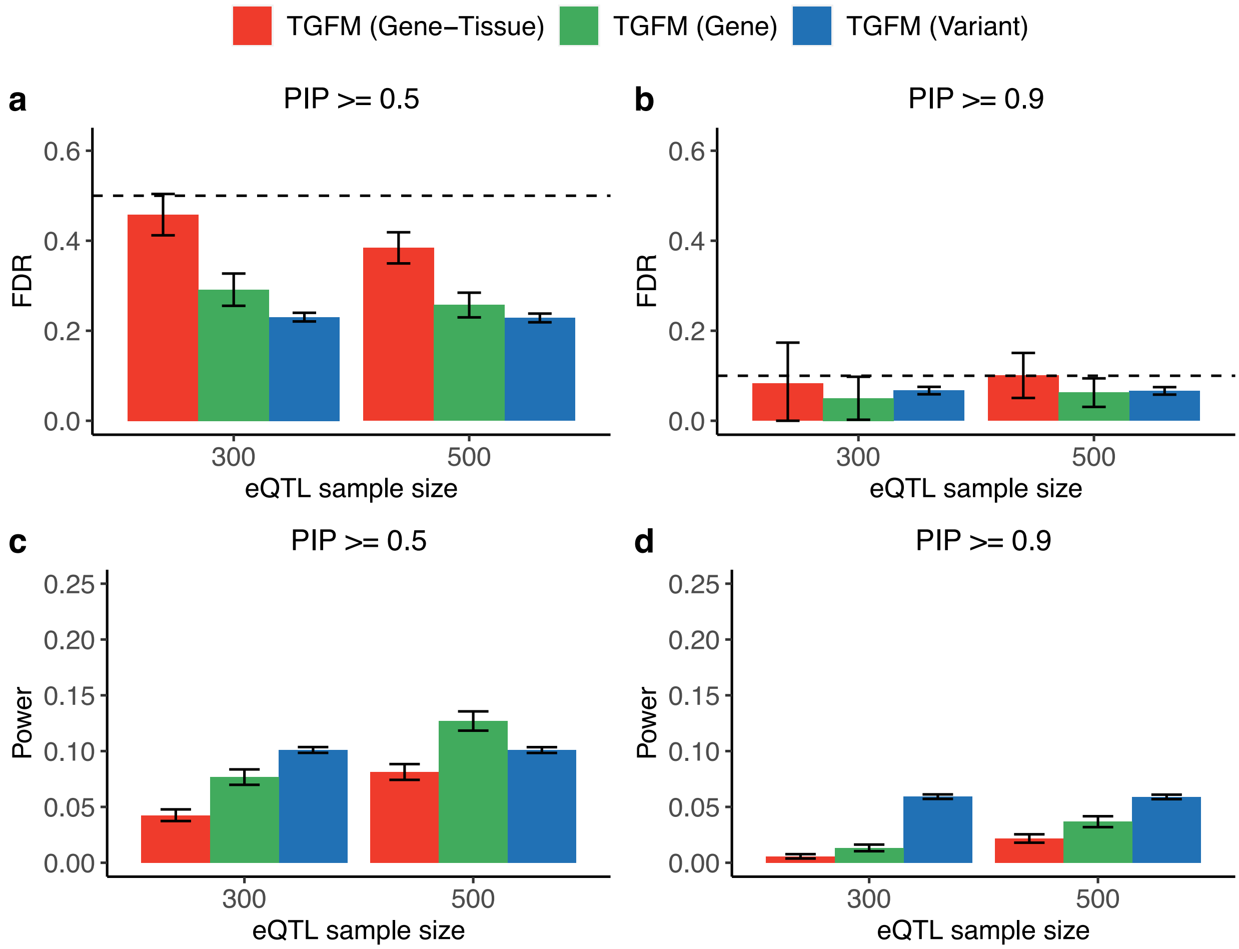


**Supplementary Figure 8: Calibration and power of fine-mapping in simulation setting where the number of causal eQTLs are shared across genes.** In this simulation setting, causal eQTLs are shared across nearby genes (and shared across tissues, as in our primary simulations). Specifically, in each simulation we randomly selected 250 pairs of genes with overlapping cis-windows (of 1,976 genes total) to have 2 shared causal eQTLs, with effect size correlation set to 0.74 across all gene-tissue pairs corresponding to the 2 genes. Two genes had overlapping cis-windows if the cis window overlap was greater than 25000 base pairs and there exist at least 10 snps in the overlapping window. Here, we report **(a,b)** Average fine-mapping FDR across 100 simulations using TGFM for different classes of genetic elements (see legend) across *eQTL* sample sizes (x-axis) at PIP=0.5 **(a)** and PIP=0.9 **(b)**. Dashed horizontal line denotes 1 – PIP threshold (see main text). **(c,d)** Average fine-mapping power across 100 simulations using TGFM for different classes of genetic elements (see legend) across *eQTL* sample sizes (x-axis) at PIP=0.5 **(c)** and PIP=0.9 **(d)**. Error bars denote 95% confidence intervals. This supplementary figure is similar to Figure 2, except it only shows simulation results at eQTL sample size of 300 and 500, *and* it based on data from an alternative simulation setting described at the beginning of the caption.

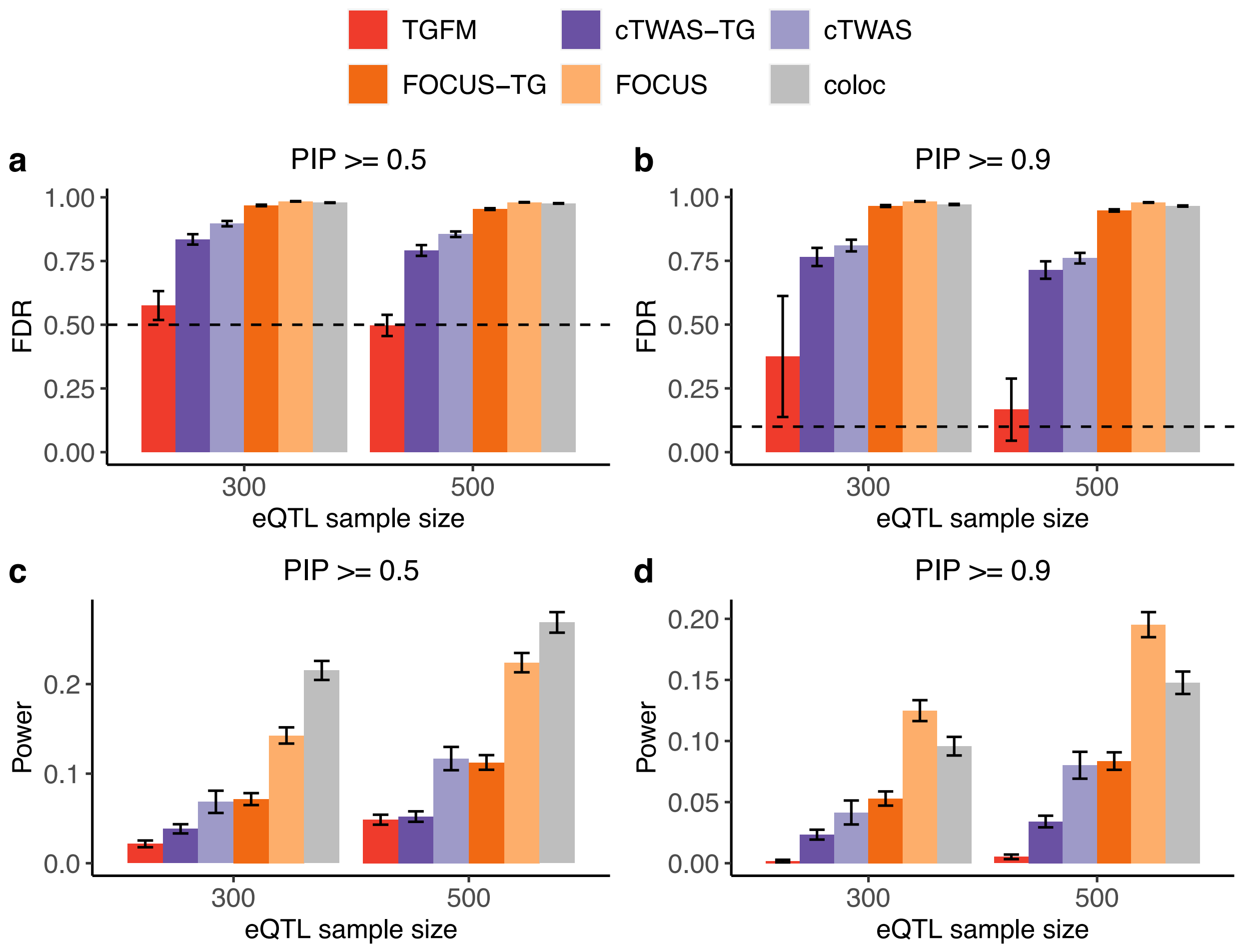


**Supplementary Figure 9: Calibration and power of fine-mapping in simulation setting where disease-causal genes are under selection.** In this simulation setting, eQTLs for disease-causal genes (actually gene-tissue pairs) had lower eQTL effect sizes, thus explaining lower cis-heritability. Specifically, the expected per-SNP heritability for a causal cis-eQTL of a causal gene-tissue pair was set to 0.01 (vs 0.015 for a causal cis-eQTL of a non-causal gene-tissue pair). If variant was a causal eQTL for more than 1 gene-tissue pair and at least 1 of the gene-tissue pairs was causal, then the eQTL had a per-SNP heritability of 0.01 for all linked gene-tissue pairs; this choice was made because selection acts on the variant, not the variant-gene-tissue pair. Here, we report **(a,b)** Average gene-tissue fine-mapping FDR across 100 simulations using various fine-mapping (see legend) across *eQTL* sample sizes (x-axis) at PIP=0.5 **(a)** and PIP=0.9 **(b)**. Dashed horizontal line denotes 1 – PIP threshold (see main text). **(c,d)** Average gene-tissue fine-mapping power across 100 simulations using various fine-mapping methods (see legend) across *eQTL* sample sizes (x-axis) at PIP=0.5 **(c)** and PIP=0.9 **(d)**. Error bars denote 95% confidence intervals. This supplementary figure is similar to Figure 1, except it only shows simulation results at eQTL sample size of 300 and 500, *and* it based on data from an alternative simulation setting described at the beginning of the caption.

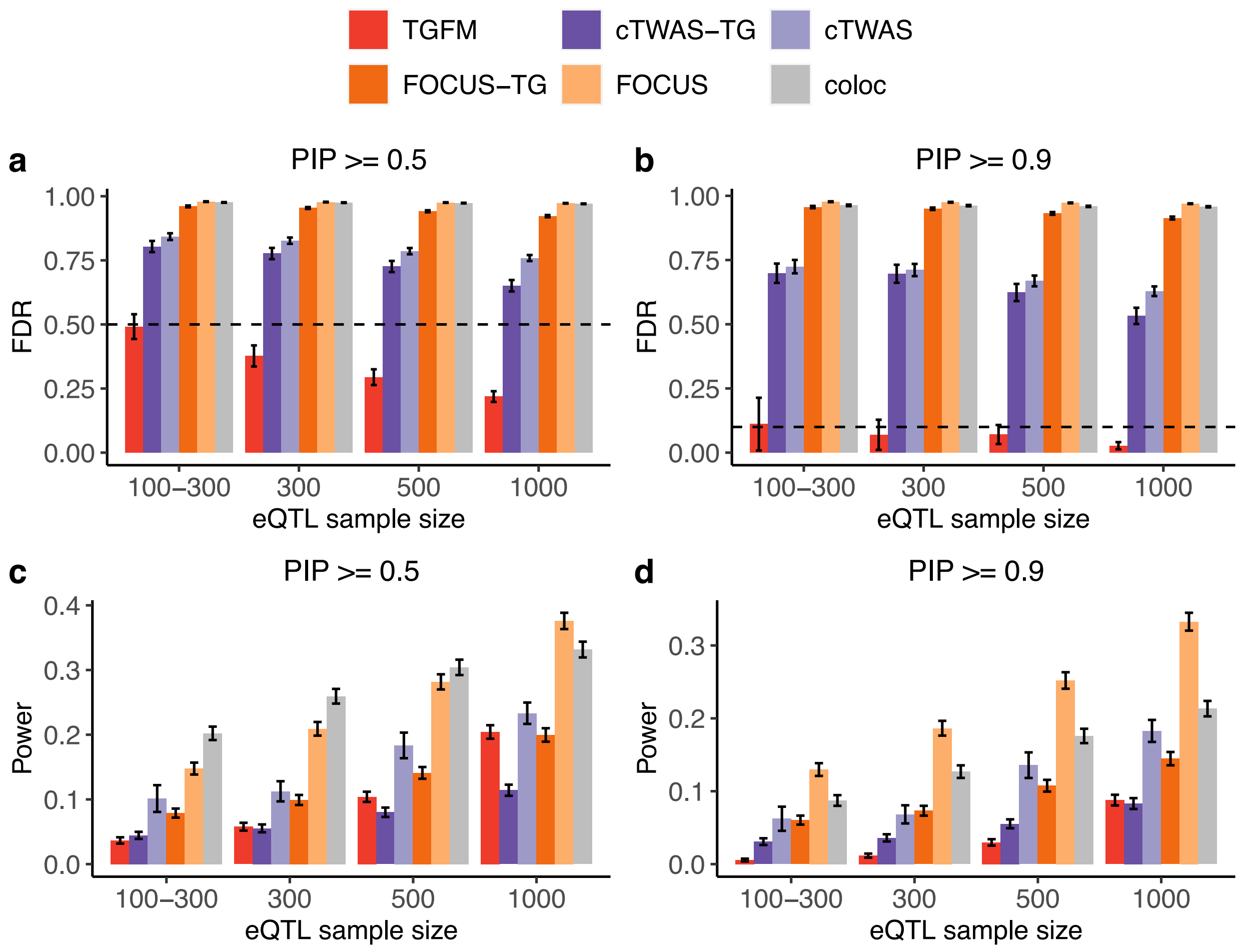


**Supplementary Figure 10: Calibration and power of tissue-gene fine-mapping methods in simulations with one causal tissue.** This supplementary figure is similar to Figure 1, except it is performed in a simulation setting with a single causal tissue per simulated trait (instead of two causal tissues per-trait in the primary simulations). **(a,b)** Average tissue-gene fine-mapping FDR across 100 simulations for various fine-mapping methods (see legend) across eQTL sample sizes (x-axis) at PIP=0.5 **(a)** and PIP=0.9 **(b)**. Single thick, dashed horizontal line denotes 1 – PIP threshold (see main text). **(c,d)** Average tissue-gene fine-mapping power across 100 simulations for various fine-mapping methods (see legend) across eQTL sample sizes (x-axis) at PIP=0.5 **(c)** and PIP=0.9 **(d)**. Error bars denote 95% confidence intervals.

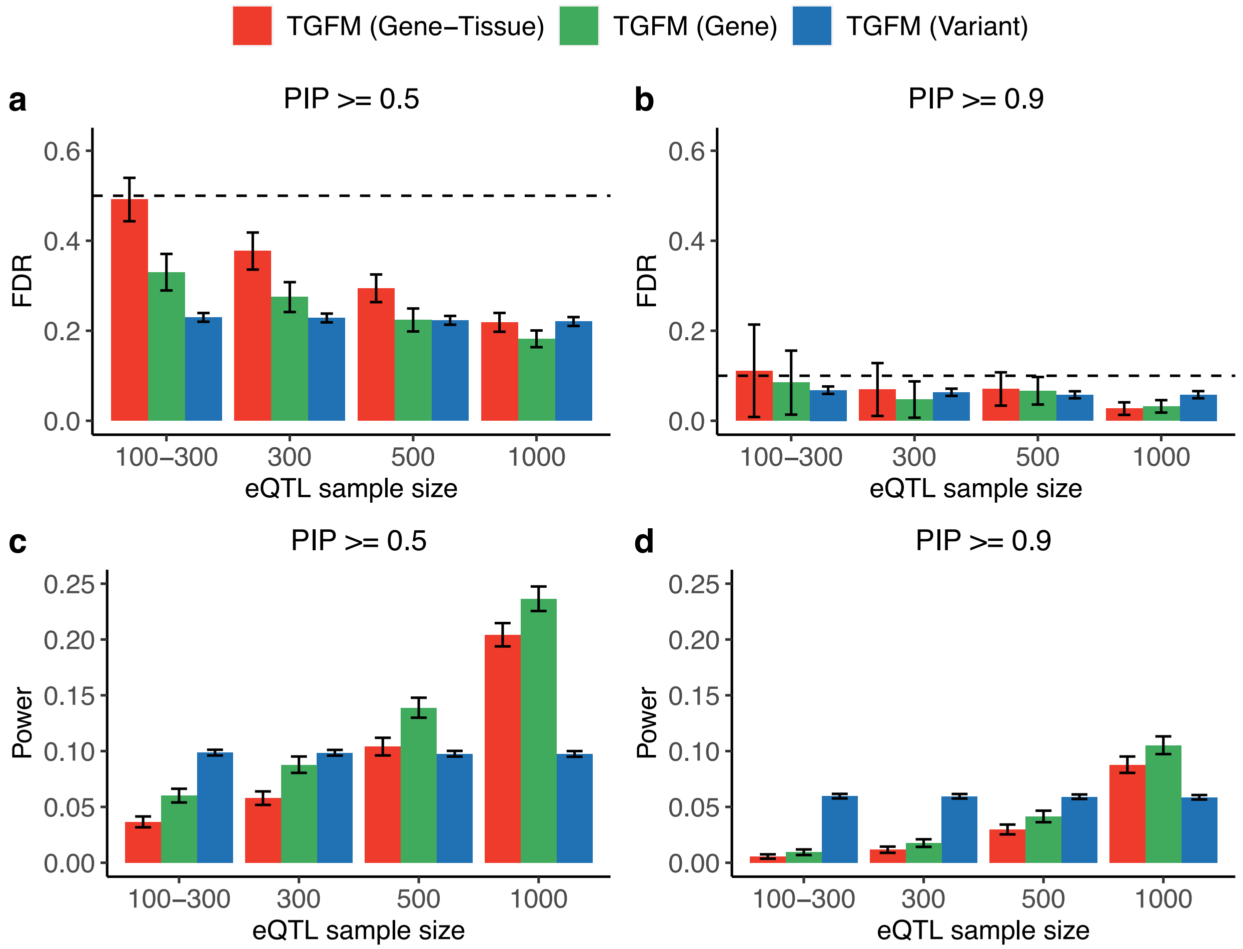


**Supplementary Figure 11: Calibration and power of fine-mapping different classes of genetic elements with TGFM in simulations with one causal tissue.** This supplementary figure is similar to Figure 2, except it is performed in a simulation setting with a single causal tissue per simulated trait (instead of two causal tissues per-trait in the primary simulations). **(a,b)** Average fine-mapping FDR across 100 simulations using TGFM for different classes of genetic elements (see legend) across *eQTL* sample sizes (x-axis) at PIP=0.5 **(a)** and PIP=0.9 **(b)**. Dashed horizontal line denotes 1 – PIP threshold (see main text). **(c,d)** Average fine-mapping power across 100 simulations using TGFM for different classes of genetic elements (see legend) across *eQTL* sample sizes (x-axis) at PIP=0.5 **(c)** and PIP=0.9 **(d)**. Error bars denote 95% confidence intervals.

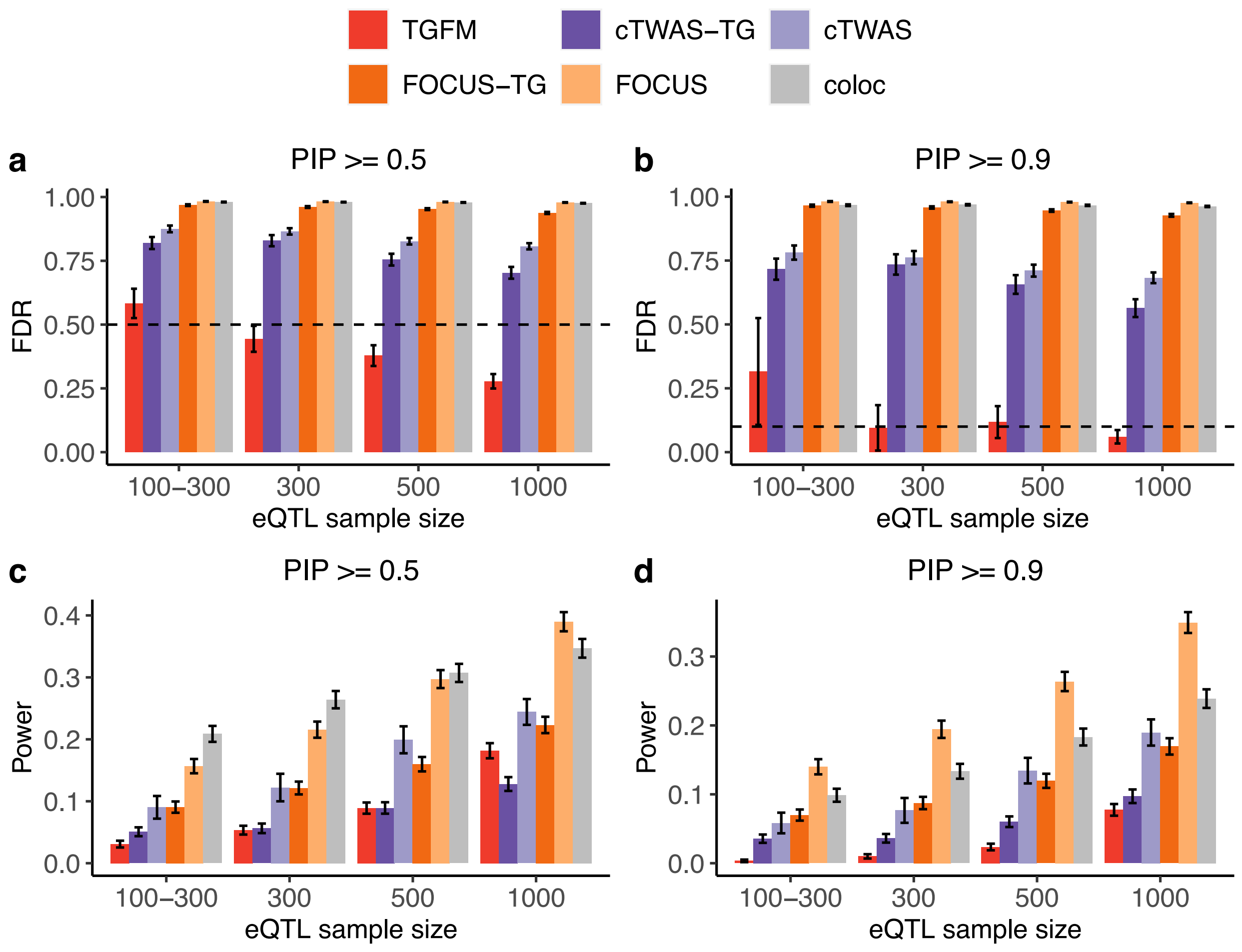


**Supplementary Figure 12: Calibration and power of tissue-gene fine-mapping methods when restricting to loci containing at most 1 causal gene-tissue pair in simulations.** This supplementary figure is similar to Figure 1, except it is restricted to loci containing at most 1 causal gene-tissue pair. Specifically, gene-tissue pairs were only evaluated in this analysis if there exists at most 1 causal gene-tissue pair within 500kb of the TSS of the gene-tissue pair. **(a,b)** Average tissue-gene fine-mapping FDR across 100 simulations for various fine-mapping methods (see legend) across eQTL sample sizes (x-axis) at PIP=0.5 **(a)** and PIP=0.9 **(b)**. Single thick, dashed horizontal line denotes 1 – PIP threshold (see main text). **(c,d)** Average tissue-gene fine-mapping power across 100 simulations for various fine-mapping methods (see legend) across eQTL sample sizes (x-axis) at PIP=0.5 **(c)** and PIP=0.9 **(d)**. Error bars denote 95% confidence intervals.

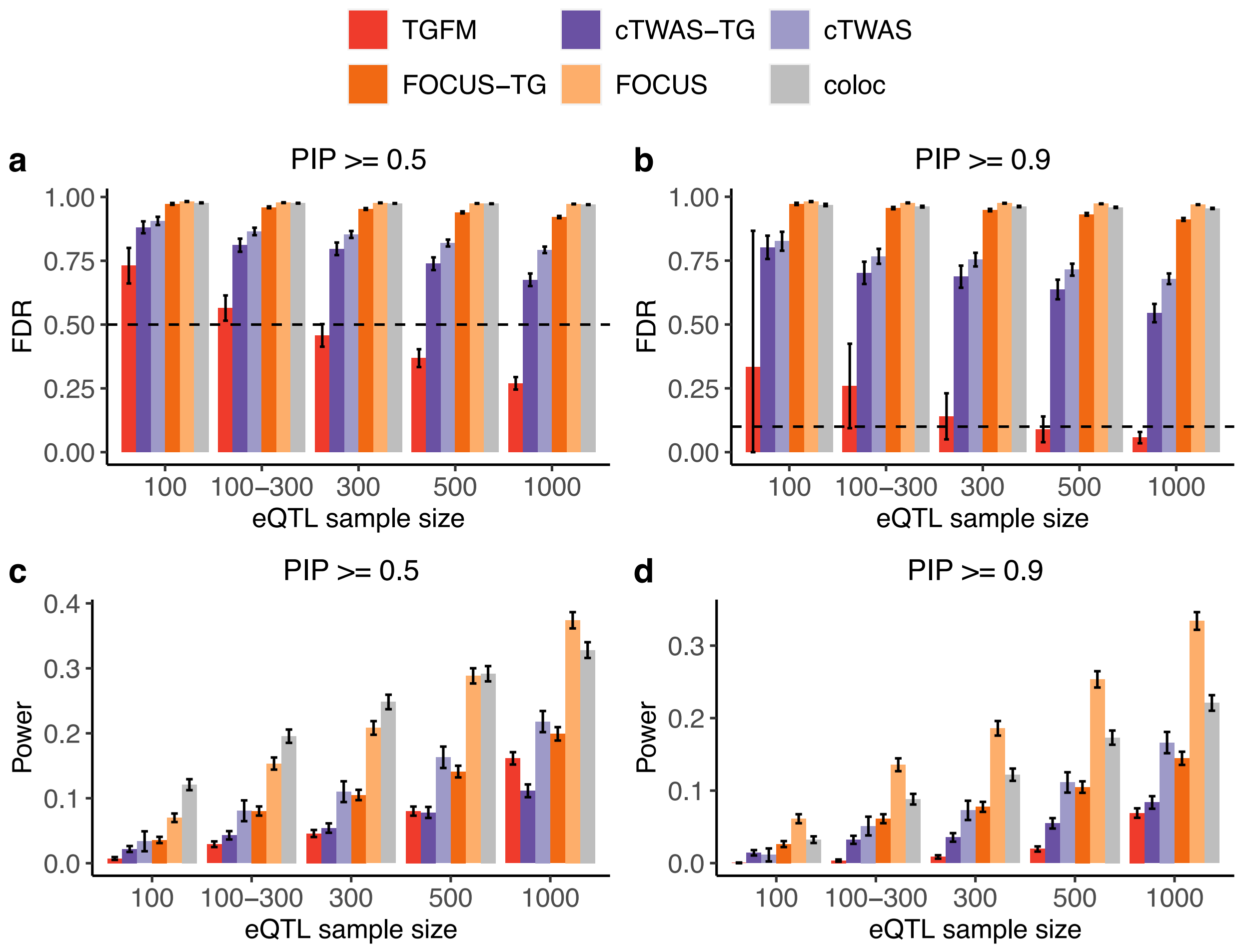


**Supplementary Figure 13: Calibration and power of tissue-gene fine-mapping methods at various PIP thresholds and eQTL sample sizes in simulations.** This supplementary figure is similar to Figure 1, except it also shows simulations results at eQTL sample of 100. **(a,b)** Average tissue-gene fine-mapping FDR across 100 simulations for various fine-mapping methods (see legend) across eQTL sample sizes (x-axis) at PIP=0.5 **(a)** and PIP=0.9 **(b)**. Single thick, dashed horizontal line denotes 1 – PIP threshold (see main text). **(c,d)** Average tissue-gene fine-mapping power across 100 simulations for various fine-mapping methods (see legend) across eQTL sample sizes (x-axis) at PIP=0.5 **(c)** and PIP=0.9 **(d)**. Error bars denote 95% confidence intervals.

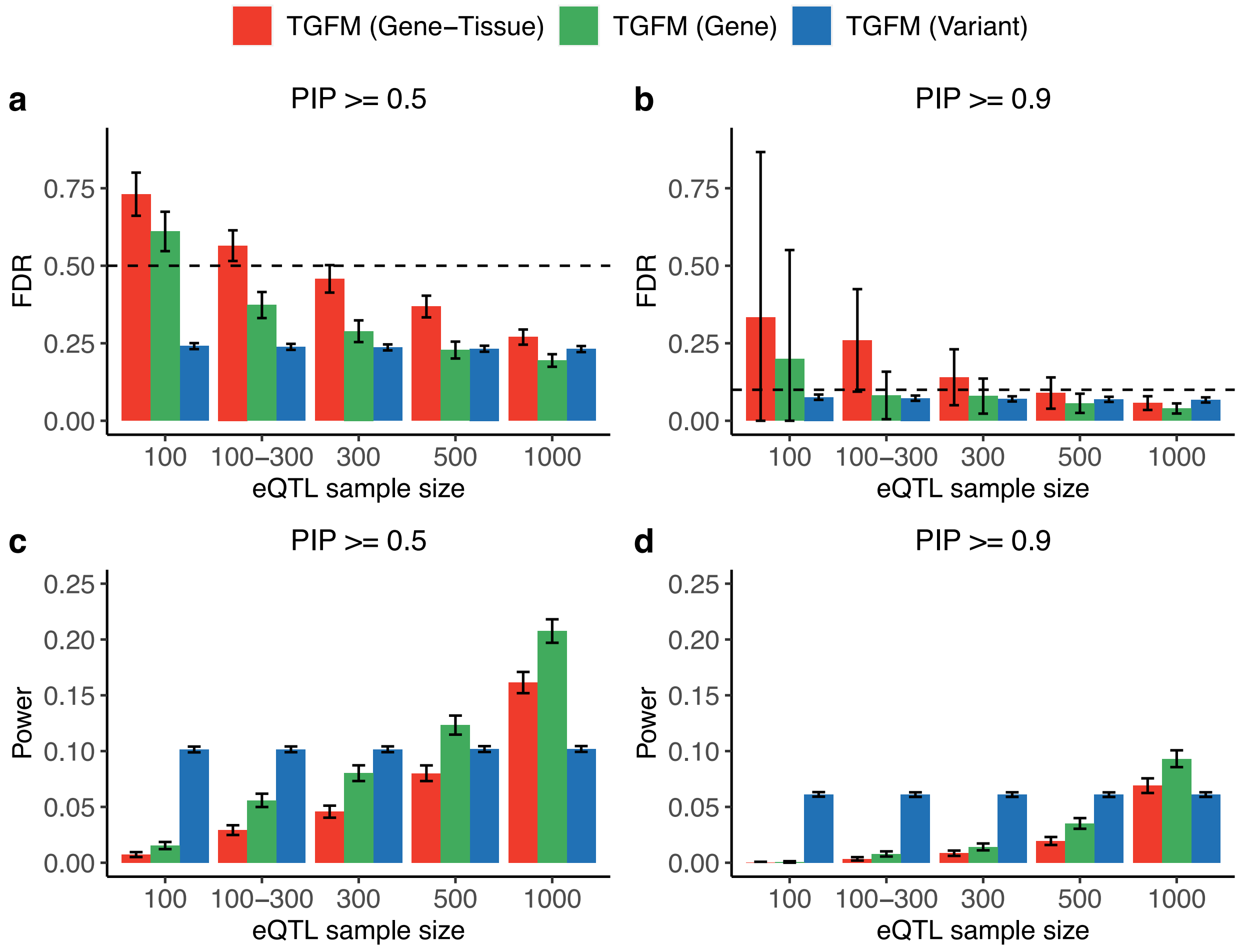


**Supplementary 14: Calibration and power of fine-mapping different classes of genetic elements with TGFM in simulations for various simulated eQTL sample sizes.** This supplementary figure is similar to Figure 2, except it also shows simulations results at eQTL sample of 100. **(a,b)** Average fine-mapping FDR across *100* simulations using TGFM for different classes of genetic elements (see legend) across *eQTL* sample sizes (x-axis) at PIP=0.5 **(a)** and PIP=0.9 **(b)**. Dashed horizontal line denotes 1 – PIP threshold (see main text). **(c,d)** Average fine-mapping power across *100* simulations using TGFM for different classes of genetic elements (see legend) across *eQTL* sample sizes (x-axis) at PIP=0.5 **(c)** and PIP=0.9 **(d)**. Error bars denote 95% confidence intervals.

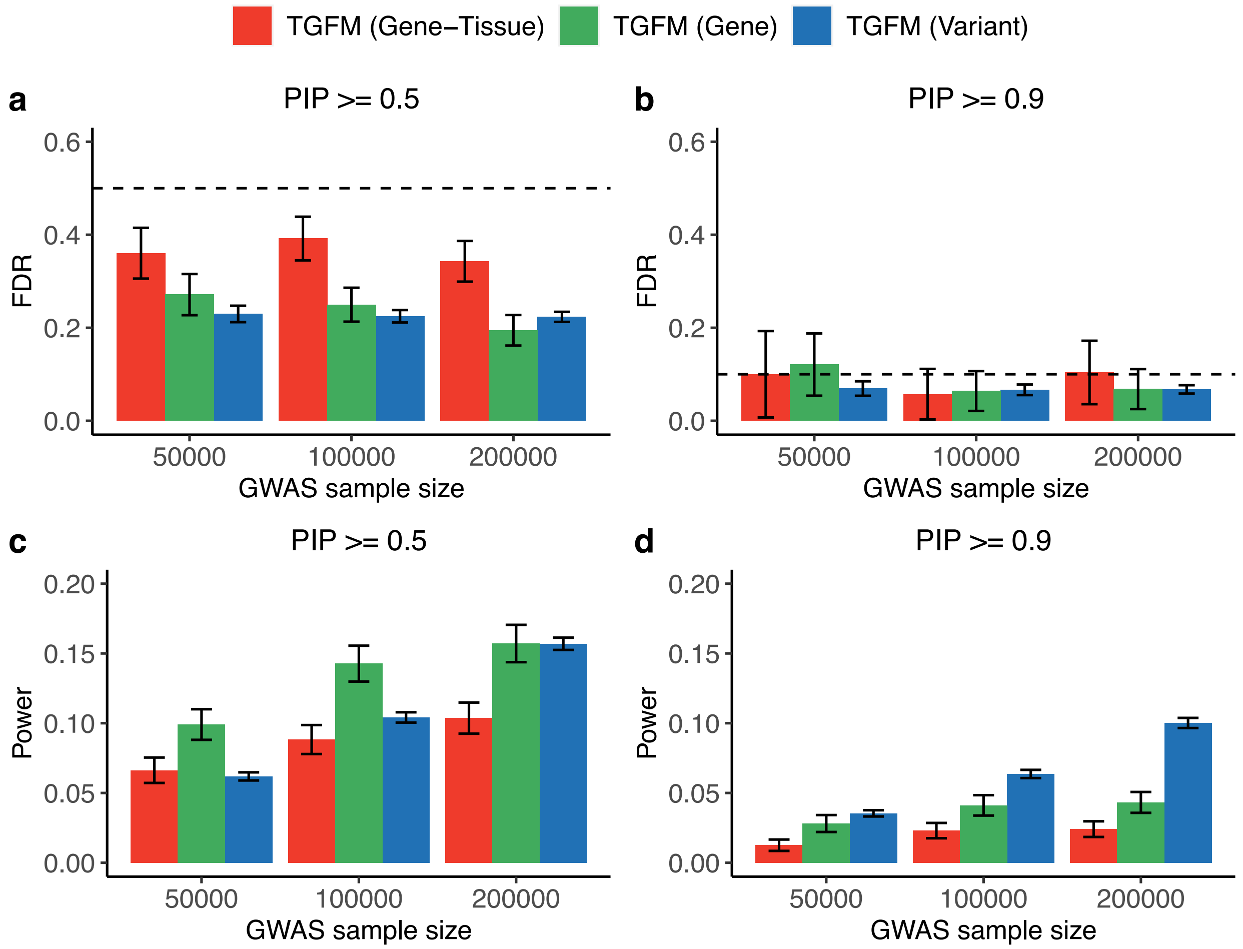


**Supplementary Figure 15: Calibration and power of fine-mapping different classes of genetic elements with TGFM in simulations for various simulated GWAS sample sizes. (a,b)** Average fine-mapping FDR across 50 simulations using TGFM for different classes of genetic elements (see legend) across GWAS sample sizes (x-axis) at PIP=0.5 **(a)** and PIP=0.9 **(b)** at eQTL sample size of 500. Dashed horizontal line denotes 1 – PIP threshold (see main text). **(c,d)** Average fine-mapping power across 50 simulations using TGFM for different classes of genetic elements (see legend) across GWAS sample sizes (x-axis) at PIP=0.5 **(c)** and PIP=0.9 **(d)** at eQTL sample size of 500. Error bars denote 95% confidence intervals. This supplementary figure is similar to Figure 2, except it shows results at a fixed eQTL sample of 500, and varies the GWAS sample size.

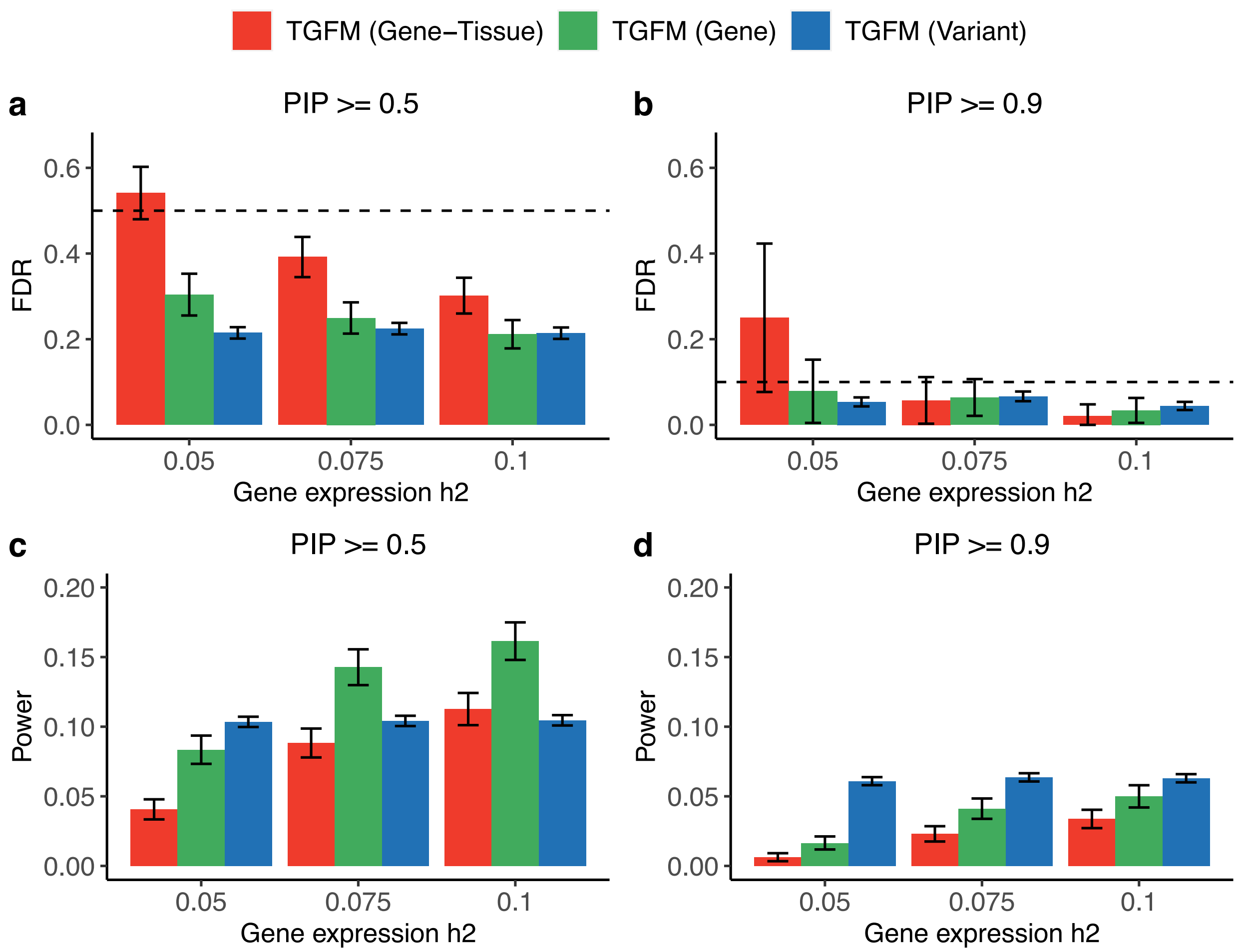


**Supplementary Figure 16: Calibration and power of fine-mapping different classes of genetic elements with TGFM in simulations for various simulated gene expression heritabilities. (a,b)** Average fine-mapping FDR across 50 simulations using TGFM for different classes of genetic elements (see legend) across simulated gene expression heritabilities (x-axis) at PIP=0.5 **(a)** and PIP=0.9 **(b)** at eQTL sample size of 500. Dashed horizontal line denotes 1 – PIP threshold (see main text). **(c,d)** Average fine-mapping power across 50 simulations using TGFM for different classes of genetic elements (see legend) across simulated gene expression heritabilities (x-axis) at PIP=0.5 **(c)** and PIP=0.9 **(d)** at eQTL sample size of 500. Error bars denote 95% confidence intervals. This supplementary figure is similar to Figure 2, except it shows results at a fixed eQTL sample of 500, and varies the gene expression heritability.

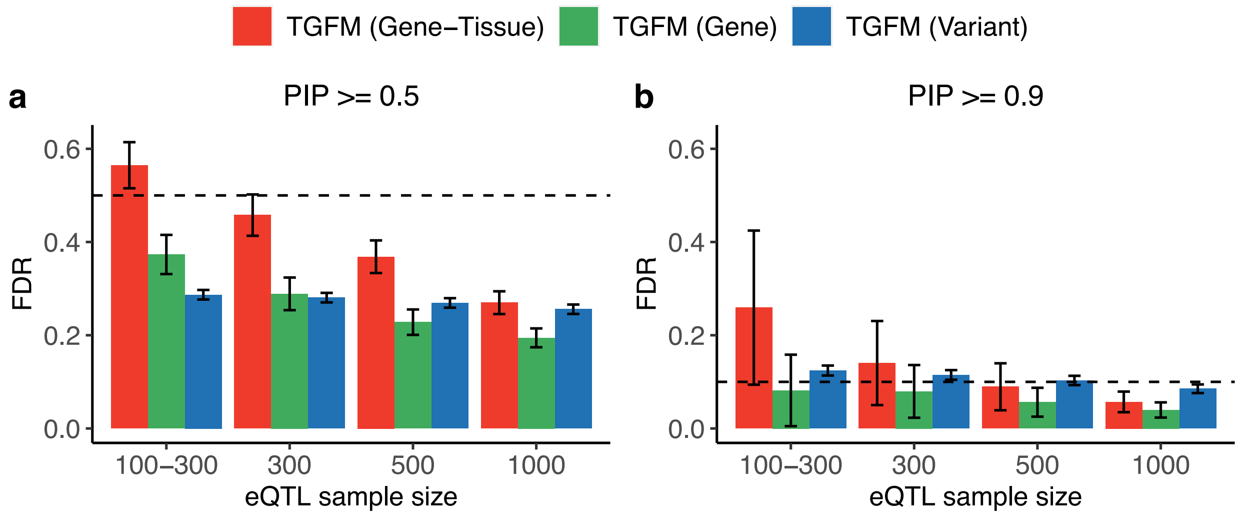


**Supplementary Figure 17: Calibration of fine-mapping genetic elements with TGFM at various PIP thresholds in simulations using alternative definition of false-positive variants.** Average fine-mapping FDR across 100 simulations using TGFM for different classes of genetic elements (see legend) across eQTL sample sizes (x-axis) at PIP=0.5 **(a)** and PIP=0.9. Dashed horizontal line denotes 1 – PIP threshold (see main text). Error bars denote 95% confidence intervals. Unlike Figure 2a-b, causal eQTL variants for causal gene-tissue pairs were considered false positives for variant-level calibration. TGFM (Variant) is mis-calibrated at eQTL sample size=300 and PIP >= 0.9. The calibration worsened at small eQTL sample sizes, likely due to decreased power to detect causal gene-tissue pairs at small eQTL sample sizes, forcing the unmodeled gene-tissue pair effects to be captured by non-mediated variants.

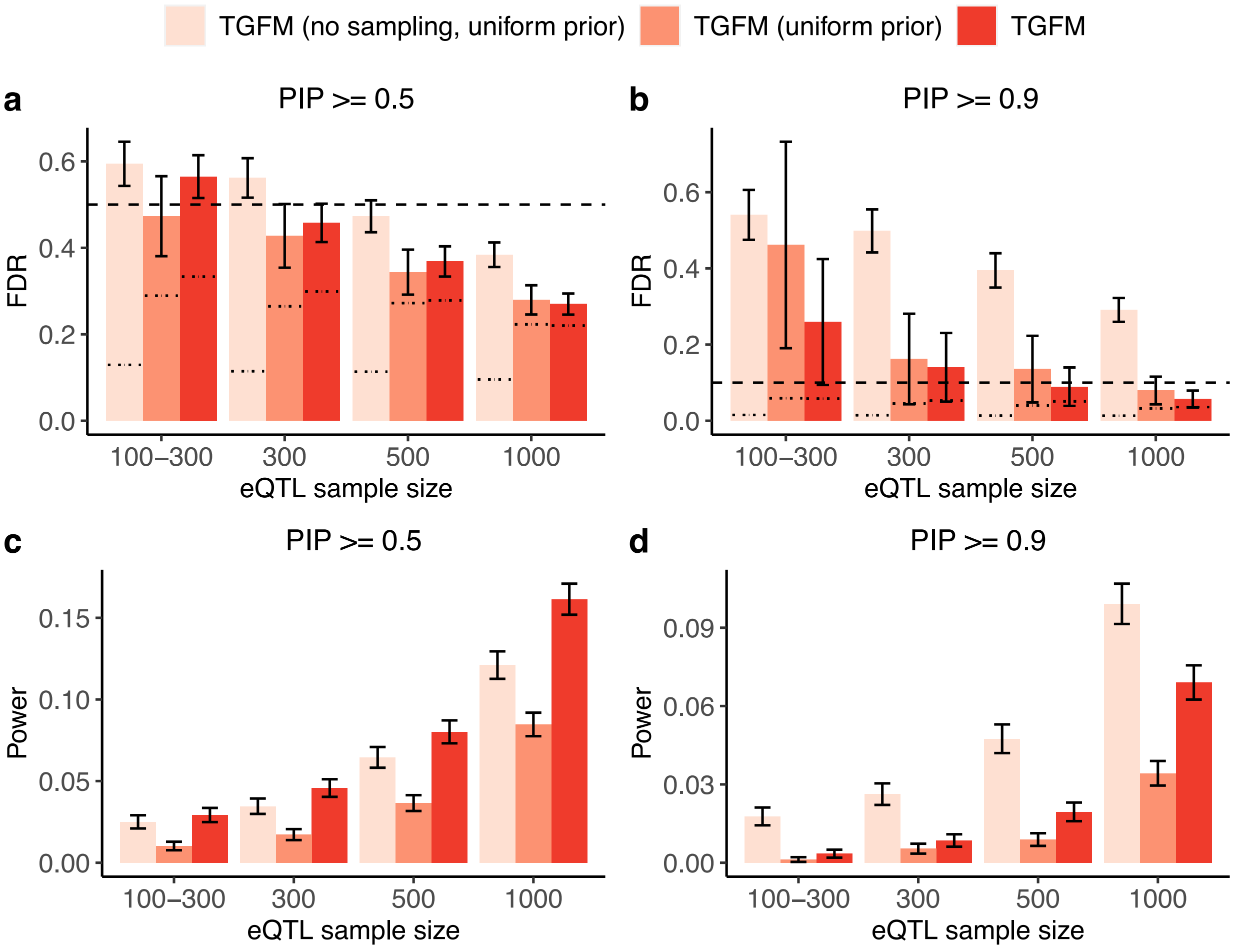


**Supplementary Figure 18: Calibration and power of tissue-gene fine-mapping for various versions of TGFM in simulations. (a,b)** Average tissue-gene fine-mapping FDR across 100 simulations for various fine-mapping methods (see legend) across eQTL sample sizes (x-axis) at PIP=0.5 **(a)** and PIP=0.9 **(b)**. Single thick, dashed horizontal line denotes 1 – PIP threshold (see main text). The thin dashed horizontal lines specific to each bar denotes (1 – average PIP) (where average is taken across all genetic elements belonging to that bar; see main text). **(c,d)** Average tissue-gene fine-mapping power across 100 simulations for various fine-mapping methods (see legend) across eQTL sample sizes (x-axis) at PIP=0.5 **(c)** and PIP=0.9 **(d)**. Error bars denote 95% confidence intervals. This supplementary figure is similar to Figure 1, except it shows calibration and power of additional tissue-gene fine-mapping methods. "TGFM (no sampling, uniform prior)" corresponds to TGFM (Gene-Tissue) with a uniform prior and a single cis-predicted expression model (based on posterior mean causal cis-eQTL effect sizes) instead of averaging results across 100 sampled cis-predicted expression models. "TGFM (uniform prior)" corresponds to TGFM (Gene-Tissue) with a uniform prior. "TGFM" corresponds to the default version of TGFM (Gene-Tissue) (shown in Figure 1).

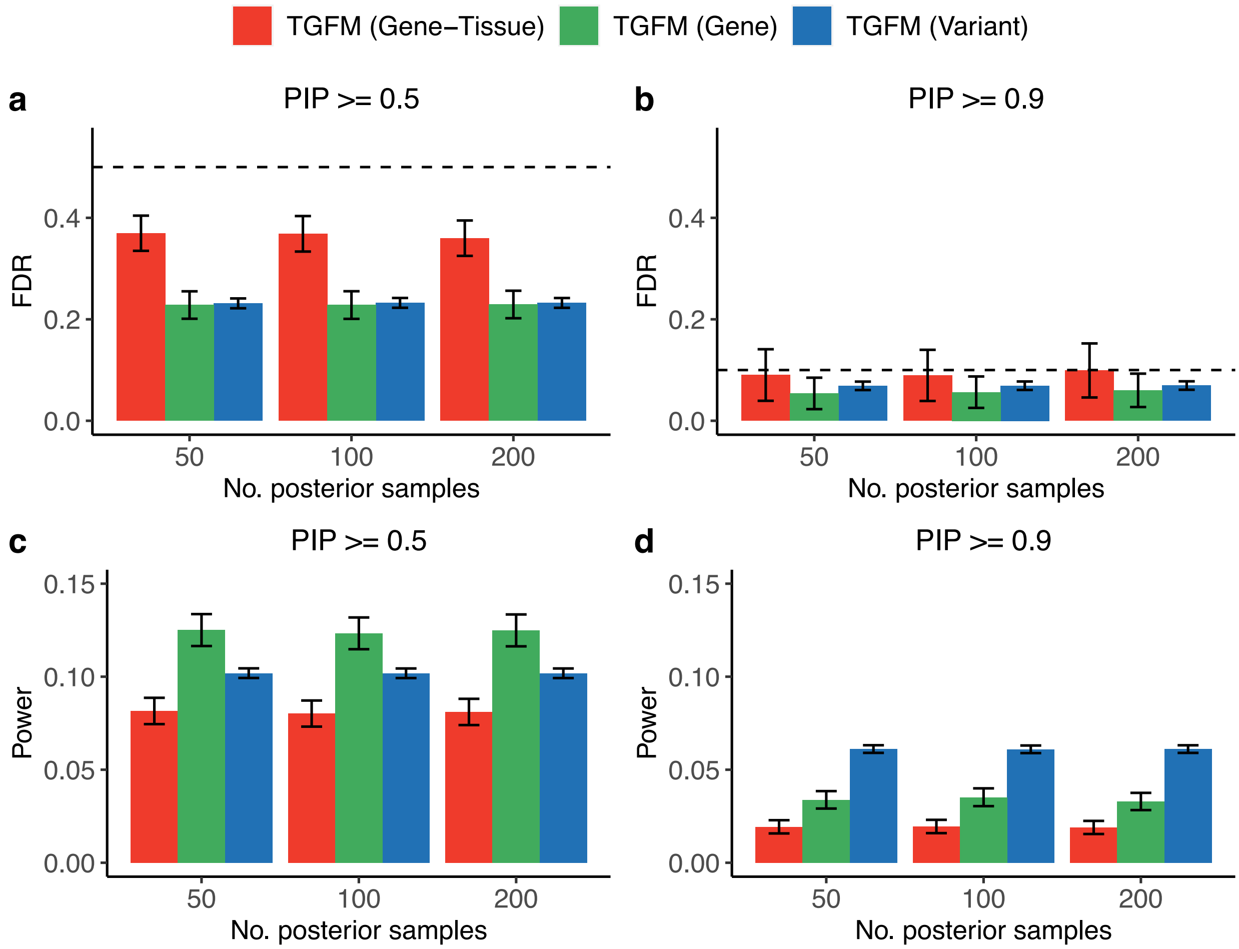


**Supplementary Figure 19*:* Calibration and power of fine-mapping different classes of genetic elements with TGFM using a variable number of posterior samples in simulations. (a,b)** Average fine-mapping FDR across 100 simulations using TGFM for different classes of genetic elements (see legend) as a function of the number of posterior samples used by TGFM (x-axis) at PIP=0.5 **(a)** and PIP=0.9 **(b)** at an eQTL sample size of 500. Dashed horizontal line denotes 1 – PIP threshold (see main text). **(c,d)** Average fine-mapping power across *100* simulations using TGFM for different classes of genetic elements (see legend) as a function of the number of posterior samples used by TGFM (x-axis) at PIP=0.5 **(c)** and PIP=0.9 **(d)** at an eQTL sample size of 500. Error bars denote 95% confidence intervals. This supplementary figure is similar to Figure 2, except it shows results at a fixed eQTL sample of 500, and varies the number of posterior samples used by TGFM.

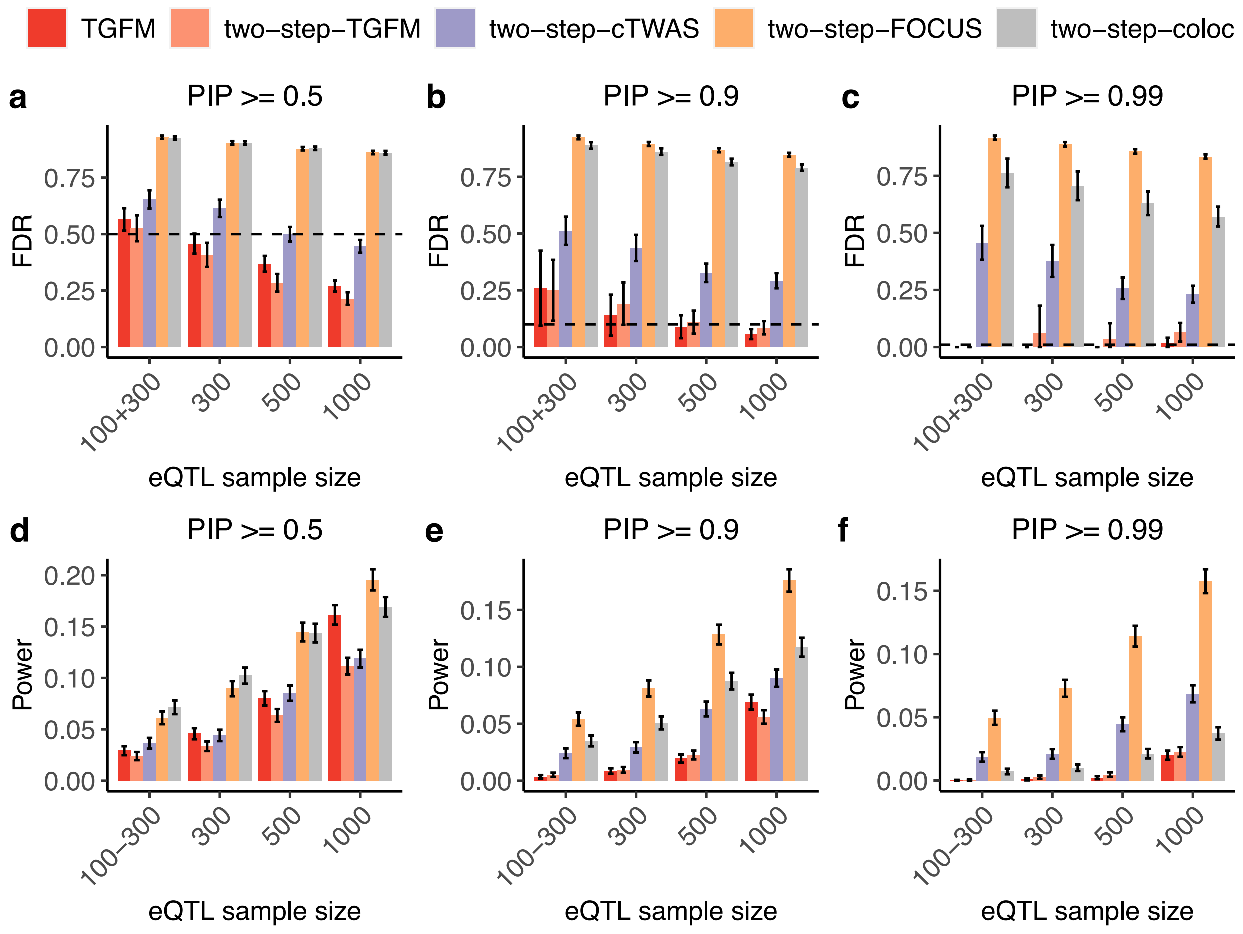


**Supplementary Figure 20: Calibration and power of gene-tissue fine-mapping using two-step fine-mapping methods in simulations with 2 causal tissues. (a-c)** Average gene-tissue fine-mapping FDR across 100 simulations using TGFM and various two-step fine-mapping (see legend) across *eQTL* sample sizes (x-axis) at PIP=0.5 **(a)**, PIP=0.9 **(b)**, and PIP=0.99. Dashed horizontal line denotes 1 – PIP threshold (see main text). **(d-f)** Average gene-tissue fine-mapping power across 100 simulations using TGFM and various two-step fine-mapping methods (see legend) across *eQTL* sample sizes (x-axis) at PIP=0.5 **(d)**, PIP=0.9 **(e)**, and PIP=0.99 **(f)**. Error bars denote 95% confidence intervals. Analysis was performed using simulated data from the primary simulations. The two-step fine-mapping approach first infers the causal tissue and then performs traditional gene-level fine-mapping in the inferred causal tissue. We inferred the causal tissue using the TGFM tissue-specific prior by taking the tissue with the smallest p-value, and performed gene-level fine-mapping in the inferred causal tissue using either TGFM (applied to consider only a single tissue), coloc, FOCUS, or cTWAS; we refer to these two-step fine-mapping methods as two-step-TGFM, two-step-coloc, two-step-FOCUS, and two-step-cTWAS, respectively.

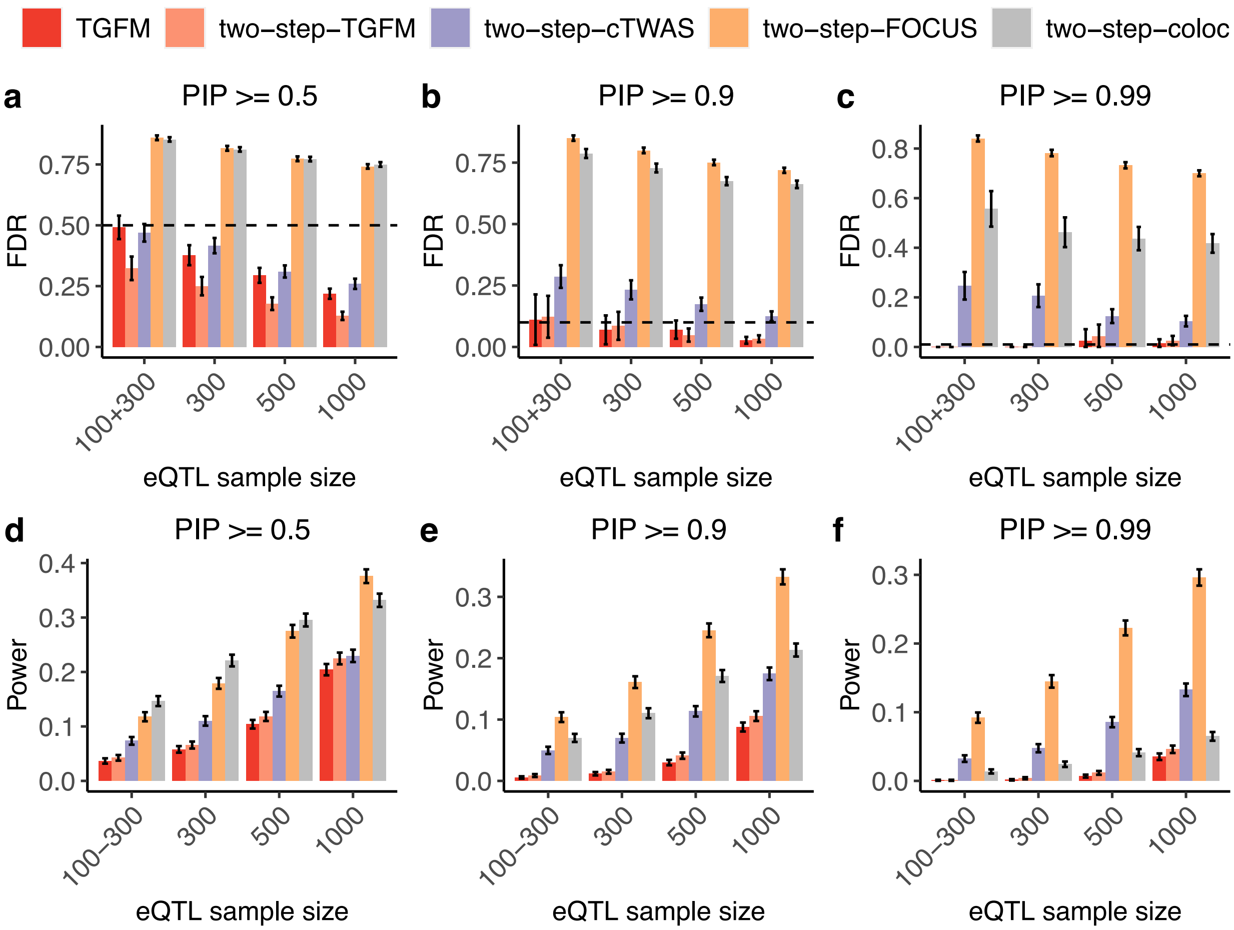


**Supplementary Figure 21: Calibration and power of gene-tissue fine-mapping using two-step fine-mapping methods in simulations with 1 causal tissue. (a-c)** Average gene-tissue fine-mapping FDR across 100 simulations using TGFM and various two-step fine-mapping (see legend) across *eQTL* sample sizes (x-axis) at PIP=0.5 **(a)**, PIP=0.9 **(b)**, and PIP=0.99. Dashed horizontal line denotes 1 – PIP threshold (see main text). **(d-f)** Average gene-tissue fine-mapping power across 100 simulations using TGFM and various two-step fine-mapping methods (see legend) across *eQTL* sample sizes (x-axis) at PIP=0.5 **(d)**, PIP=0.9 **(e)**, and PIP=0.99 **(f)**. Error bars denote 95% confidence intervals. Analysis was performed using simulated data from a simulation setting with one causal tissue (instead of 2 causal tissues in primary simulation). The two-step fine-mapping approach first infers the causal tissue and then performs traditional gene-level fine-mapping in the inferred causal tissue. We inferred the causal tissue using the TGFM tissue-specific prior by taking the tissue with the smallest p-value, and performed gene-level fine-mapping in the inferred causal tissue using either TGFM (applied to consider only a single tissue), coloc, FOCUS, or cTWAS; we refer to these two-step fine-mapping methods as two-step-TGFM, two-step-coloc, two-step-FOCUS, and two-step-cTWAS, respectively.

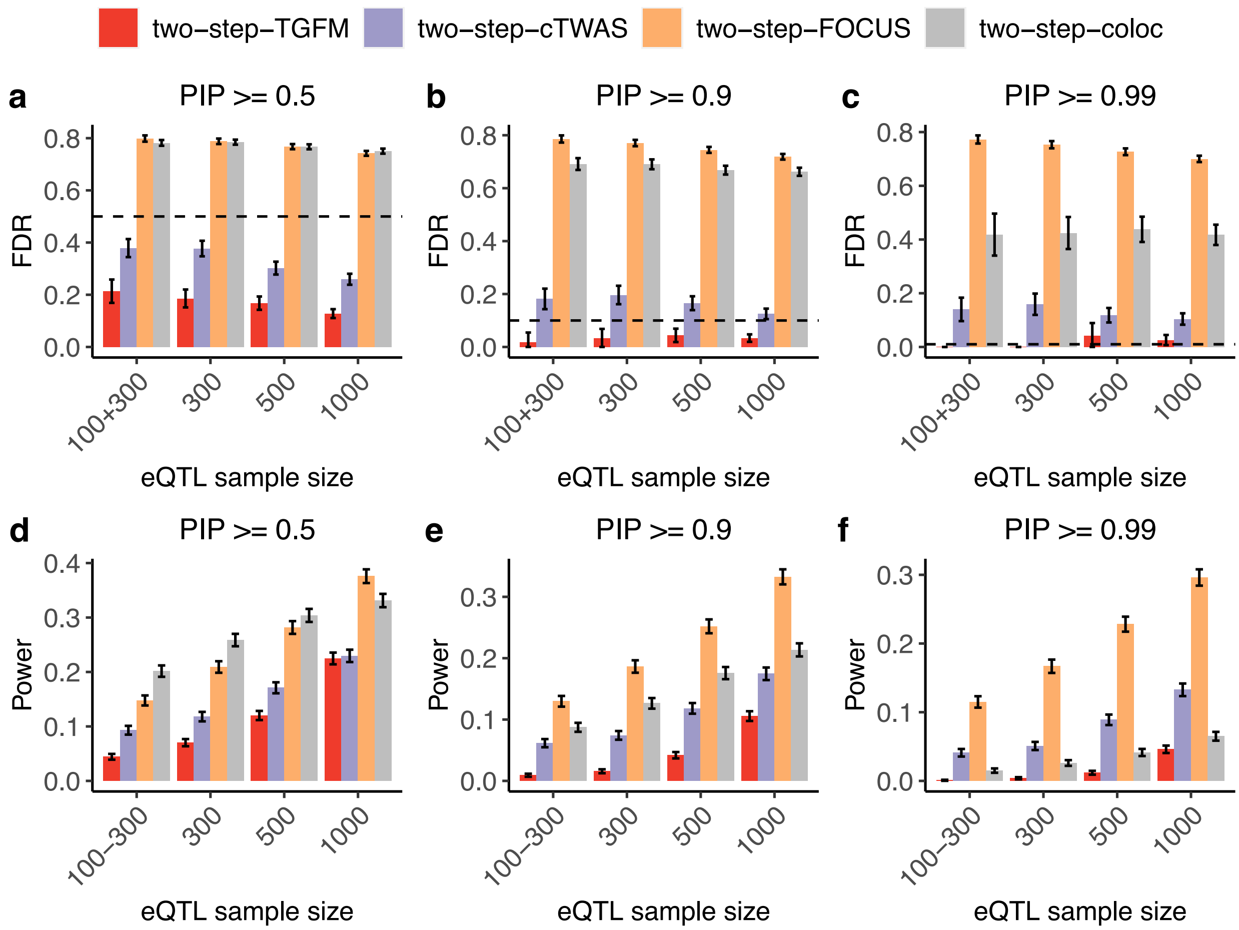


**Supplementary Figure 22*:* Calibration and power of gene fine-mapping in a simulation setting where single causal tissue is known.** Here, we consider a simulation setting in which the single causal tissue is known, and we compared TGFM (applied to consider only a single tissue) to coloc, FOCUS, and cTWAS. **(a,b)** Average gene fine-mapping FDR across 100 simulations using various fine-mapping (see legend) across *eQTL* sample sizes (x-axis) at PIP=0.5 **(a)** and PIP=0.9 **(b)**. Dashed horizontal line denotes 1 – PIP threshold (see main text). **(c,d)** Average gene fine-mapping power across 100 simulations using various fine-mapping methods (see legend) across *eQTL* sample sizes (x-axis) at PIP=0.5 **(c)** and PIP=0.9 **(d)**. Error bars denote 95% confidence intervals. TGFM remains well-calibrated, whereas coloc, FOCUS, and cTWAS were strongly mis-calibrated; this implies that TGFM is an advance over previous methods even within the scope of traditional gene-level fine-mapping.

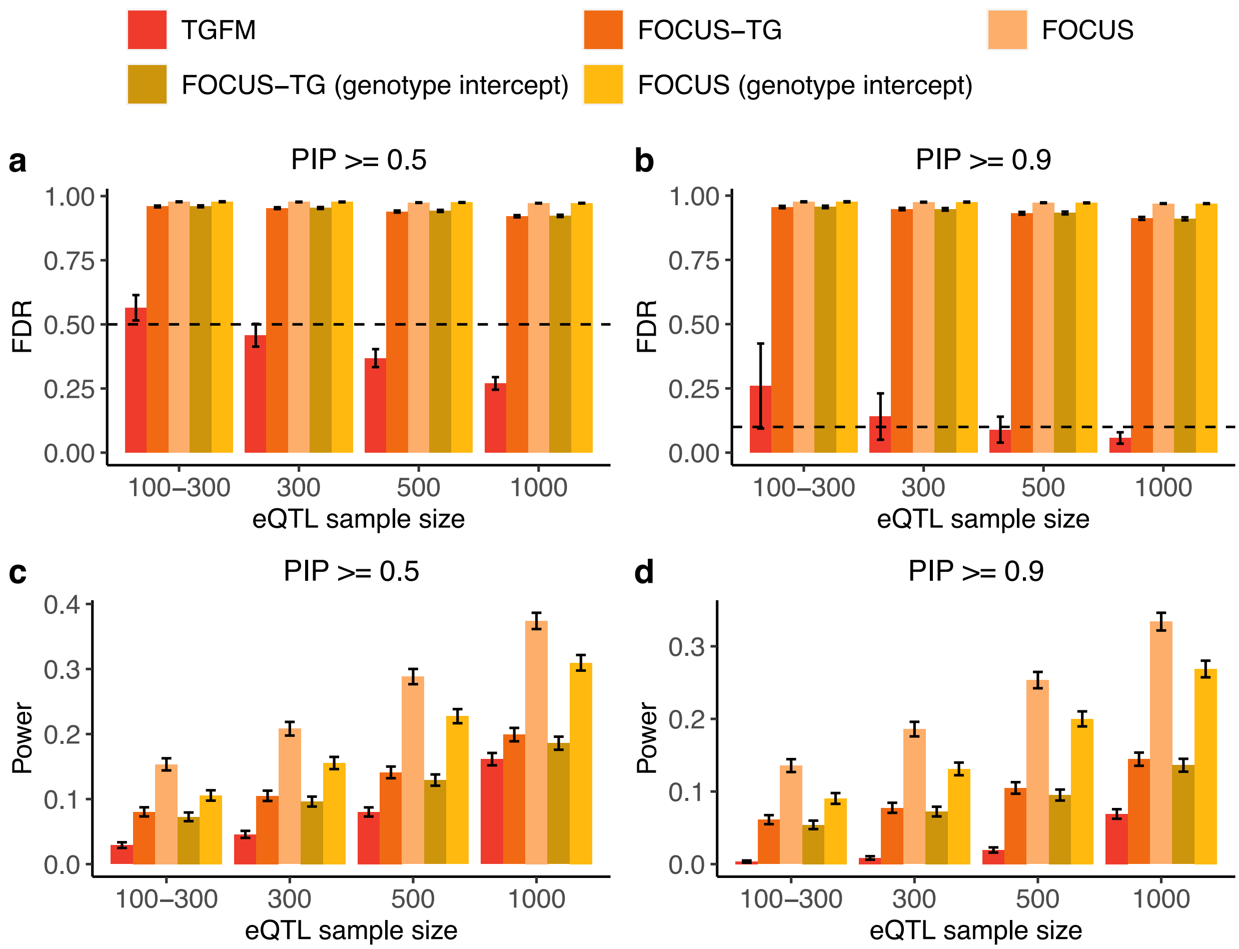


**Supplementary Figure 23: Calibration and power of gene-tissue fine-mapping using FOCUS with and without the genotype intercept. (a,b)** Average gene-tissue fine-mapping FDR across 100 simulations using various fine-mapping methods (see legend) across *eQTL* sample sizes (x-axis) at PIP=0.5 **(a)** and PIP=0.9 **(b)**. Dashed horizontal line denotes 1 – PIP threshold (see main text). **(c,d)** Average gene-tissue fine-mapping power across 100 simulations using various fine-mapping methods (see legend) across *eQTL* sample sizes (x-axis) at PIP=0.5 **(c)** and PIP=0.9 **(d)**. Error bars denote 95% confidence intervals. This supplementary figure is similar to Figure 1, except it only shows simulation results for TGFM, FOCUS-TG, FOCUS, FOCUS-TG (genotype intercept), and FOCUS (genotype intercept). The (genotype intercept) suffix refers to running FOCUS with the “intercept” parameter set to True; the “intercept” parameter is set to False by default (the default version of FOCUS is used in the primary analysis, e.g. Figure 1). The “intercept” parameter allows FOCUS to model non-mediated genetic effects via single genotype intercept term shared across all variants. More specifically, the genotype intercept parameter estimates and removes the average amount of TWAS signal attributed to non-mediated genetic variants at a locus. All genes at the locus are therefore assumed to capture the same amount of signal from non-mediated variants; TGFM and cTWAS allow the strength of non-mediated genetic effects to vary across the locus.

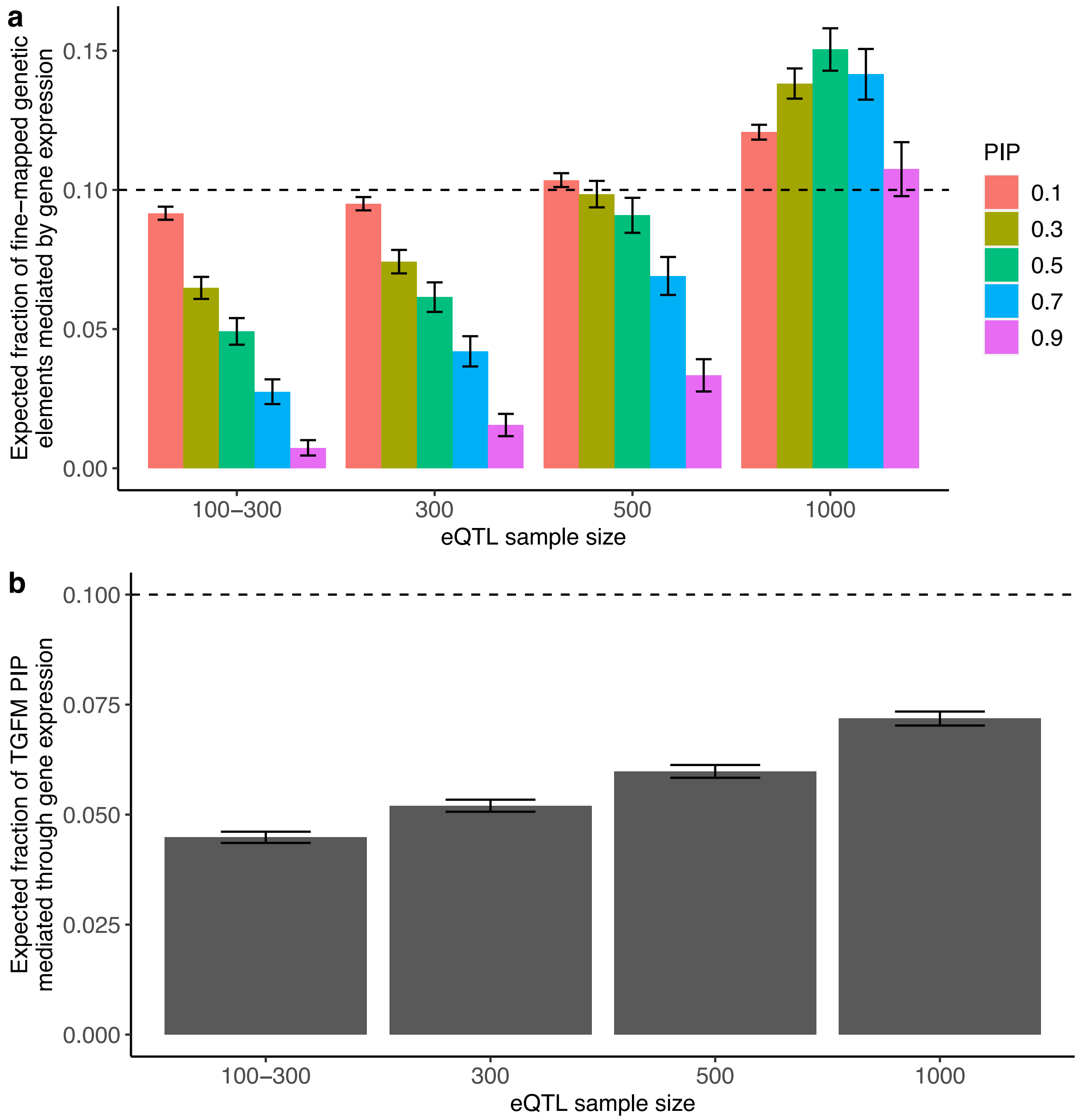


**Supplementary Figure 24: Evaluating bias in estimating the proportion of genetic elements that are gene-tissue pairs in simulations.** The horizontal dashed line in plot **(a)** and **(b)** represents the simulated, true proportion of genetic elements that are gene-tissue pairs. **(a)** We report the average proportion of fine-mapped genetic elements across 100 simulations that are gene-tissue pairs (y-axis) at various PIP thresholds (see legend) across various eQTL sample sizes (x-axis). This approach yielded either upward or downward biased estimates of the true proportion depending on eQTL sample size and PIP threshold, reflecting differential discovery power of non-mediated variants and gene-tissue pairs as a function of both eQTL sample size and PIP threshold. **(b)** We report the average expected proportion of causal genetic elements that are gene-tissue pairs (y-axis) across 100 simulations (computed by summing PIPs across genetic elements), shown at various eQTL sample sizes (x-axis). This approach yielded conservative estimates of the true proportion, becoming less conservative at larger eQTL sample sizes, suggesting that this statistic can provide a conservative lower bound on the true proportion of causal genetic elements that are gene-tissue pairs. Error bars denote 95% confidence intervals.

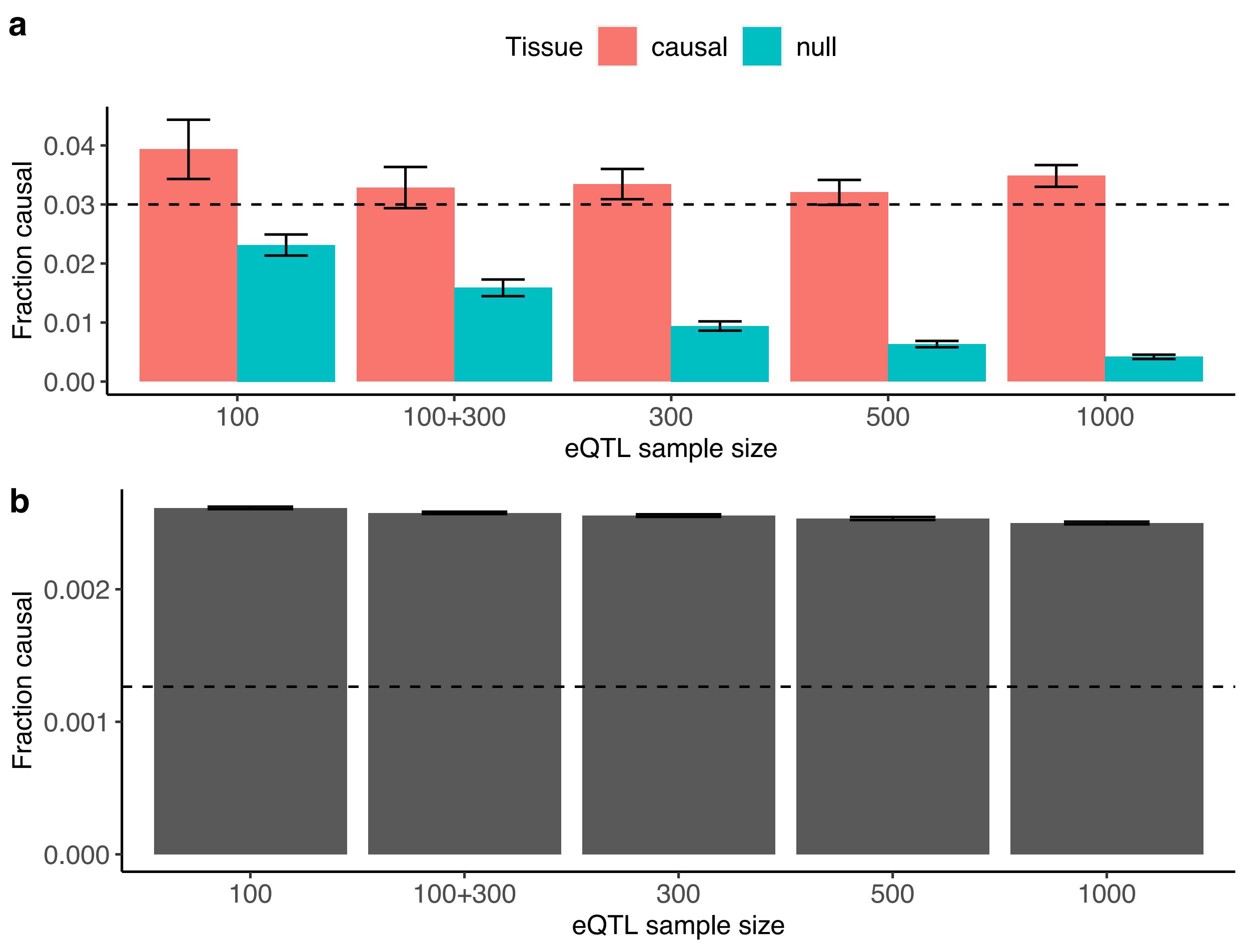


**Supplementary Figure 25: Unbiasedness of the prior causal probabilities inferred by the TGFM tissue-specific prior in simulations. (a)** The average prior causal probabilities for (gene-tissue pairs) involving causal and non-causal (null) tissues (see legend) across 100 simulations at various eQTL sample sizes (x-axis). Dashed horizontal line denotes the true (simulated) fraction of causal gene-tissue pairs from a causal tissue. The true (simulated) fraction of causal gene-tissue pairs from a null tissue is zero. **(b)** The average prior causal probabilities for non-mediated variants across 100 simulations at various eQTL sample sizes (x-axis). Dashed horizontal line denotes the true (simulated) fraction of causal non-mediated variants.

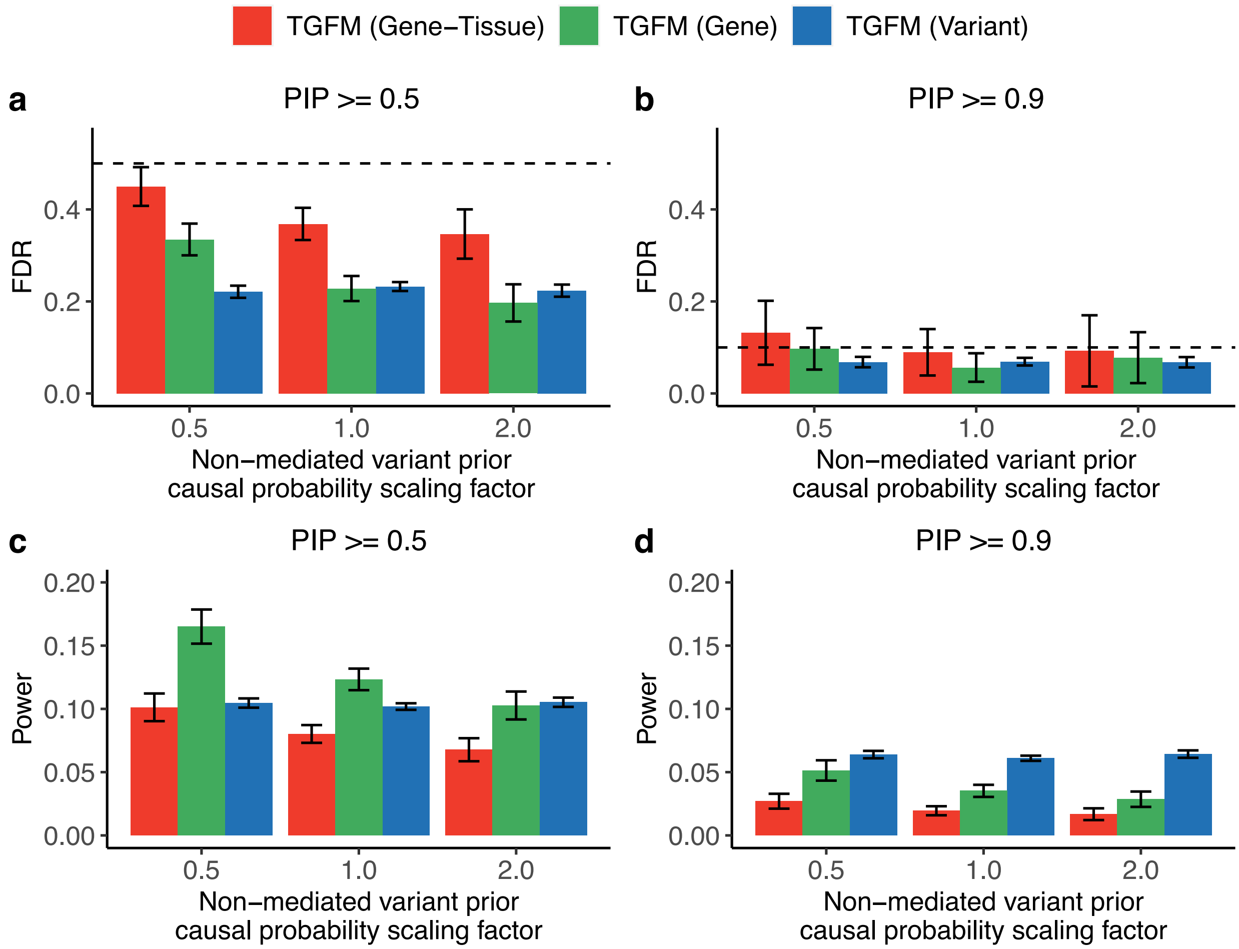


**Supplementary Figure 26*: Calibration and power of fine-mapping different classes of genetic elements with TGFM after scaling the prior causal probabilities on non-mediated variants.* (a,b)** Average fine-mapping FDR across 100 simulations using TGFM for different classes of genetic elements (see legend) as a function of the scaling factor applied to the TGFM tissue-specific prior causal probability on non-mediated variants (x-axis) at PIP=0.5 **(a)** and PIP=0.9 **(b)** at an eQTL sample size of 500. Dashed horizontal line denotes 1 – PIP threshold (see main text). **(c,d)** Average fine-mapping power across 100 simulations using TGFM for different classes of genetic elements (see legend) as a function of the scaling factor applied to the TGFM tissue-specific prior causal probability on non-mediated variants (x-axis) at PIP=0.5 **(c)** and PIP=0.9 **(d)** at an eQTL sample size of 500. Error bars denote 95% confidence intervals. This supplementary figure is similar to Figure 2, except it shows results at a fixed eQTL sample of 500, and varies the scaling factor applied to the TGFM tissue-specific prior causal probability on non-mediated variants.

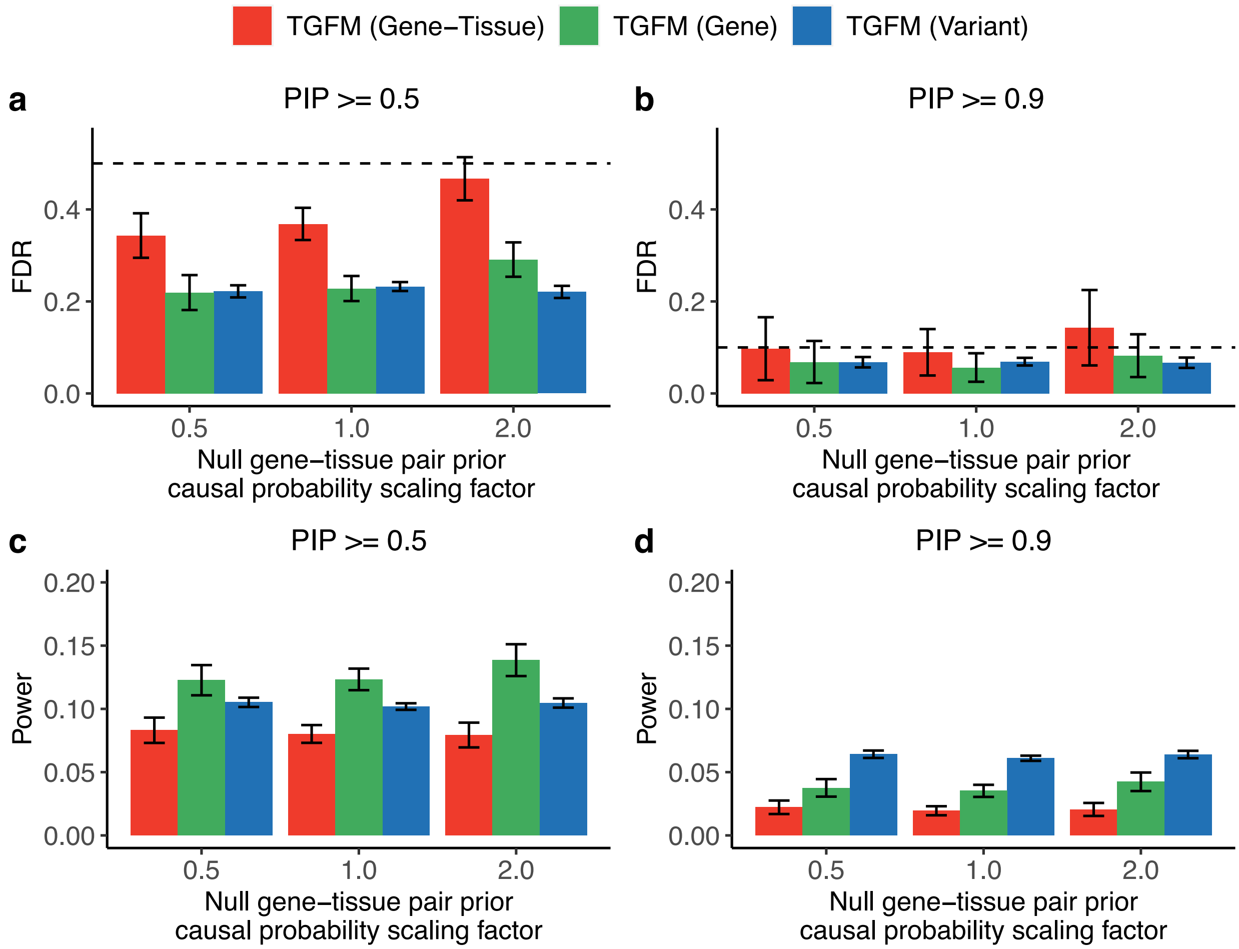


**Supplementary Figure 27*:* Calibration and power of fine-mapping different classes of genetic elements with TGFM after scaling the prior causal probabilities on gene-tissue pairs of null tissues. (a,b)** Average fine-mapping FDR across 100 simulations using TGFM for different classes of genetic elements (see legend) as a function of the scaling factor applied to the TGFM tissue-specific prior causal probability on gene-tissue pairs from null (non-causal) tissues (x-axis) at PIP=0.5 **(a)** and PIP=0.9 **(b)** at an eQTL sample size of 500. Dashed horizontal line denotes 1 – PIP threshold (see main text). **(c,d)** Average fine-mapping power across 100 simulations using TGFM for different classes of genetic elements (see legend) as a function of the scaling factor applied to the TGFM tissue-specific prior causal probability on gene-tissue pairs from null (non-causal) tissues (x-axis) at PIP=0.5 **(c)** and PIP=0.9 **(d)** at an eQTL sample size of 500. Error bars denote 95% confidence intervals. This supplementary figure is similar to Figure 2, except it shows results at a fixed eQTL sample of 500, and varies the scaling factor applied to the TGFM tissue-specific prior causal probability on gene-tissue pairs from null (non-causal) tissues.

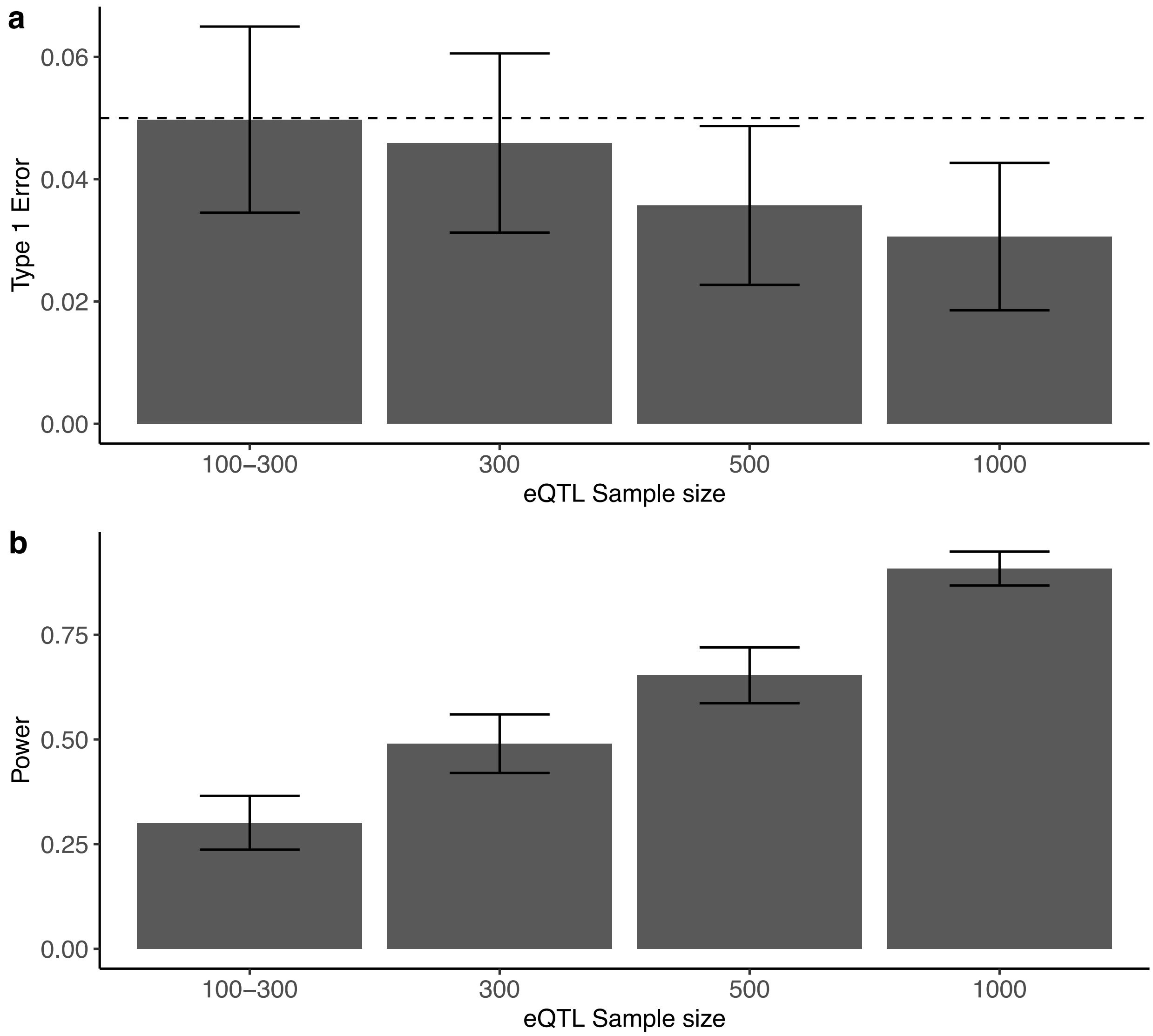


**Supplementary Figure 28: Evaluating power and type 1 error of identifying disease-critical tissues using the TGFM tissue-specific prior in simulations. (a)** Average type 1 error across 100 simulations for identifying disease-critical tissues using the TGFM tissue-specific prior at a p-value threshold of 0.05 (y-axis) at various eQTL sample sizes (x-axis). Error bars denote 95% confidence intervals. **(b)** Average power across 100 simulations for identifying disease-critical tissues using the TGFM tissue-specific prior at a p-value threshold of 0.05 (y-axis) at various eQTL sample sizes (x-axis). Error bars represent 95% confidence intervals.

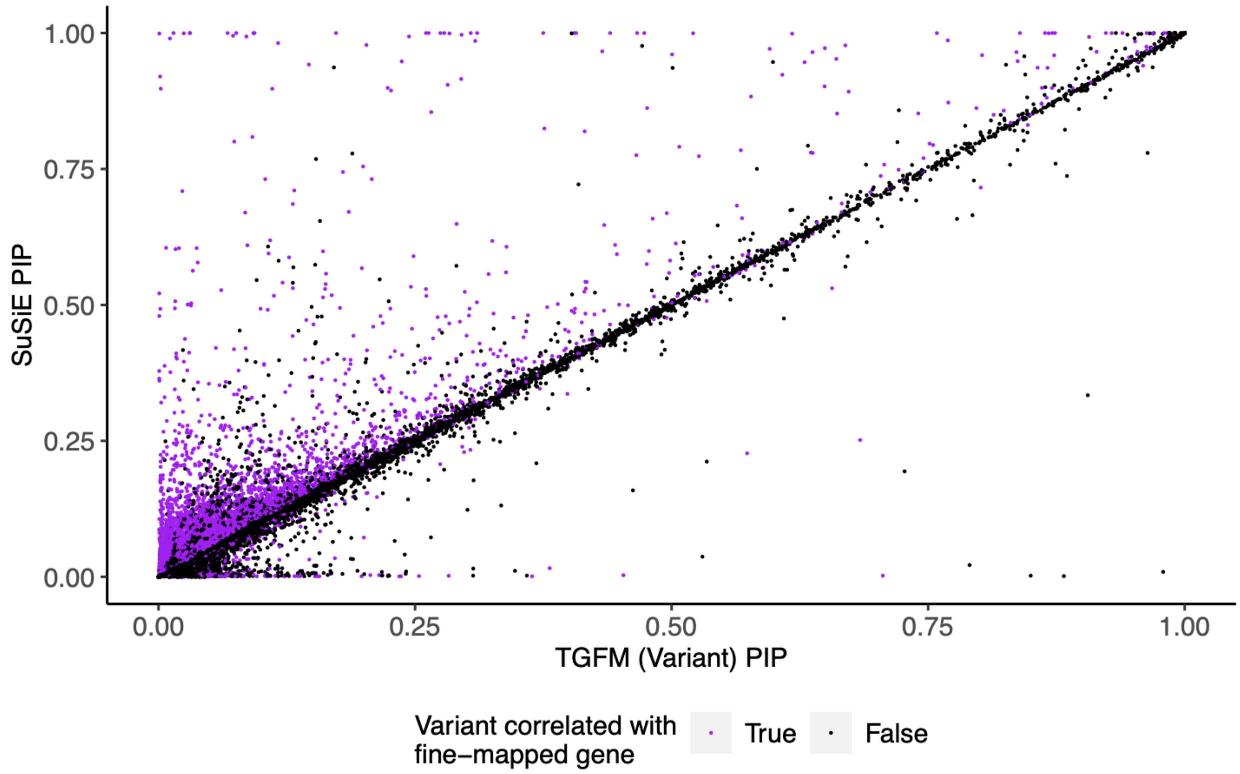


**Supplementary Figure 29: Direct comparison of TGFM (Variant) PIPs and SuSiE PIPS in simulations.** For each non-mediated variant analyzed across 20 default simulations at eQTL sample size of 500, we plot the TGFM (Variant) PIP (x-axis) and the SuSiE PIP (y-axis) colored by whether the variant had absolute correlation > 0.25 with a fine-mapped (PIP > 0.05) gene-tissue pair. The correlation across all variants was 0.975 (slope of regression of SuSiE PIP on TGFM (Variant) PIP: 1.016), the correlation across all variants not correlated with a fine-mapped gene-tissue pair (black points) was 0.993 (slope: 0.999), and the correlation across all variants correlated with a fine-mapped gene-tissue pair (purple points) was 0.889 (slope: 1.160). For strictly visualization purposes, we only visualize a random subset of 100,000 variants with both SuSiE PIP < 0.01 and TGFM (Variant) PIP < 0.01.

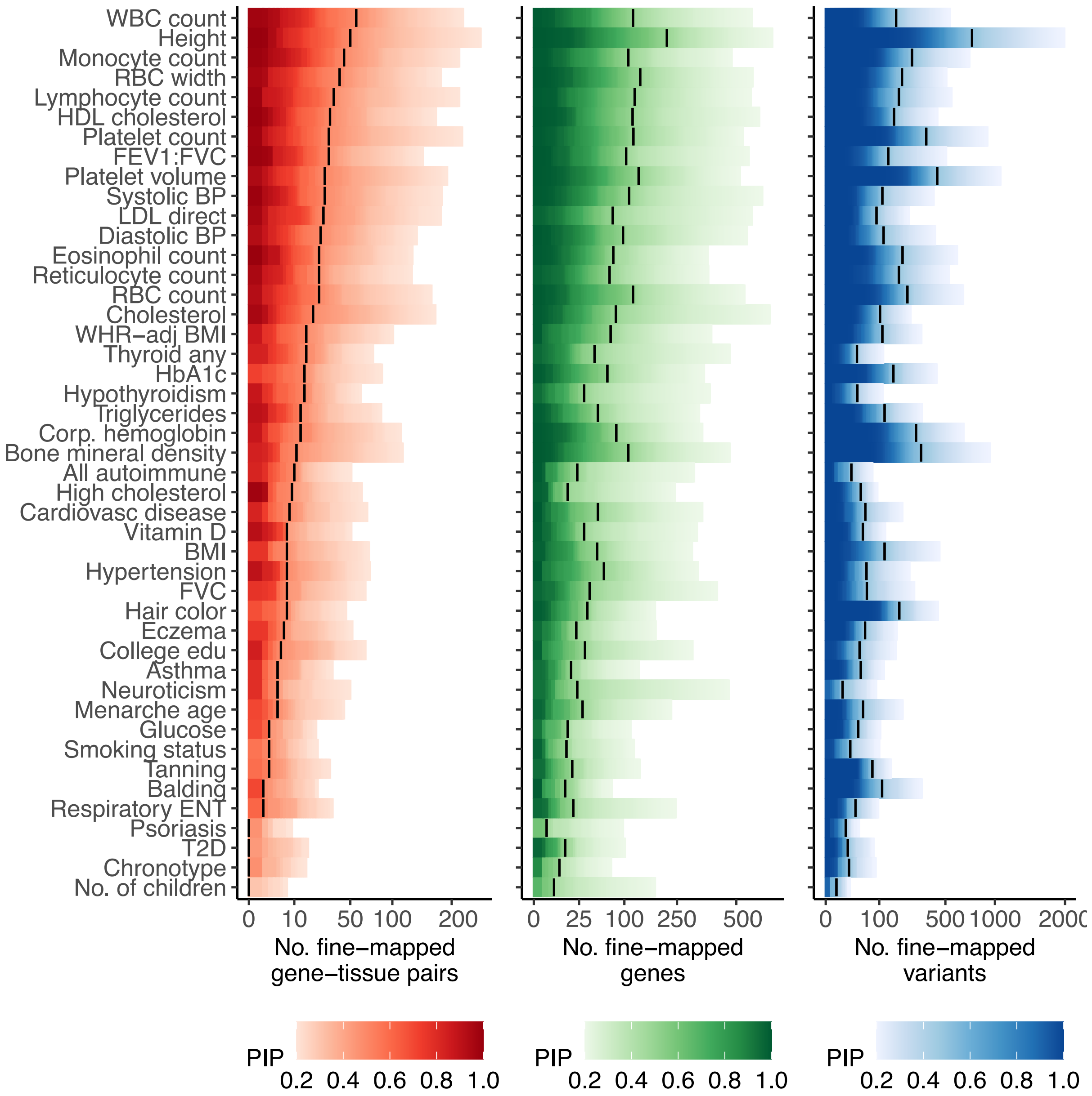


**Supplementary Figure 30: Summary of fine-mapping genetic elements with TGFM for all 45 UK Biobank disease and traits.** We report the number of **(a)** Gene-tissue pairs, **(b)** Genes, and **(c)** (non-mediated) Variants fine-mapped using TGFM (x-axis; square root scale) across 45 UK Biobank traits (y-axis) at various PIP thresholds ranging from 0.2 to 1.0 (color-bars). Vertical black lines denote the number of genetic elements fine-mapped at PIP=0.5. This supplementary figure is similar to Figure 3, except it shows results across all 45 traits, and the x-axis and y-axis are flipped.

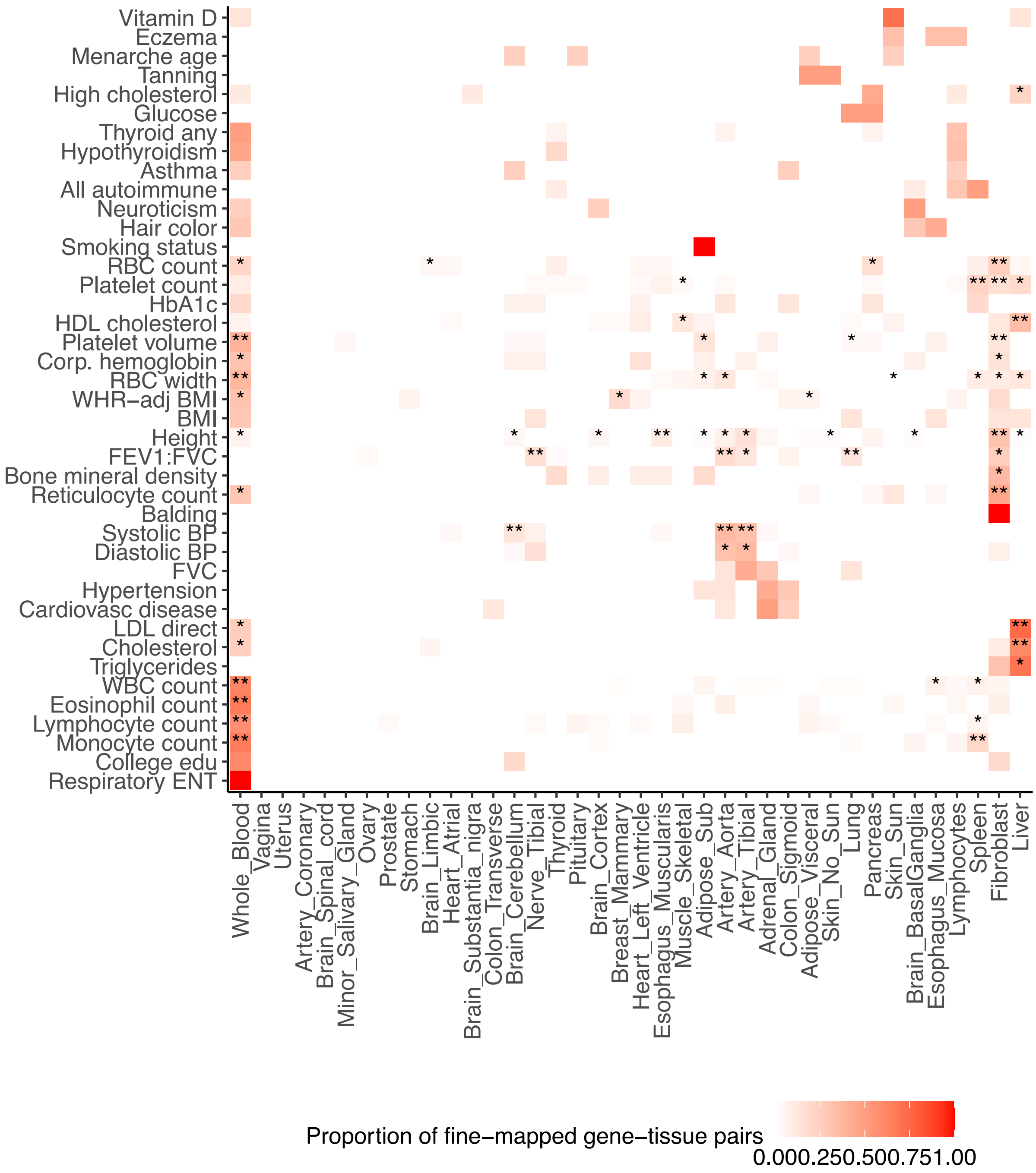


**Supplementary Figure 31: Proportion of fine-mapped gene-tissue pairs in each tissue for all traits.** Proportion of fine-mapped gene-tissue pairs in each tissue (x-axis) for 45 traits (y-axis). Proportions for each trait were calculated by counting the number of gene-tissue pairs with TGFM PIP > 0.5 in each tissue and normalizing the counts across tissues. Statistical significance of each tissue-disease pair by applying genomic bootstrapping to the TGFM tissue-specific prior. ** represents FDR $\leq 0.05$, and $*$ represents FDR $\leq 0.2$. Numerical results are reported in Supplementary Table 9.

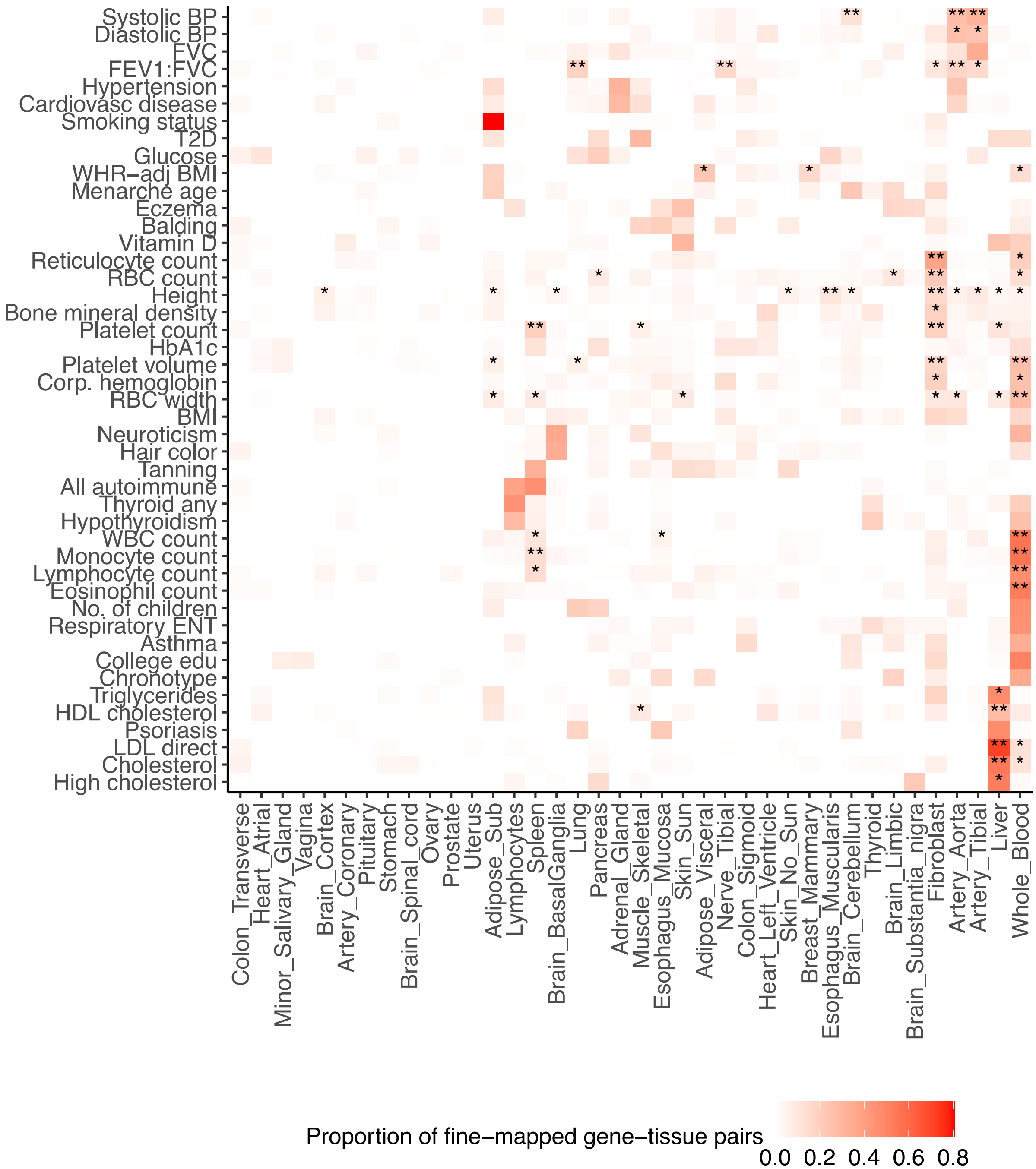


**Supplementary Figure 32: Proportion of fine-mapped gene-tissue pairs in each tissue for all traits at alternative threshold (PIP $> 0.1$).** Proportion of fine-mapped gene-tissue pairs in each tissue (x-axis) for 45 traits (y-axis). Proportions for each trait were calculated by counting the number of gene-tissue pairs with TGFM PIP > 0.1 in each tissue and normalizing the counts across tissues. Statistical significance of each tissue-disease pair by applying genomic bootstrapping to the TGFM tissue-specific prior. ** represents FDR $\leq 0.05$, and $*$ represents FDR $\leq 0.2$.

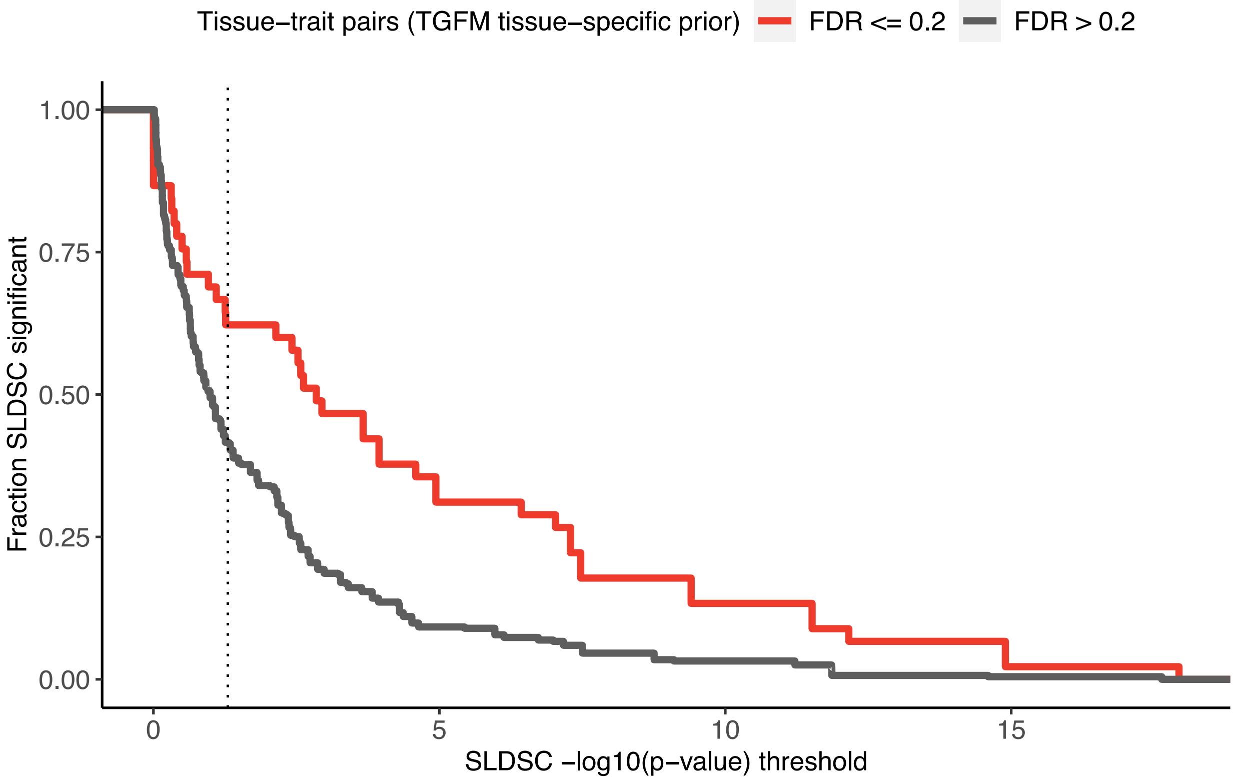


**Supplementary Figure 33: S-LDSC analyses using chromatin data at alternative TGFM tissue-specific prior FDR threshold.** Proportion of tissue-trait pairs identified as statistically significant in S-LDSC analyses using chromatin data (y-axis) over a range of S-LDSC significance thresholds (x-axis) for all 45 analyzed traits. Tissue-trait pairs are stratified according to significance (FDR $\leq$ 0.2) in the TGFM tissue-specific prior. This supplementary figure is similar to Figure 4b, except it shows results across at TGFM tissue-specific prior threshold of 0.2 instead of 0.05.

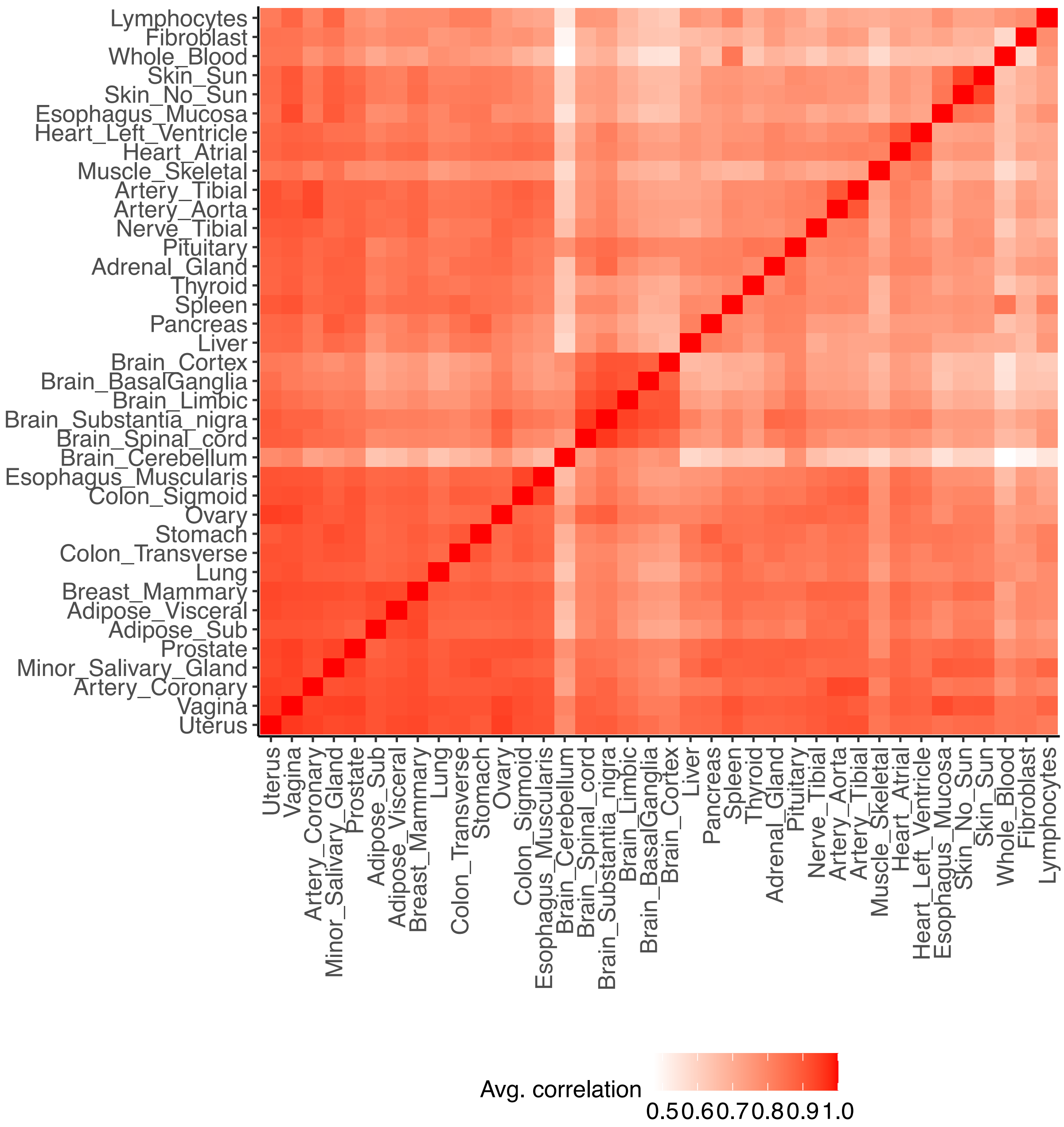


**Supplementary Figure 34: Average correlation in cis-predicted gene expression between pairs of GTEx tissues.** We report the average correlation in cis-predicted gene expression between each pair of GTEx tissues across all genes included in TGFM. For a given pair of tissues, we only compute the correlation across genes with a cis-predicted expression model in both tissues. We observed that several of the low sample size tissues (e.g. uterus, vagina, artery coronary, minor salivary gland, prostate; Supplementary Table 6) have high correlation with nearly all other tissues. We assume this is a technical artifact of low sample size tissues being limited to discovering very large eQTL effect size variants (due to statistical power), which are known to be less tissue-specific than small eQTL effect size variants.

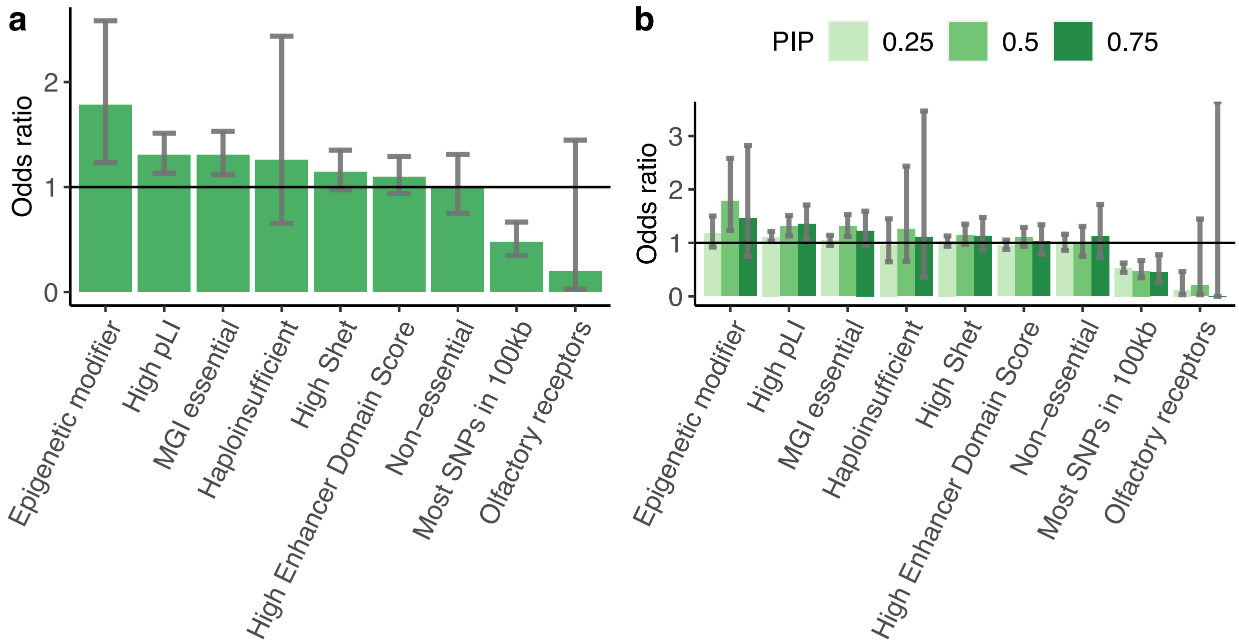


**Supplementary Figure 35: Enrichment of fine-mapped TGFM genes within non-disease-specific gene sets. (a)** Enrichment of genes with TGFM (Gene) PIP > 0.5 within non-disease-specific gene sets meta-analyzed over 16 independent traits. Error bars represent 95% confidence intervals. Odds ratios and standard errors on the odds ratio were computed using logistic regression. **(b)** Enrichment of genes with TGFM (Gene) PIP > 0.25, 0.5, and 0.75 (see legend) within non-disease-specific gene sets meta-analyzed over 16 independent traits. Error bars represent 95% confidence intervals. Odds ratios and standard errors on the odds ratio were computed using logistic regression. Numerical results reported in Supplementary Table 14.

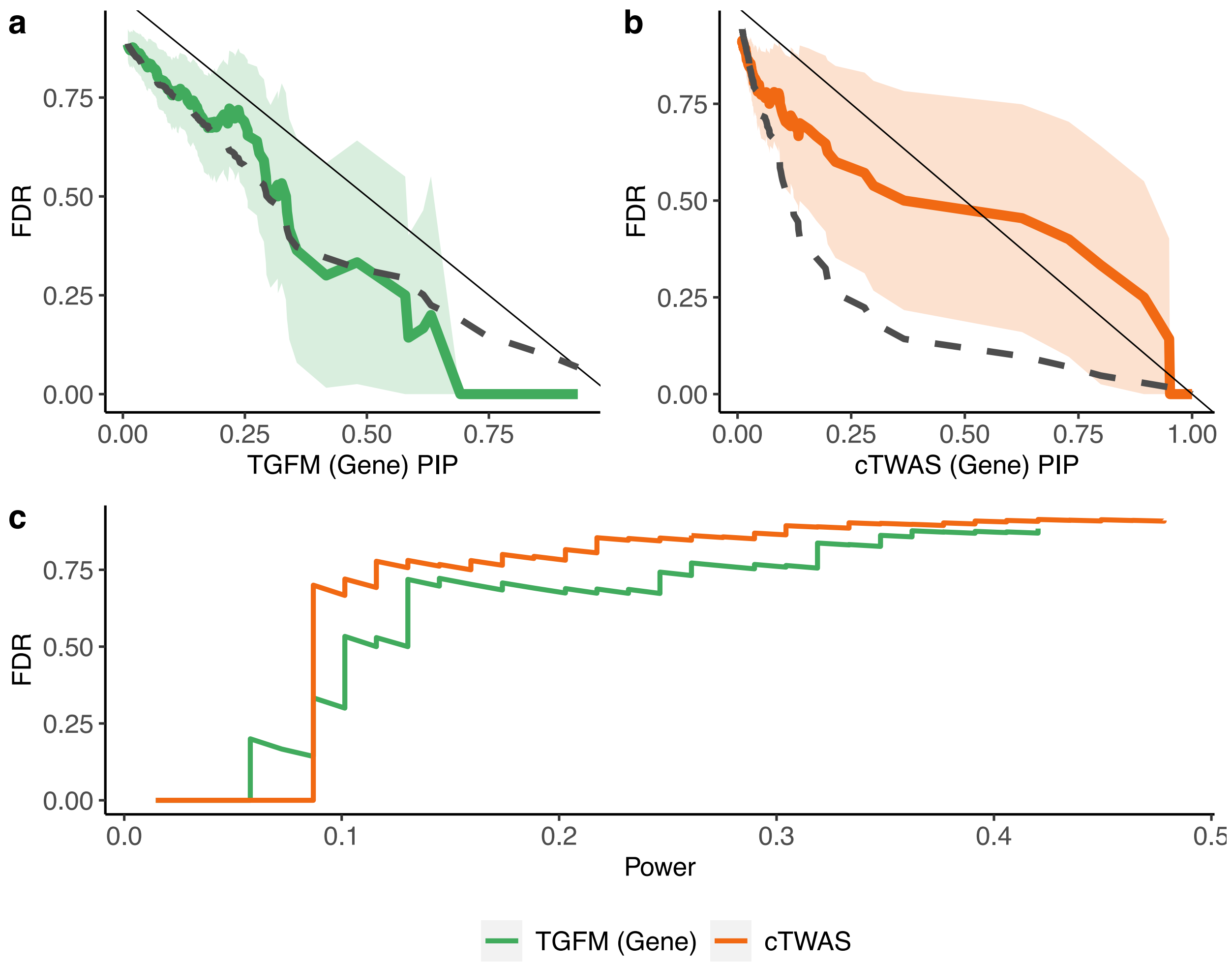


**Supplementary Figure 36: Comparison of TGFM (Gene) and cTWAS calibration and power using silver standard gene set of 69 known LDL cholesterol genes. (a-b)** Empirical FDR (y-axis) using silver-standard gene set of 69 known LDL cholesterol genes at PIP greater than or equal to a range of PIP thresholds (x-axis) for TGFM (Gene) **(a)** and cTWAS applied to GTEx liver **(b)**. Light shading denotes 95% confidence intervals. Black dashed line denotes (1 – average PIP), a less conservative choice than (1 – PIP threshold). **(c)** Average gene fine-mapping power (x-axis) at a specific level of FDR (y-axis) based on silver-standard gene set of 69 known LDL cholesterol genes for TGFM (gene) and cTWAS applied to GTEx liver (see legend). PIPs for cTWAS applied to GTEx liver extracted from Supplementary Table 2 of ref. [26].

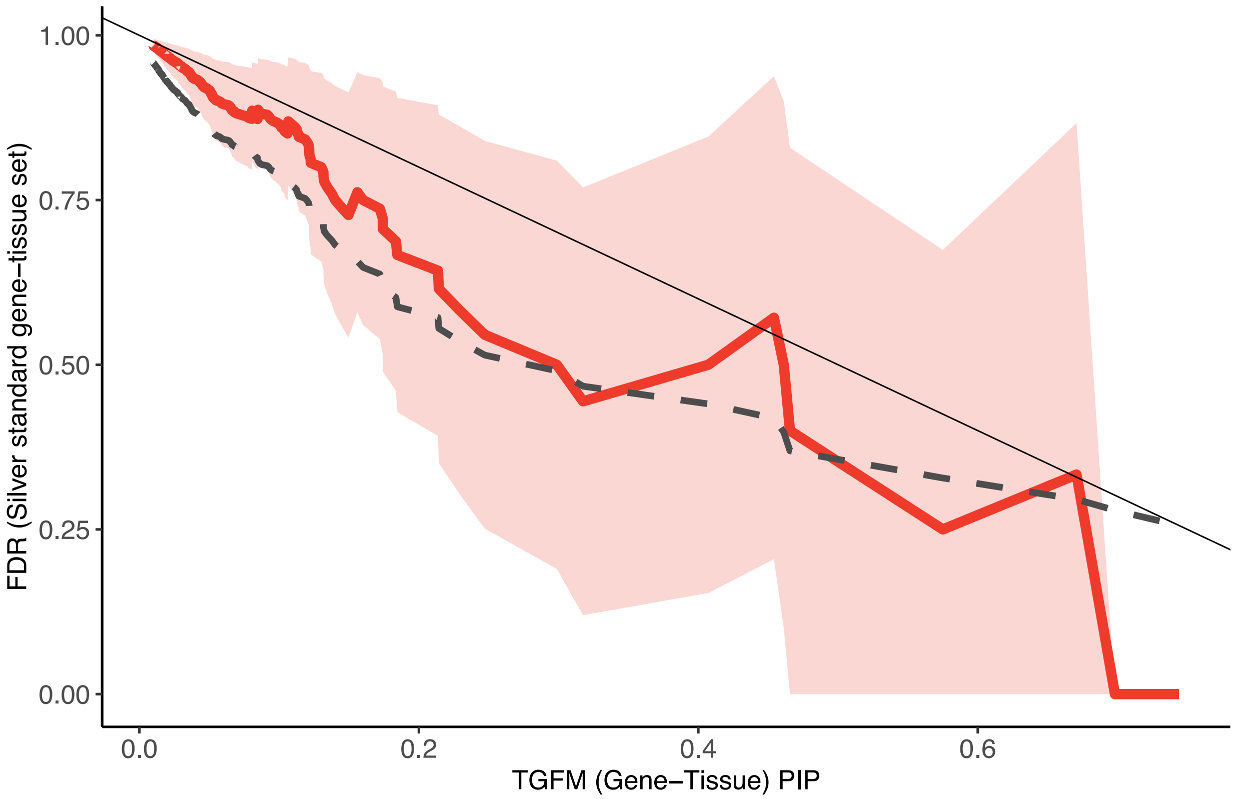


**Supplementary Figure 37: Calibration of TGFM (Gene-Tissue) using silver standard gene set of 69 known LDL cholesterol genes.** Empirical FDR (y-axis) at PIP greater than or equal to a range of PIP thresholds (x-axis) for TGFM (Gene-Tissue) using the silver-standard gene set of 69 known LDL cholesterol genes while assuming that all 69 known LDL cholesterol genes act in liver tissue; specifically, we used the 69 LDL cholesterol genes in liver as the positive set, and used both the 69 LDL cholesterol genes in any tissue other than liver and the nearby genes in any tissue as the negative set. Light shading denotes 95% confidence intervals. Black dashed line denotes (1 – average PIP), a less conservative choice than (1 – PIP threshold).

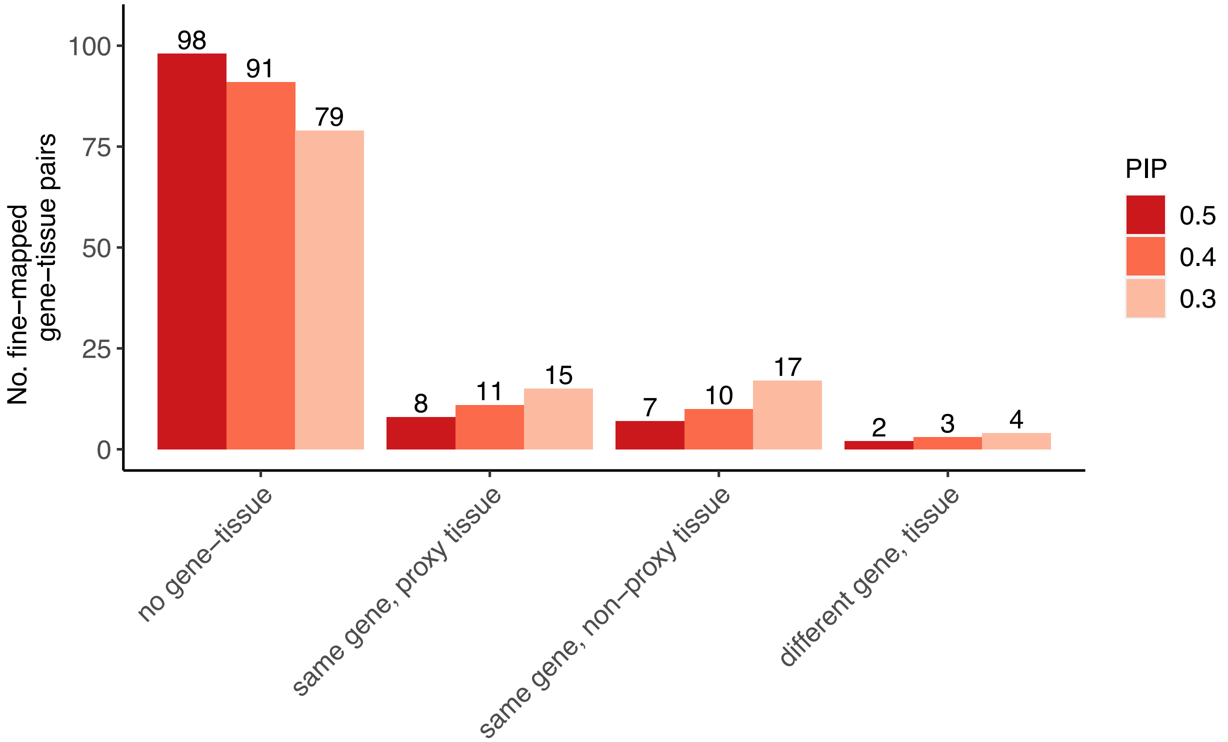


**Supplementary Figure 38: Per-category number of loci identified in tissue ablation analysis at various TGFM PIP thresholds.** Results of tissue ablation analysis of 115 gene-tissue pairs for 18 representative traits for which the ablated tissue was prioritized by TGFM (PIP > 0.5) in the primary analysis. We report the number of loci in the tissue ablation analysis with no gene-tissue pair prioritized by TGFM (PIP > PIP threshold); a gene-tissue pair prioritized by TGFM (PIP > PIP threshold) corresponding to the same gene and the best proxy tissue (see Methods); a gene-tissue pair prioritized by TGFM (PIP > PIP threshold) corresponding to the same gene and a non-proxy tissue; or a gene-tissue pair prioritized by TGFM (PIP > PIP threshold) corresponding to a different gene. These results are shown at various PIP thresholds (see legend). This figure is identical to Figure 5a, except it shows results at various PIP thresholds instead of just PIP > 0.5.

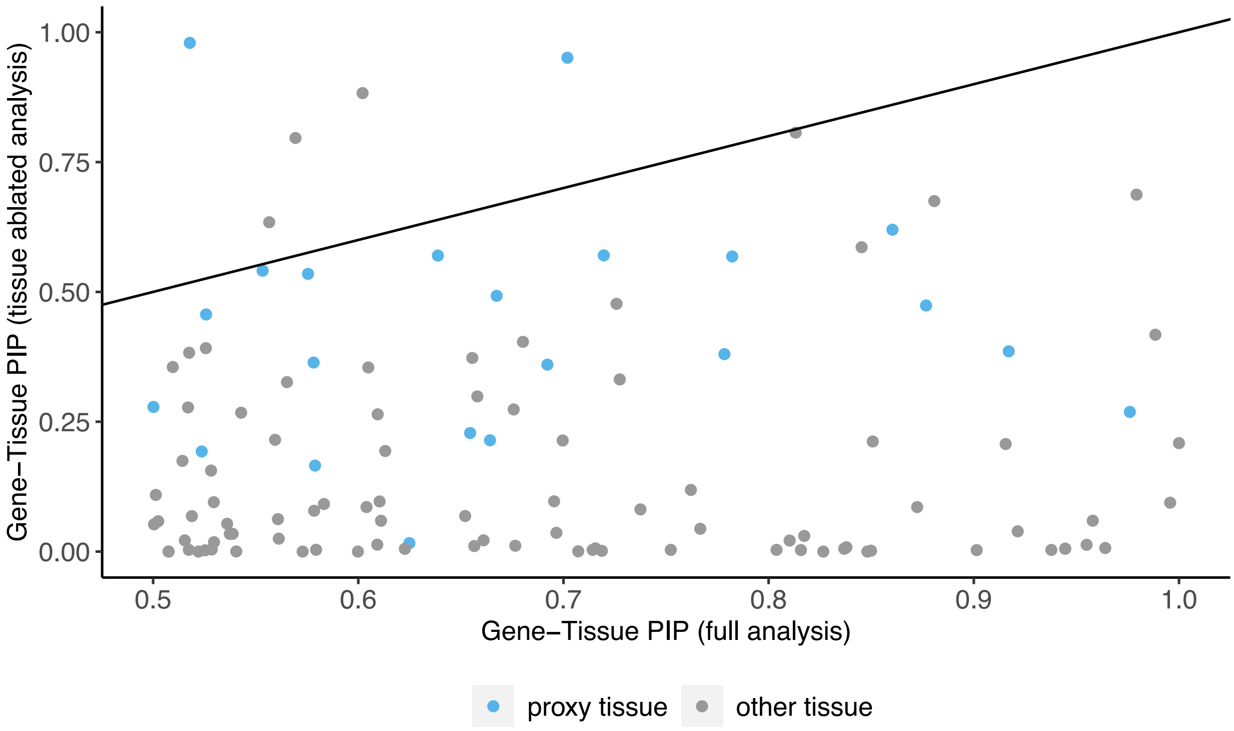


**Supplementary Figure 39: Distribution of gene-tissue PIPs in tissue ablation analysis.** In the tissue ablation analysis, we considered 115 gene-tissue pairs from 18 representative traits for which the ablated tissue was prioritized for TGFM (PIP > 0.5) in the primary analysis. We report the TGFM (Gene-Tissue) PIP in the primary analysis (x-axis) and the maximum (across tissues) TGFM (Gene-Tissue) PIP in the tissue ablation analysis (y-axis). Genes (points) are colored by whether the tissue with largest PIP in the tissue ablation analysis was a proxy tissue with respect to the ablated tissue.

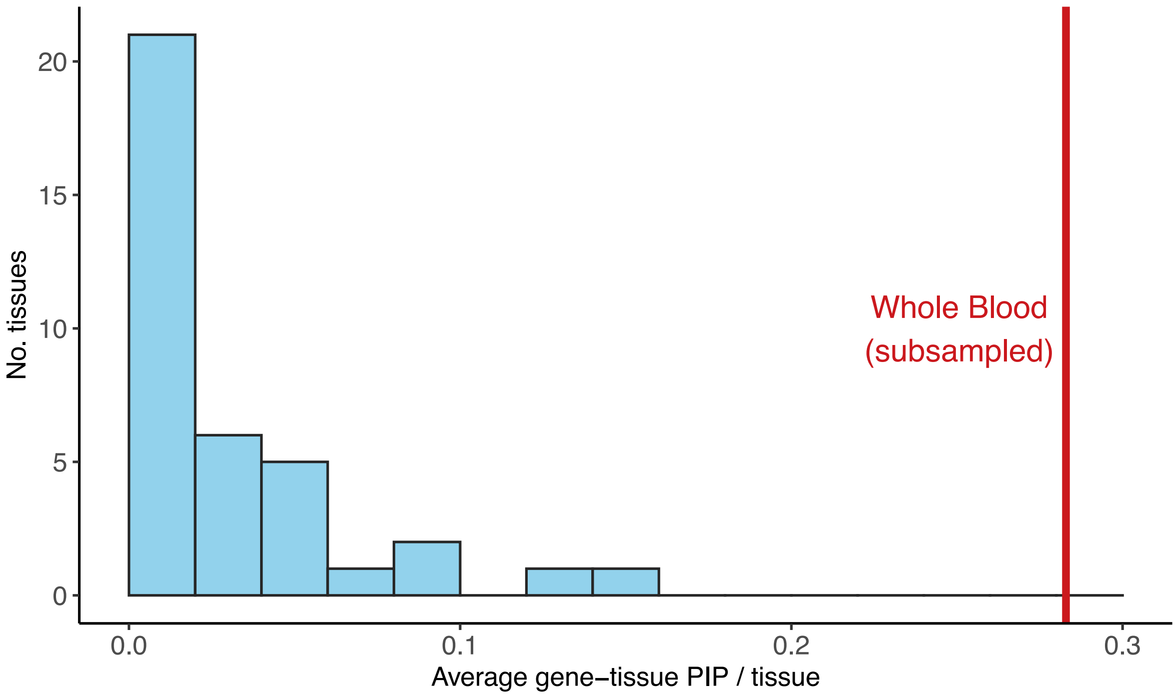


**Supplementary Figure 40: Replacement of whole blood tissue with down-sampled whole blood tissue.** Results of replacing GTEx whole blood (*N*=320) with down-sampled whole blood (*N*=113) for 62 gene-trait pairs for 18 representative traits that TGFM fine-mapped for GTEx whole blood (PIP > 0.5) in the primary analysis. The vertical red line denotes the average PIP in down-sampled whole blood, and the histogram summarizes the average PIP of each GTEx tissue (excluding whole blood). The 18 representative traits consist of the 16 independent traits (Figure 3) and two additional, interesting traits (All autoimmune and Vitamin D levels).

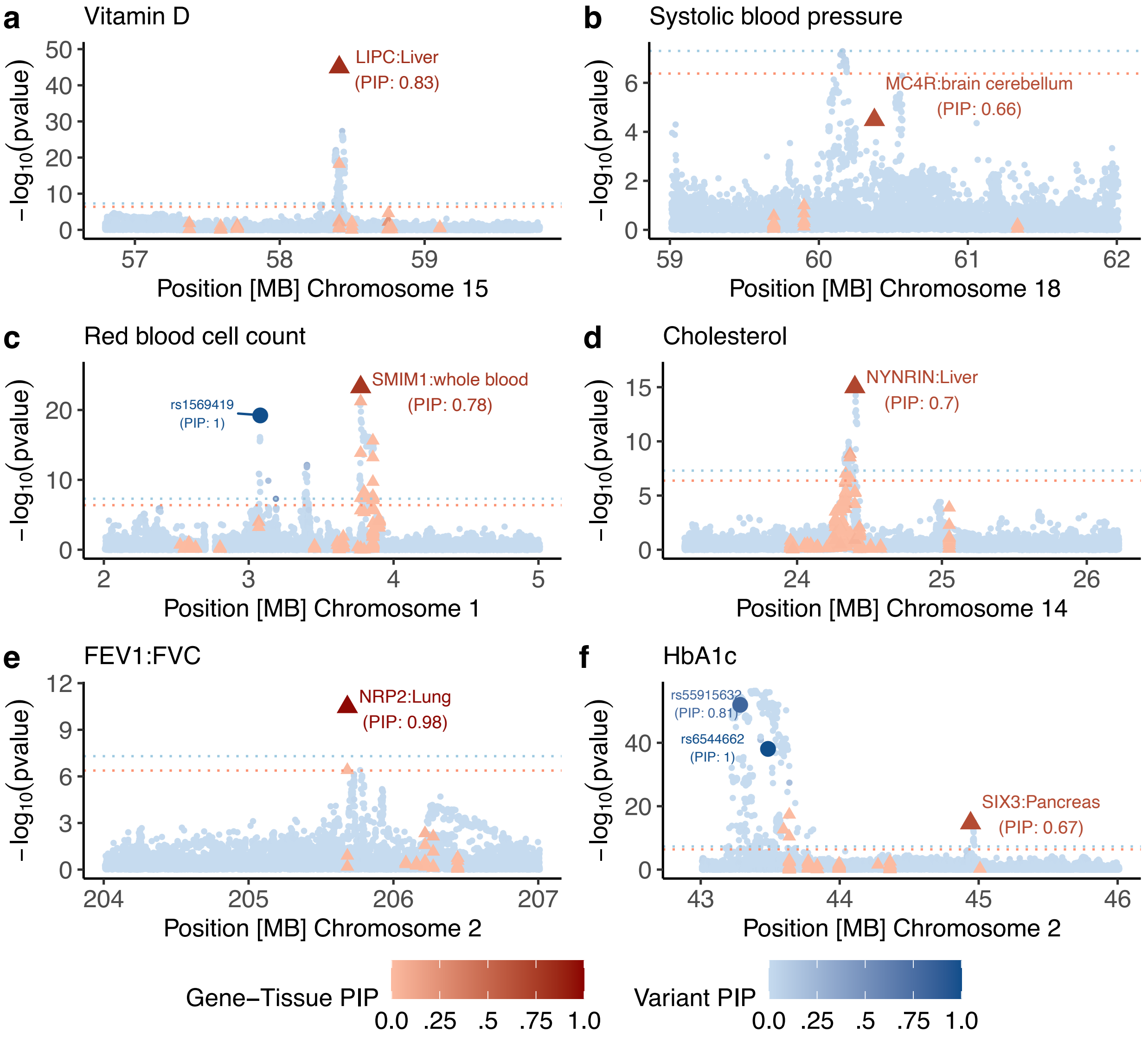


**Supplementary Figure 41: Additional examples of fine-mapped gene-tissue-disease triplets identified by TGFM.** We report 6 example loci for which TGFM fine-mapped a gene-tissue pair (PIP > 0.5). In each example we report the marginal GWAS and TWAS association -$\log_{10}$ p-values (y-axis) of non-mediated variants (blue circles) and gene-tissue pairs (red triangles). Marginal TWAS association -$\log_{10}$ p-values were calculated by taking the median -$\log_{10}$ TWAS p-value across the 100 sets of sampled cis-predicted expression models for each gene-tissue pair. The genomic position of each gene-tissue pair (x-axis) was based on the gene's TSS. The color shading of each variant and gene-tissue pair was determined by its TGFM PIP. Any genetic element with TGFM PIP > 0.5 was made larger in size. Dashed horizontal blue and red lines represent GWAS significance ($5 \times 10^{-8}$) and TWAS significance ($4.2 \times 10^{-7}$) thresholds, respectively.

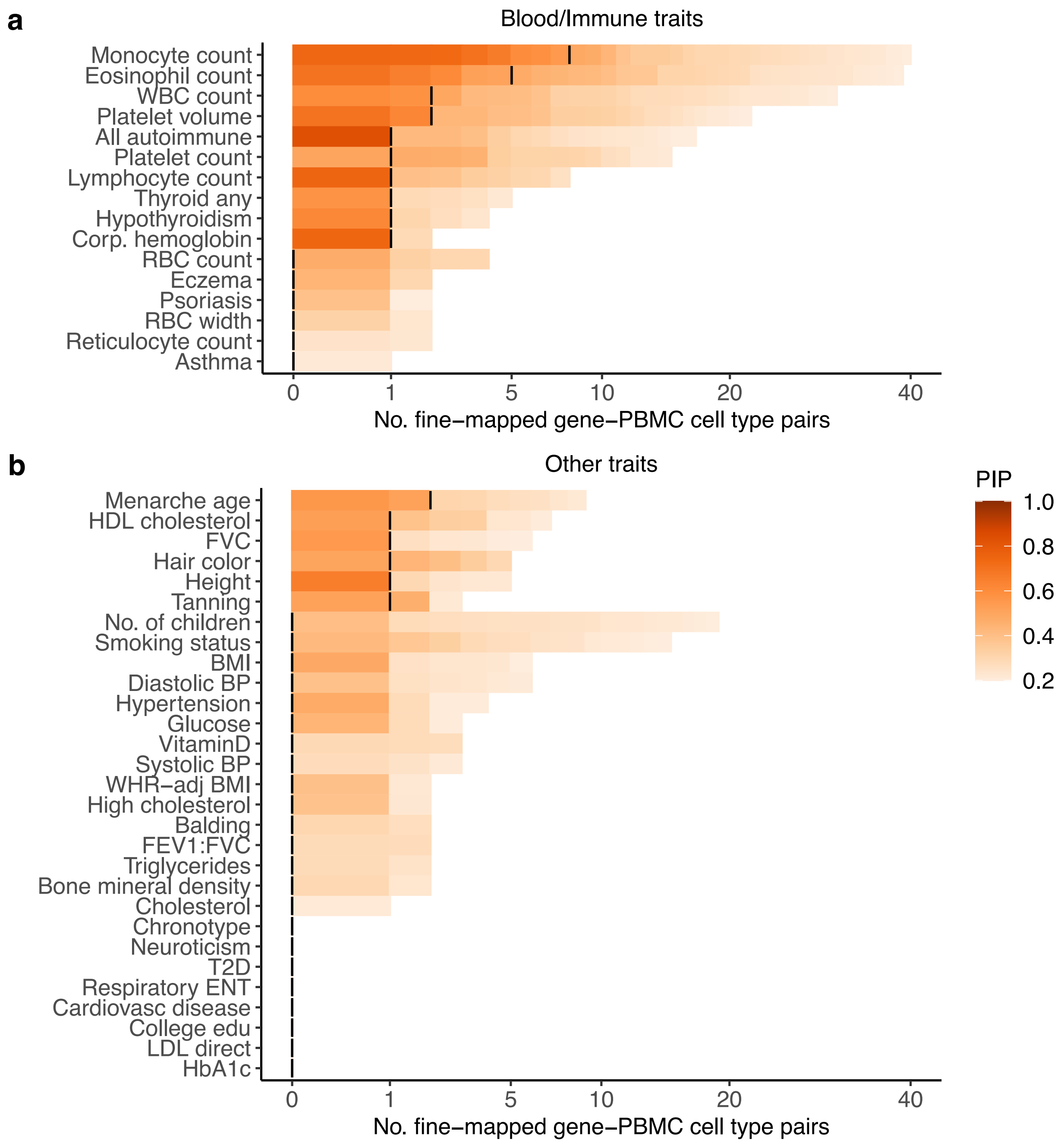


**Supplementary Figure 42: Summary of fine-mapping gene-PBMC cell type pairs with TGFM for all 45 UK Biobank diseases and traits.** Number of gene-PBMC cell type pairs fine-mapped using TGFM (x-axis; square root scale) across 18 representative UK Biobank traits (y-axis) at various PIP thresholds ranging from 0.2 to 1.0 (color-bar), distinguishing between **(a)** autoimmune diseases and blood cell traits and **(b)** non-blood-related traits. Vertical black lines denote the number of gene-PBMC cell type pairs fine-mapped at PIP=0.5. Numerical results are reported in Supplementary Table 20. This supplementary figure is similar to Figure 7a-b, except it shows results across all 45 traits, and the x-axis and y-axis are flipped.

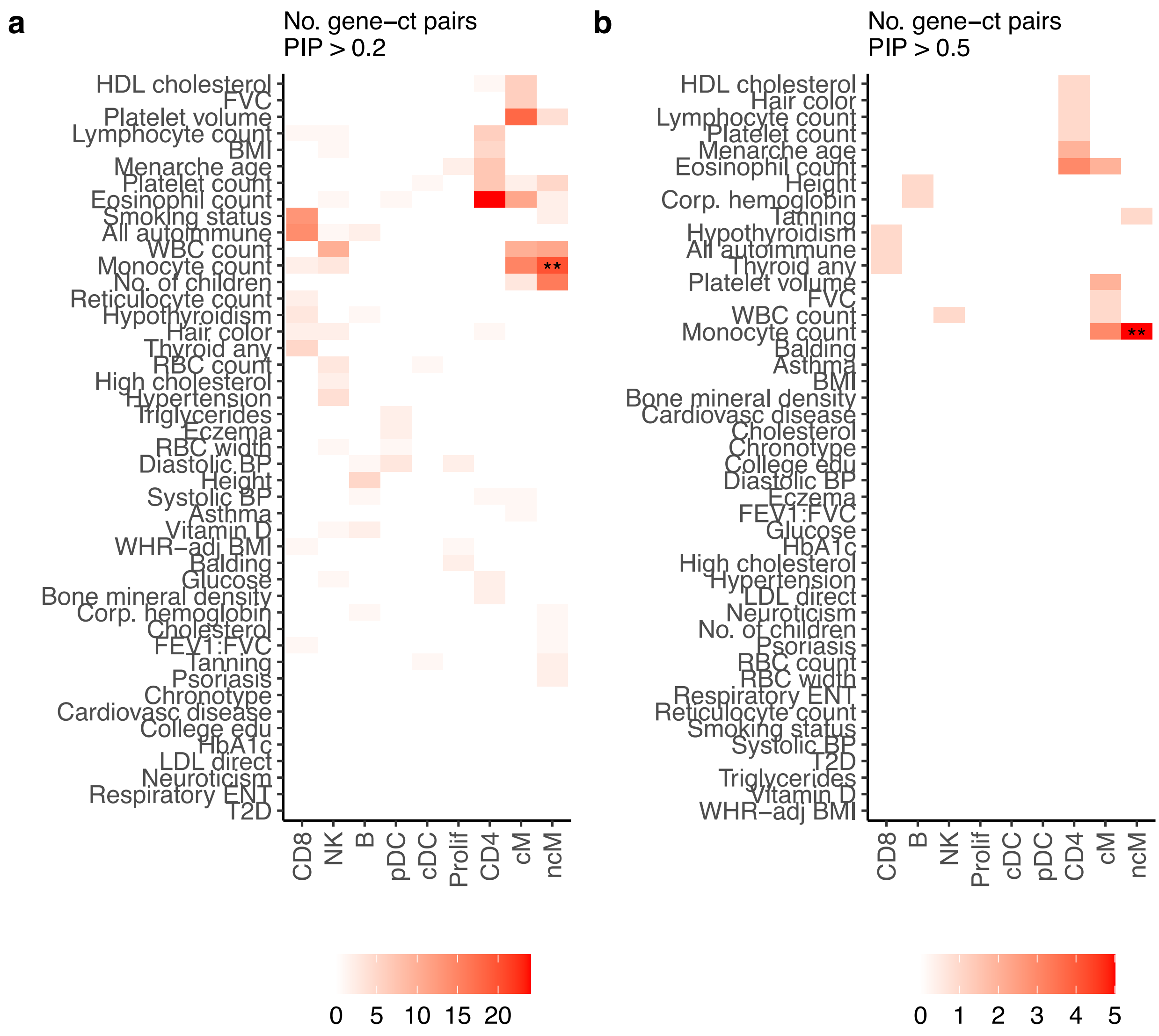


**Supplementary Figure 43: Number of gene-PBMC cell type pairs corresponding to each trait-PBMC cell type across 45 UK Biobank diseases and traits.** The number of fine-mapped gene-PBMC cell type pairs at PIP > 0.2 **(a)** and PIP > 0.5 **(b)** corresponding to each trait-PBMC cell type pair across 45 UK Biobank diseases and traits. Numerical results are reported in Supplementary Table 20. Statistical significance of each trait-PBMC cell type pair by applying genomic bootstrapping to the TGFM tissue-specific prior. ** represents FDR $\leq 0.05$, and $*$ represents FDR $\leq 0.2$.

### *References*


1. Connelly, P. W. The role of hepatic lipase in lipoprotein metabolism. *Clin. Chim. Acta* **286**, 243–255 (1999).
2. Santamarina-Fojo, S., Haudenschild, C. & Amar, M. The role of hepatic lipase in lipoprotein metabolism and atherosclerosis. *Curr. Opin. Lipidol.* **9**, 211–219 (1998).
3. Manousaki, D. *et al.* Genome-wide association study for vitamin D levels reveals 69 independent loci. *Am. J. Hum. Genet.* **106**, 327–337 (2020).
4. Li, Y. *et al.* Association of changes in lipid levels with changes in vitamin D levels in a real-world setting. *Sci. Rep.* **11**, 1–7 (2021).
5. Hyppönen, E., Vimaleswaran, K. S. & Zhou, A. Genetic determinants of 25-hydroxyvitamin D concentrations and their relevance to public health. *Nutrients* **14**, 4408 (2022).
6. Mancuso, N. *et al.* Probabilistic fine-mapping of transcriptome-wide association studies. *Nat. Genet.* **51**, 675–682 (2019).
7. Ongen, H. *et al.* Estimating the causal tissues for complex traits and diseases. *Nat. Genet.* **49**, 1676–1683 (2017).
8. Finucane, H. K. *et al.* Heritability enrichment of specifically expressed genes identifies disease-relevant tissues and cell types. *Nat. Genet.* **50**, 621–629 (2018).
9. Amariuta, T., Siewert-Rocks, K. & Price, A. L. Modeling tissue co-regulation estimates tissue-specific contributions to disease. *Nat. Genet.* **55**, 1503–1511 (2023).
10. Maier, T. & Hoyer, J. Modulation of blood pressure by central melanocortinergic pathways. *Nephrol. Dial. Transplant* **25**, 674–677 (2010).

11. da Silva, A. A., do Carmo, J. M., Wang, Z. & Hall, J. E. The brain melanocortin system, sympathetic control, and obesity hypertension. *Physiology (Bethesda)* **29**, 196–202 (2014).

12. do Carmo, J. M. *et al.* Role of the brain melanocortins in blood pressure regulation. *Biochim. Biophys. Acta Mol. Basis Dis.* **1863**, 2508–2514 (2017).

13. Wang, Y. & Wang, J.-G. Genome-wide association studies of hypertension and several other cardiovascular diseases. *Pulse (Basel)* **6**, 169–186 (2019).

14. Jacob, J., Chopra, S. & Baby, C. Neuro-endocrine regulation of blood pressure. *Indian J. Endocrinol. Metab.* **15**, 281 (2011).

15. Figueira, L. & Israel, A. Role of cerebellar adrenomedullin in blood pressure regulation. *Neuropeptides* **54**, 59–66 (2015).

16. Cvejic, A. *et al.* SMIM1 underlies the Vel blood group and influences red blood cell traits. *Nat. Genet.* **45**, 542–545 (2013).

17. Aniweh, Y., Nyarko, P. B., Quansah, E., Thiam, L. G. & Awandare, G. A. SMIM1 at a glance; discovery, genetic basis, recent progress and perspectives. *Parasite Epidemiol. Control* **5**, e00101 (2019).

18. Weeks, E. M. *et al.* Leveraging polygenic enrichments of gene features to predict genes underlying complex traits and diseases. *Nat. Genet.* **55**, 1267–1276 (2023).

19. van der Harst, P. *et al.* Seventy-five genetic loci influencing the human red blood cell. *Nature* **492**, 369–375 (2012).

20. Talmud, P. J. *et al.* Use of low-density lipoprotein cholesterol gene score to distinguish patients with polygenic and monogenic familial hypercholesterolaemia: a case-control study. *Lancet* **381**, 1293–1301 (2013).

21. Olmastroni, E. *et al.* Twelve variants polygenic score for low-density lipoprotein cholesterol distribution in a large cohort of patients with clinically diagnosed familial hypercholesterolemia with or without causative mutations. *J. Am. Heart Assoc.* **11**, (2022).

22. Immormino, R. M., Jania, C. M., Tilley, S. L. & Moran, T. P. Neuropilin-2 regulates airway inflammation in a neutrophilic asthma model. *Immun. Inflamm. Dis.* **10**, (2022).

23. Wang, B. *et al.* Alveolar macrophage-derived NRP2 curtails lung injury while boosting host defense in bacterial pneumonia. *J. Leukoc. Biol.* **112**, 499–512 (2022).

24. Arda, H. E. *et al.* Age-dependent pancreatic gene regulation reveals mechanisms governing human β cell function. *Cell Metab.* **23**, 909–920 (2016).

25. Bevacqua, R. J. *et al.* SIX2 and SIX3 coordinately regulate functional maturity and fate of human pancreatic β cells. *Genes Dev.* **35**, 234–249 (2021).

26. Zhao, S. *et al.* Adjusting for genetic confounders in transcriptome-wide association studies improves discovery of risk genes of complex traits. *Nat. Genet.* **56**, 336–347 (2024).
